# Excellence is a habit: Enhancing predictions of language impairment by identifying stable features in clinical perfusion scans

**DOI:** 10.1101/2023.09.13.23295370

**Authors:** Alex Teghipco, Hana Kim, Chris Rorden, Roger Newman-Norlund, Massoud Sharif, Darek Sikorski, Argye E. Hillis

**Affiliations:** University of South Carolina; University of Florida; Johns Hopkins University School of Medicine

## Abstract

Perfusion images guide acute stroke management, yet few studies have been able to systematically investigate CT perfusion collected during routine care because the measures are stored in proprietary formats incompatible with conventional research analysis pipelines. We illustrate the potential of harnessing granular data from these routine scans by using them to identify the association between specific areas of hypoperfusion and severity of object naming impairment in 43 acute stroke patients. Traditionally, similar analyses in such sample sizes face a dilemma—simple models risk being too constrained to make accurate predictions, while complex models risk overfitting and producing poor out-of-sample predictions. We demonstrate that evaluating the stability rather than out-of-sample predictive capacity of features in a nested cross-validation scheme can be an effective way of controlling model complexity and stabilizing model estimates across a variety of different regression techniques. Specifically, we show that introducing this step can determine model significance, even when the regression model already contains an embedded feature selection or dimensionality reduction step, or if a subset of features is manually selected prior to training based on expert knowledge. After improving model performance using more complex regression techniques, we discover that object naming performance relies on an extended language network encompassing regions thought to play a larger role in different naming tasks, right hemisphere regions distal to the site of injury, and regions and tracts that are less typically associated with language function. Our findings especially emphasize the role of the left superior temporal gyrus, uncinate fasciculus, and posterior insula in successful prediction of object naming impairment. Collectively, these results highlight the untapped potential of clinical CT perfusion images and demonstrate a flexible framework for enabling prediction in the limited sample sizes that currently dominate clinical neuroimaging.

## 1. Introduction

### 1.1 Clinical neuroimaging in stroke

Rapid medical advances over the last half-century have facilitated a sharp decline in stroke mortality in the United States, yet the resulting increase in stroke survivors is poised to present massive challenges in the coming years (Madsen et al., 2020; Gorelick, 2019). Stroke is presently the leading cause of severe long-term disability (Tsao et al., 2023) and its impact on quality of life surpasses that of most common medical conditions, including Alzheimer’s disease, cancer and even depression (Lam & Wodchis, 2010). Recent reports highlight a concerning rise in stroke incidence among younger and middle-aged adults, which is thought to reflect the growing prevalence of stroke risk factors such as obesity and diabetes (Ananth et al., 2022). These younger individuals have a long residual life expectancy, heralding a surge in people living with stroke related impairments. The consequences of a growing number of stroke survivors are far-reaching, extending beyond the affected individuals and their families and imposing a substantial economic burden on all parties, including healthcare systems (Rajsic et al., 2019).

Neuroimaging in particular has proven to be an indispensable tool for acute stroke management and may be key to improving stroke treatment and management strategies. Indeed, revolutionary recanalization therapies that are now standard of care were motivated by the observation that brain tissue that is receiving enough blood to survive but not enough to function (i.e., the ischemic penumbra, Astrup et al., 1981) can be salvaged and can restore function. Thus, stroke treatment is currently informed by neuroimaging techniques that identify the general location and extent of the ischemic penumbra (e.g., Adams et al., 2007; de Haan et al., 2006; Latchaw et al., 2009; Yew & Cheng, 2009). While endovascular therapy and thrombolysis have become widely used to restore blood flow to the penumbra, they carry some risks, including sometimes fatal hemorrhagic transformation of the infarct. Therefore, in weighing risks versus benefits of intervention, it is essential to predict what functions will recover if the area of hypoperfused tissue identified on imaging is reperfused. To this end it is necessary to bridge an important gap in knowledge about the functional correlates of specific areas of hypoperfusion. Here, we illustrate a successful approach to identifying the functional correlates of hypoperfusion identified with CT perfusion, by identifying areas of hypoperfusion in participants with left hemisphere acute ischemic stroke that are associated with acute impairments of object naming, arguably the most common cortical deficit in this patient group.

### 1.2 Clinical perfusion imaging

Clinical neuroimaging is essential for triaging patients for targeted interventions such as thrombolysis or thrombectomy (Adams et al., 2007; Powers et al., 2019). To that end, studies have emphasized the overlooked potential of perfusion imaging, which can be used to measure the dysfunctional but intact penumbra (Wintermark et al., 2002; Chalet et al., 2022). However, perfusion methods have traditionally lacked uniformity, leading to ambiguity regarding the best practices for acquiring, modeling, and thresholding perfusion data for clinical interpretation (Christense & Lansberg, 2019). While some methodological elements are becoming more established (e.g., Olivot et al., 2009), qualitative assessment of imaging data in the emergency department remains relatively common, despite demonstrably significant inter-rater variability (Barber et al., 2000; Campbell et al., 2015; Gupta et al., 2012). At the same time, quantitative identification and segmentation of the penumbra continues to be an active area of research and, generally, the aspects of the collected perfusion data that can be clinically relevant remain unsettled (e.g., Bani-Sadr et al., 2023; Forkert et al., 2013; Kamalian et al., 2012; Lu et al., 2022; Olivot et al., 2009; Wouters et al., 2017; Zaro-Weber et al., 2019). Considering this lack of consensus, it is remarkable that recent large-scale clinical trials have demonstrated some existing approaches are highly effective and can be exploited to extend treatment windows and improve patient outcomes (Albers et al., 2018; Kim-Tenser et al., 2021; Nougueira et al., 2018; Jovin et al., 2015; Goyal et al., 2015; Mocco et al., 2021).

Perfusion is most commonly estimated with Computed Tomography (CT). Although Magnetic Resonance Imaging (MRI) has higher sensitivity and specificity for stroke diagnosis, CT scanners are more widely available because they are both faster at acquiring images and cheaper to purchase, maintain, and use (Chalela et al., 2007). Processing the acquired images involves deconvolving each voxel’s time-attenuation curves using one of a variety of methods, ultimately resulting in perfusion parameter maps (e.g., Boutelier et al., 2012; Chalet et al., 2022; Ferreira et al., 2010; Yang, 2010; Wu et al., 2005). For example, such maps may include time to maximum (Tmax), which represents the time it takes for the contrast agent or blood flow to reach its maximum concentration in a region. Delays in this measure have been most frequently associated with delineation of the penumbra (e.g., Lin et al., 2014; Olivot et al., 2009) and it has been used for extending the therapeutic window of acute ischemic stroke interventions (e.g., Leigh et al., 2014).

Implementation of industry software solutions for automated data processing has helped standardize perfusion methods in the clinical setting. Nevertheless, software packages present significant variations in post-processing techniques, analysis strategies, and algorithms, introducing unique challenges (Austein et al., 2016). As an illustration, different software appears to have slightly different optimal Tmax thresholds for characterizing the penumbra, despite the fact that a single threshold has been widely recommended in the field (Bani-Sadr et al., 2023; Deutschmann et al., 2021). A more fundamental issue with these solutions is their reliance on proprietary code and technology. One consequence of this is the storage of processed perfusion data within opaque formats that preclude the pooling of granular perfusion data into larger groups, challenging application of state-of-the-art research analysis methods to understand relevant imaging features for evaluating the penumbra.

### 1.3 Machine learning as a new perspective on clinical neuroimaging data

Machine learning offers a novel data-driven perspective on the most consequential features of perfusion by maximizing the predictive accuracy of models seeking to relate whole brain perfusion measures to stroke outcomes. Emphasis on prediction represents a radical departure from traditional inferential statistics that has characterized biomedical research, where tightly controlled clinical trials are enshrined, and the impact of individual variations tends to be smoothed out during the modeling process in order to make more generalized conclusions about populations (e.g., Arbashiriani et al., 2017; Ashley, 2016; Bonkhoff & Grefkes, 2022; Bzdok et al., 2018; 2020; 2020; 2021; Bzdok & Ioannidis, 2019; Kosorok & Laber, 2019; Poldrack et al., 2019; Shmueli, 2010). Application of machine learning to clinical data affords the opportunity to advance more individualized clinical care (e.g., Bonkhoff & Grefkes, 2022; Bzdok et al., 2020; Kosorok & Laber, 2019). That is, effectively treating patients requires consideration of unique idiosyncrasies that can be irrelevant to understanding the average patients’ response to intervention. In judging the success of models on their ability to exploit statistical regularities that can generalize best to unseen *individual* patients, machine learning explicitly seeks to capitalize on more unique patient-specific qualities.

Machine learning approaches are particularly well-suited to the multivariate analysis of complex and wide imaging data (e.g., Arbashiriani et al., 2017; Bzdok et al., 2018), whereas the inferential methods that are more typically used (e.g., ordinary least squares linear regression) struggle in such settings (e.g., Harrell et al., 1996; Hastie et al., 2009; Simon et al., 2004; Wang et al., 2016). Not only do such inferential models require a remarkably large number of samples per feature to maintain relatively small variance around the parameter estimates, but their interpretation becomes complicated as the degrees of freedom increases, making it difficult to estimate the amount of model variance that can be attributed to an individual feature (Algina & Olejnik, 2000; Efron, 2012; Hastie et al., 2001; Knofczynski & Mundfrom, 2008; Miller and Kunce, 1973; Pedhazur & Schmelkin, 1991). This problem is amplified by the fact that these models generally neglect the impact of measurement noise, precipitating increased type 1 error rates in establishing incremental validity, particularly in large sample sizes and when the reliability of predictor measurements is moderate (Westfall & Yarkoni, 2016). Therefore, traditional inferential statistics of brain imaging data are typically applied in a mass univariate fashion where an independent estimate is calculated for each and every voxel of the brain in isolation. In contrast, multivariate machine learning can identify the patterns of injury across voxels that best predict behavior. In other words, machine learning embraces model complexity while leveraging mechanisms like regularization to discourage deleteriously complex models and distinguish signal from noise. For example, penalty terms weighted by out-of-sample errors improve the estimation of parameters used for prediction in high dimensional feature spaces (e.g., Bzdok et al., 2018; Heinze et al., 2018). In many cases, these algorithms are also explicitly designed to capture complex and nonlinear interactions with more flexibility and efficiency (e.g., Noble, 2006), enabling large-scale multivariate modeling robust to multicollinearities by, for instance, performing dimensionality reduction or feature selection during model estimation (e.g., Geladi & Kowalski, 1986; Zou & Hastie, 2005).

### 1.4 Applications of machine learning in acute stroke

Already, application of machine learning to acute stroke imaging is showing impressive achievements (see Sheth et al., 2023; Kamal et al., 2018 for reviews). First, studies have empirically demonstrated that prioritizing model prediction over inference can provide more realistic (i.e., lower) estimates of model variance attributable to factors like initial severity when predicting stroke outcomes, underscoring the importance of research that remains to be conducted (see Bonkhoff & Grefkes for review; see Bzdok et al., 2020 for more examples of this phenomenon in biomedicine). Moreover, a growing literature has applied machine learning principles developed in computer vision (e.g., convolutional neural networks) for quickly, automatically, and accurately identifying the presence of stroke within acquired brain images, as well as segmenting regions of interest such as the core lesion and penumbra. As one example, occlusion detection models that rely on CT angiography images have achieved remarkably high area under the curve in large out-of-sample datasets (0.91; Yhav-Dorvat et al., 2021). Such models have demonstrably reduced treatment times and have a potentially massive impact on health care costs (Hassan et al., 2020; Kunz et al., 2020). Although segmenting regions of the core and penumbra has proven to be more challenging, state-of-the-art performance achieves dice similarity scores of around 0.51 (Hakim et al., 2021). For comparison, private clinical software for perfusion analysis performs substantially worse (e.g., dice of ∼0.34) although it’s unclear why as the algorithms are not public.

While the more thoroughly explored goals of identifying and segmenting strokes have clear and immediately appreciable clinical utility, understanding how imaging features relate to outcomes can also be informative for clinical decision making (e.g., Sheth et al., 2023). Machine learning provides a powerful tool for understanding how perfusion within, but also outside of the penumbra may work together to deliver more accurate predictions of stroke outcomes. The presence of a small amount of potentially salvageable tissue may mask the amount of recovery that is made possible by other unique patient characteristics gleaned from whole brain perfusion data. For example, collateral flow is an important predictor of outcome and recent work shows CT perfusion can be used to quantify collateral flow in acute stroke with high accuracy (Lin et al., 2020). Further, peri-infarct depolarizations can cause disruptions in perfusion outside the penumbra and can provide additional predictive information to the model (Strong et al., 2007). Ultimately, regions extending beyond the penumbra may reflect factors that influence perfusion and improve model predictions. Many of these have been associated with outcome in prior work and include location of injury (e.g., Hope et al., 2013), rate of atrophy after stroke (Seghier et al., 2014), rate of age-associated morphological changes (e.g., Aamodt et al., 2023; Kristinsson et al., 2022; Seghier et al., 2014), existing comorbidities like white matter hyperintensities (e.g., Sagnier et al., 2020) and microbleeds (e.g., Sheikj-Bahaei et al., 2019), and individual variability in functional anatomy (Crafton, Mark & Cramer, 2003).

Studies relying on neuroimaging to predict outcomes have generally reported good accuracy for various measures, including those capturing motor (e.g., Rehme et al., 2014; Rondina et al., 2016), visual (e.g., Smith et al., 2013; Barrett al., 2019; Zhao et al., 2017), language (e.g., Hope et al., 2013; Yourganov et al., 2016), and gross impairments (e.g., Forkert et al., 2015; Moulton et al., 2019). Intriguingly, a number of studies comparing models have reported better outcome predictions when models had access to continuous information about the lesion (i.e., DTI axial diffusion maps instead of binary lesion segmentation maps; Moulton et al., 2019), and when models included data outside of the lesion (e.g., Ramsey et al., 2017; see also Bonkhoff & Grefkes, 2022 for review). Critically, few studies have targeted CT perfusion for predicting stroke outcomes and most have relied on extracting coarse information from perfusion parameter maps, such as the number of voxels within automatically segmented core and penumbra regions (e.g., Brugnara et al., 2020; Jabal et al., 2022; Jiang et al., 2021). Moreover, models using perfusion to predict outcome within specific cognitive domains are severely lacking, despite the well-established diversity of deficits that can emerge depending on the location of injury in stroke (e.g., Zhao et al., 2017).

### 1.5 Towards more predictive neuroimaging models with ensemble feature selection

The paucity of studies applying machine learning to granular CT perfusion features to predict stroke outcomes may be partly explained by the challenges associated with analyzing high-dimensional imaging data. Neuroimaging datasets suffer from the ‘curse of dimensionality’ as they typically have a much smaller proportion of samples to features, and only a small fraction of those features is usually expected to predict the response variable (e.g., Mwangi et al., 2014). These conditions force distances between features to become approximately equal, making them more challenging to distinguish during model estimation. Thus, paradoxically, the addition of information (i.e., features) can degrade a model’s performance by diluting the amount of signal that is available, causing models to overfit to noise (e.g., Jain, 2000; Rondina et al., 2014; Ransohoff, 2004). In addition, such large feature spaces can complicate model tuning as memory and computational requirements scale, leading to either intractably long tuning times or truncated search strategies that are more likely to find suboptimal solutions (e.g., Fan & Li, 2006). Strategies for removing unproductive features can mitigate these problems and produce more predictive models.

The success of many machine learning approaches in the neuroimaging domain stems directly from their ability to reduce the feature space during model estimation. For example, the widely used LASSO (Least Absolute Shrinkage and Selection Operator) adds an L1 penalty term to simple linear regression model’s loss function based on the absolute magnitude of the coefficients, shrinking many features to zero and eliminating them from the model during estimation (Tibshirani, 1996; see Mwangi et al., 2014 for review in the context of neuroimaging). Although other approaches have been proposed to solve some of the shortcomings of LASSO (e.g., Zou, 2006; Zou & Hastie, 2006), and there is no shortage of fundamentally different strategies to handling high-dimensional data during model estimation (e.g., Bi et al., 2003; Geladi & Kowalski, 1986), models ultimately face a trade-off between stability and solution sparsity (Xu et al., 2011). Consequently, feature selection can be unstable and may deteriorate models (e.g., Munson & Caruana, 2009; Way et al., 2010).

Other approaches to feature selection work as an independent step in the model building process. External feature evaluation adds more complexity by detaching the steps for model estimation and feature selection but can also be a powerful approach for improving model accuracy (e.g., Guyon et al., 2004). One illustration of this was described by Jollans and colleagues (2019), who modeled both simulated and real neuroimaging data using a variety of algorithms and sample sizes. These authors reported that the addition of an external feature selection step to the model building process that involved evaluating the stability and cross-validated accuracy of OLS linear regression models trained on each feature separately improved model accuracy when sample sizes were small (N<75). Although these authors also relied on model accuracy for feature selection, a handful of neuroimaging studies have eschewed accuracy altogether to explore the concept of identifying reliably predictive or associated features through stability analysis (e.g., Cribben, Wager & Lindquist, 2013; Fan & Chou, 2016; Liang et al., 2018; Ryali et al., 2012). By resampling the data many times, evaluation of feature stability may mitigate overfitting by measuring model robustness to noise and outliers (Meinshausen & Buhlmann, 2010). Intriguingly, some studies have shown that training models on neuroimaging features identified to be stable in this way can improve classification model accuracy (e.g., Rondina et al., 2014), and similar effects have been described outside of neuroimaging (e.g., Gopakumar et al., 2015; Seijo-Pardo et al., 2017).

A particularly versatile framework for evaluating feature stability is stability selection, which integrates the results of feature selection performed over many hyperparameter configurations, as well as models trained on different subsets of the data, in order to assemble an ensemble of the most robust features while also providing some error control for false positives in this stable feature set (Meinshausen & Buhlmann, 2010). This approach can be wrapped around any method for feature selection to identify consistently predictive features. While the approach does not consider the contribution of features to out-of-sample prediction error, it can be nested inside of a cross-validation scheme to identify reliable features that can stabilize predictive models.

While stability selection is conceptually straightforward, incorporating external feature selection into the model building process has proven challenging in practice for many clinical neuroimaging studies. Examples abound of feature selection being performed prior to nested cross-validation when developing models for reconstructing white matter tracts (Poulin et al., 2019), predicting psychiatric features from neuroimaging (Eitel et al., 2021; Kambeitz et al., 2017; Pulini et al., 2019; Whelan & Garavan, 2014), and predicting Alzheimer’s disease, autism, and traumatic brain injury from structural neuroimaging data (Mateos-Perez et al., 2018; Yagis et al., 2021). In one recent survey, as many as 10 of 57 neuroimaging studies surveyed performed dimensionality reduction before cross-validation (Poldrack et al., 2020). Similar forms of data leakage that are highly common present when the entire dataset is used to select significant features that will be modeled during cross-validation, or when some intercorrelated features are initially removed to reduce redundancy. While this leakage does not necessarily invalidate the findings that are reported, it often severely inflates model performance, injecting a large optimistic bias into model evaluations, and contributing to the ongoing reproducibility crisis (Kapoor & Narayanan, 2022; Rosenblatt et al., 2023)

### 1.6 Current study

The present work sought to demonstrate the potential of valuable data embedded within CT perfusion scans collected during routine care by showing that whole brain patterns of perfusion can be used to predict individual impairments on an object naming task. To enable this investigation, we first describe a method for extracting scalar perfusion measures from proprietary clinical images. We then demonstrate using simple linear regression with L1 and/or L2-norm penalties that evaluating the stability of features using the same kind of regression approaches as a preliminary model building step (i.e., using stability selection) can be a highly robust way of improving the predictive accuracy of models trained on relatively small samples of patients. Going further, we establish that evaluating feature stability using this approach outperforms models with embedded dimensionality reduction as well as alternative external feature selection methods that select features by minimizing out-of-sample error. After improving our predictions using more complex machine learning algorithms, which we show can also benefit from stability selection, we investigate the grey and white matter structures that contribute to good model predictions and relate them to the prior literature on the neural correlates of object naming.

Our results argue that stability selection can be highly effective for clinical neuroimaging studies, where small sample sizes are endemic (Szucs & Ioannidis, 2020), by generating many perturbed datasets across which noise can be better assessed to guard against model overfitting. This approach is highly flexible but has received limited attention in neuroimaging. To enable other researchers to more easily capitalize on this strategy for their studies, we develop and share a MATLAB toolbox that implements stability selection with over 14 different feature selection algorithms that can be used for regression and classification problems. In response to the rising misuse of feature selection in clinical neuroimaging, we package our stabSel toolbox with a variety of tutorials that not only demonstrate how stability selection can be appropriately used, but also how cross-validation schemes can be properly set up to avoid misuses such as data leakage.

## 2. Methods

### 2.1 Participants

Forty-three participants (19 women; mean age: 66.6 +/- 12.8 years) with acute ischemic stroke localized to the left hemisphere were enrolled at Johns Hopkins Hospital or Johns Hopkins Bayview Medical Center in accordance with the internal review board policies at Johns Hopkins University School of Medicine. All participants were adults, fluent in English prior to injury, had normal or corrected-to-normal vision and hearing, and had no history of previous stroke, dementia, or other neurological diseases. Absence of hemorrhage was confirmed with CT on presentation and CT perfusion was included in the imaging protocol. Perfusion images and Boston Naming Test (BNT) scores were obtained for every participant. More detailed information and additional demographics can be found in Table 1.

**Table 1:** Patient demographics and behavioral data.

| Participant | BNT | Days | Age | Sex | Educatio |
| --- | --- | --- | --- | --- | --- |

Stable perfusion features improve prediction of language impairment
|  |  | Post injury (scan) |  |  | n |
| --- | --- | --- | --- | --- | --- |
| P01 | 0 | 17 | 60s | F | 12 |
| P02 | 15 | 3 | 60s | F | Unknown |
| P03 | 53 | 1 | 70s | M | 18 |
| P04 | 97 | 2 | 60s | M | 20 |
| P05 | 70 | 2 | 70s | M | 12 |
| P06 | 36.67 | 3 | 70s | F | 11 |
| P07 | 86.7 | 1 | 50s | M | 12 |
| P08 | 0 | 3 | 70s | M | 13 |
| P09 | 27 | 1 | 40s | F | 14 |
| P10 | 90 | 2 | 70s | F | 14 |
| P11 | 0 | 4 | 60s | F | 18 |
| P12 | 91 | 1 | 50s | M | 12 |
| P13 | 3.33 | 1 | 40s | M | 12 |
| P14 | 46.7 | 1 | 70s | F | 18 |
| P15 | 0 | 4 | 80s | F | Unknown |
| P16 | 0 | 4 | 60s | F | Unknown |
| P17 | 87 | 0 | 70s | M | 18 |
| P18 | 0 | 0 | 60s | M | Unknown |
| P19 | 77 | 3 | 60s | M | 13 |
| P20 | 67 | 1 | 50s | M | 16 |
| P21 | 50 | 2 | 80s | F | 13 |
| P22 | 7 | 4 | 60s | F | 6 |
| P23 | 0 | 6 | 60s | M | 18 |
| P24 | 77 | 2 | 40s | M | 9 |
| P25 | 56.7 | 1 | 90s | F | 12 |
| P26 | 43.3 | 4 | 70s | F | 12 |
| P27 | 87 | 1 | 40s | M | 12 |
| P28 | 13.3 | 2 | 40s | F | 13 |
| P29 | 20 | 4 | 60s | F | 12 |
| P30 | 37 | 2 | 80s | F | 24 |
| P31 | 23 | 2 | 90s | F | 16 |
| P32 | 90 | 5 | 60s | M | 12 |
| P33 | 93 | 5 | 60s | M | 16 |
| P34 | 90 | 1 | 60s | F | 14 |
| P35 | 73 | 2 | 60s | M | 14 |
| P36 | 13 | 5 | 60s | M | 12 |
| P37 | 87 | 1 | 70s | M | 19 |
| P38 | 73 | 1 | 60s | M | 12 |
| P39 | 83 | 1 | 80s | M | 13 |
| P40 | 57 | 3 | 70s | M | 12 |
| P41 | 45.2 | 6 | 30s | M | Unknown |
| P42 | 0 | 2 | 50s | F | 12 |
| P43 | 80 | 1 | 80s | M | 11 |

### 2.2 Behavioral assessment

Object naming performance was evaluated using the BNT (Kaplan et al., 1983), a confrontation naming task commonly administered in the clinic (Kiran et al., 2018). Administration of the task followed previous work (Mack et al., 1992), with participants being instructed to name black and white line drawings of objects. Participants performed a short version of the BNT that includes 30 objects. Only the first uncued response was scored (mean BNT score: 47.6 +/- 35 points). In most cases, BNT performance was collected within one (N=16) to two (N=10) days of the stroke but occasionally up to 6 days later (N=11), and in a single case, 17 days later (mean days since stroke: 2.7 +/- 2.7 days).

### 2.3 Imaging data

Computed Tomography (CT) imaging was performed using a Siemens Somatom Drive (Siemens, Erlangen, Germany) at Johns Hopkins Hospital. Head CT images without contrast were collected, 512×512×49 voxels, 0.4×0.4×3 mm. Perfusion images were postprocessed with RAPID CT (RapidAI, Menlo Park, CA, United States), resulting in proprietary red-green-blue (RGB) images,256 x 286 x 15 voxels, 0.8×0.8×10mm, for multiple perfusion measures: time to drain, mean transit time, time to peak, cerebral blood flow, and cerebral blood volume.

### 2.4 Imaging data preprocessing

Signal-to-noise of the structural CT data was enhanced by creating a mean structural image using SPM12’s (Ashburner et al., 2012) realignment function. Structural data was deskulled with FMRIB’s BET (Smith, 2000) as described previously by Muschelli and colleagues (2015). No smoothing or thresholding of the data was performed at this stage. In cases where this pipeline did not work optimally, we were able to resolve brain extraction with the following adjustments: i) lowering the fractional intensity value below the default 0.1 when brain tissue was erroneously removed, ii) cropping the field of view when neck removal was difficult (bias field and neck cleanup performed less well), and iii) performing robust brain center estimation. In-house code was developed to quantify the measures contained within proprietary CT perfusion images by converting Siemens’ RGB color schemes (as embedded in image colorbars) to scalar intensities (https://github.com/neurolabusc/rgb2scalar). Note, the origin of both CT structural and perfusion images was set to the center of brightness. Deskulled structural images were warped to the scalar CT perfusion maps using SMP12’s coregistration routine (i.e., using normalized mutual information, sampling steps of 4 and 2mm, FWHM of 7mm). SPM12’s normalization routines were used to calculate the deformation required to transform a CT template from the clinical toolbox (Rorden et al., 2012) to the native space of the structural scan. In some cases, smoothing the template by an additional 2mm substantially improved normalization (the default smoothing was set to 8mm, participant data were not smoothed). The deformation was used with nearest neighbor interpolation to bring the JHU-MNI atlas (Faria et al., 2012) into register with participant anatomy while preserving discrete regions. In a final step, participant-level perfusion maps were mean scaled using perfusion in the right hemisphere (i.e., by multiplying each value in the map by the quotient of 100 and mean perfusion across the right hemisphere). Regional perfusion measures were finally extracted for 189 grey and white matter regions of the JHU-MNI atlas by averaging across voxels belonging to the same region. Code for replicating this pipeline can be found at: https://github.com/alexteghipco/CTPerfusionPipeline.

### 2.5 Overview of entire model building procedure

A range of complementary regression algorithms were tested for their ability to predict BNT scores from regional CT perfusion: ridge, lasso, elastic net, principal component, partial least squares, support vector, random forest, and gaussian process regression. Because the number of features in our data (i.e., different perfusion measures within brain regions; N = 756) vastly exceeds the number of samples (N=43), thereby increasing the risk of overfitting, we elected to constrain our feature space to a single perfusion measure: time-to-maximum (Tmax; 189 features).

Although the algorithms that we tested exploited all Tmax features, we also applied the same algorithms to smaller subsets of features in an effort to further improve model performance, reduce model complexity, and decrease model training and tuning time. Thus, algorithms were also tested on theory-defined Regions of Interest (ROIs) based on prior studies on object naming, as well as feature subsets identified with a range of different data-driven feature selection methods, including wrappers (i.e., sequential forward selection using linear regression and elastic net, stepwise linear regression using the Bayesian Information Criterion, Akaike Information Criterion, sum of square errors, and the adjusted R2, stability selection using elastic net, and genetic algorithm), filters (i.e., strongest correlations, most stable correlations across subsamples, significant correlations, RreliefF and neighborhood component analysis algorithms) and embedded methods (i.e., lasso, elastic net, partial least squares with variable importance projection). See Figure 1 for visual overview of the methods we employed and supplemental materials for more details.

**Figure 1.**
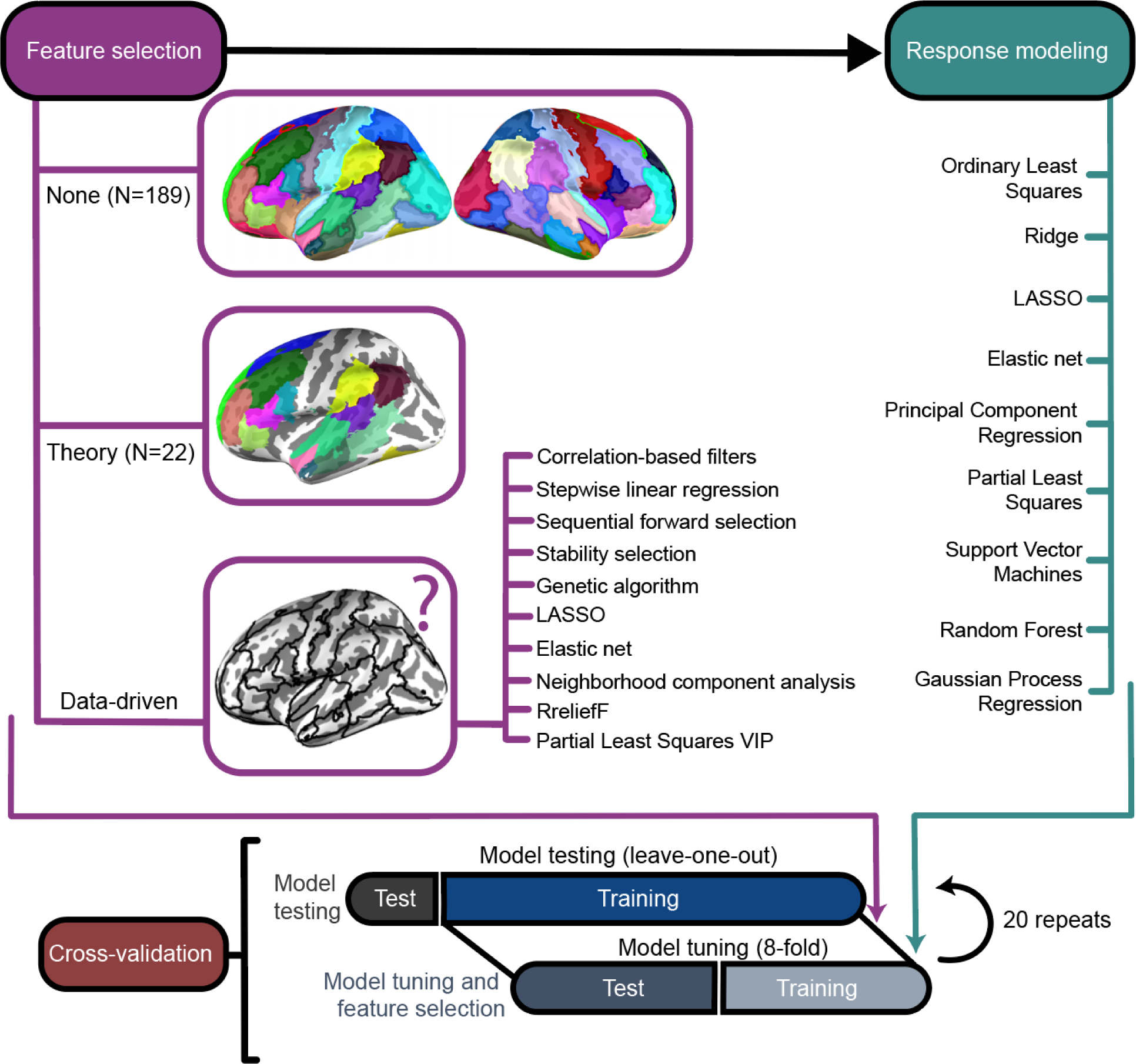
Overview of model building pipeline. Three general approaches to feature selection (i.e., retaining all features, theoretically motivated feature selection, and data-driven feature selection; in purple on left) were paired with 10 different regression modeling algorithms (i.e., in teal on the right). Top surface rendering visualizes all structures (i.e., features) in the dataset. Structures were extracted from the JHU atlas and projected onto a cortical surface using RF-ANTs purely for visualization (Wu et al., 2018). Each structure is assigned a persistent color across all figures that attempts to maximize perceptual distinction in “lab” space (Holy, 2023). Middle surface rendering visualizes features previously identified to be part of the naming network. Bottom surface rendering visualizes only outlines of features to symbolize the uncertainty of which features will be identified as important by data-driven selection methods. It is accompanied by the full list of data-driven feature selection methods that were tested. The bottom panel shows a schematic of the cross-validation scheme. Leave-one-out was used for estimating model performance and 8 inner folds were used for tuning. Hyperparameters for feature selection were tuned alongside other hyperparameters for the regression models in the inner folds. Cross validation was repeated 20 times.

### 2.6 Cross-validation

Point-estimates of model performance were measured over 20 repeats of leave-one-out cross-validation (LOOCV). An inner 8-fold cross-validation loop was used to tune models. Cross-validation (CV) was nested to mitigate overfitting, which is more likely to occur in smaller samples and while tuning more hyperparameters (Hosseini et al., 2020; Varoquaux, 2018). In small samples, repeating CV can mitigate the influence of unrealistically optimistic partitions drawn by chance while maintaining sufficient test sizes within the CV scheme (Cearns, Hahn, Baune, 2019). For more complex machine learning algorithms with more hyperparameters, tuning was further stabilized using 30 monte-carlo style repartitions of the inner folds (i.e., for support vector, random forest, and gaussian process regression).

Partitions were preallocated to facilitate comparisons between different models. Performance was measured by computing the Pearson correlation coefficient between model predictions from the outer LOOCV loop and true BNT scores. Although it is a highly popular goodness-of-fit measure in predictive models treating neuroimaging data, correlation can poorly reflect a model’s predictive performance because it is translation and scale invariant, sensitive to outliers, insensitive to nonlinearities and biased in the case of LOOCV (e.g., Poldrack et al., 2019). Consequently, we also evaluated model performance using the accuracy percent measure, which expresses in percentage units the mean absolute error of predictions scaled to the error of a naïve model that guesses based on the median of the training data (e.g., Franses, 2016). Paired t-tests over repeats of the entire cross-validation procedure were used to determine significant differences in performance between models.

### 2.7 Feature modeling and selection overview

All features submitted to models were normalized inside the cross-validation loop. That is, normalization was independently performed in the inner and outer loops and test data was always normalized using the training data distribution. In most cases, normalization involved z-scoring features to have 0 mean and standard deviation of 1, except in the case of support vector regression, where features were rescaled to vary from 0 to 1.

First, models were trained using theory-driven language ROIs (N=22), which were identified in the left hemisphere using published work on the neural correlates of object naming and included inferior frontal (e.g., Indefrey a & Levelt, 2004), middle frontal (e.g., Farias et al., 2005), superior frontal (e.g., Farias et al., 2005), superior temporal (e.g., Baldo et al., 2013), middle temporal (e.g., Baldo et al., 2013), fusiform (e.g., Farias et al., 2005), supramarginal (e.g., Breining et al., 2022) and angular gyri (e.g., Seghier, Fagan, Price, 2010) as well as superior and middle temporal poles (e.g., Farias et al., 2005), the thalamus (Keser et al., 2021) and the splenium of the corpus collosum (e.g., Hinkley et al., 2016). Note, some gyri spanned multiple regions in the atlas (e.g., middle temporal gyrus and posterior middle temporal gyrus). White matter tracts related to language were included as well and comprised the uncinate fasciculus, superior longitudinal fasciculus, sagittal stratum (which includes portions of the inferior longitudinal fasciculus and inferior fronto-occipital fasciculus), and inferior fronto-occipital fasciculus (e.g., Baldo et al., 2013; Dick et al., 2014; Keser et al., 2021). Studies emphasize the importance of left hemisphere regions but given the potential for greater right hemisphere involvement in the context of left hemisphere damage (e.g., Cao et al., 1999), theory-driven ROIs were extended to cover right hemisphere homologues in some analyses.

The same models were trained on features identified in a purely data-driven way. Feature selection was always implemented inside the cross-validation scheme and blind to the test data. In cases where feature selection required tuning, a grid search was performed. Feature selection hyperparameters were entered into the inner cross-validation loop and tuned alongside the hyperparameters for the regression model as described below (i.e., for feature selection with lasso, elastic net, correlation filter with tuned window, RreliefF, neighborhood components analysis, forward sequential selection). In the case of some filter methods that required no tuning as well as some wrapper methods that relied on resampling techniques, feature selection was performed on all the training data and only tuning of the regression model occurred in the inner cross-validation loop (i.e., for stability selection, genetic algorithm, stepwise regression, sequential feature selection, significant and high magnitude correlations as a filter). To mitigate underestimation of errors in cross-validation, prediction accuracy was evaluated in reference to a null distribution built over 1000 random permutations (e.g., Varoquaux, 2018). In each permutation, the entire model building procedure, including feature selection, was repeated.

#### 2.8.1 External feature selection algorithms

Several simple correlation-based filters were tested. In the most straightforward approach, Pearson correlation coefficients were generated between perfusion in each of 189 regions of the brain and BNT scores. This was performed independently in each training dataset and the 30 highest absolute coefficients were selected to build a regression model using the inner cross-validation loop. In a slightly more complex feature selection procedure, the number of top correlation coefficients retained for model building was tuned in the inner loop using ordinary least squares (OLS) regression. Mean absolute error on inner test folds was used to select from among a range of filter windows (i.e., 5 to 70). In another procedure, p-values associated with the correlation coefficients were used to determine the window, retaining all features significant at p < 0.05 (see supplemental section 1.4 for more information on thresholds).

In another approach, stepwise regression methods were used to automatically add features to an ordinary least squares regression model built on training data. That is, starting with a constant model, features were added that maximized different criteria as measured in-sample for the training data. Here, we tested using BIC, AIC, adjusted R^2^ and significant difference (i.e., p < 0.05) in sum of squared errors on an *F*-test. Sequential forward feature selection was also performed. This was conceptually identical to stepwise regression but operated over the inner cross-validation loop so that cross-validation error (mean absolute error) could be used to determine the addition of features (see supplemental section 1.5 for more details).

Two feature selection methods conceptually related to the k-nearest neighbor algorithm were also used: RreliefF (Robnik-Sikonja & Kononeko, 1997) and neighborhood component analysis (NCA; Yang et al., 2012). RreliefF is a nonlinear algorithm that aims to penalize features that diverge across neighbors with similar response values and to reward features that diverge across neighbors with different response values. We tuned the *k* number of nearest neighbors in rReliefF using 15 values between 1 and 15. Because RreliefF is a filter method that simply ranks features, we also tuned the filter window (identical range to section 2.8.1). NCA is a linear distance learning method that aims to learn a Mahalanobis distance measure which would maximize a k-nearest neighbor prediction on the training data by using a stochastic neighbor selection rule, ultimately shrinking some feature weights to zero. The NCA regularization hyperparameter was tuned over 101 linearly spaced values between 0 and 100. We also tuned the ε value for the loss function (15 linearly spaced values between 0 and twice the interquartile range of the response variable). For more information see supplemental section 1.7)

#### 2.8.2 Algorithms with embedded feature selection

Simple OLS regression using either the L1-norm penalty term (i.e., Lasso; Tibshirani, 1994) or a combination of L1 and L2-norm penalty terms (i.e., elastic net; Zou & Hastie, 2005) were also used for feature selection. While the L1-norm penalty shrinks features to zero, it cannot retain more features than there are samples in the data and can fail to select the full set of predictive features in the presence of strong noise or collinearities (e.g., Meinshausen & Buhlmann, 2010; Zou & Zhang, 2009). In contrast, the L2-norm penalty (i.e., ridge regression; Hoerl and Kennard, 1970) only shrinks coefficients towards zero but assigns similar values to correlated features (Friedman et al., 2010). Combining these penalty terms provides greater flexibility during modeling but introduces a hyperparameter that determines the weight given to one term over the other. For all of these methods, a total of 1000 log-distributed λ values were generated between 0 and the highest possible value that would return a non-null model. The weight of L1/L2 norm penalty, a, was tested with 21 linearly spaced values ranging from 0.0001 (i.e., ridge regression) to 1 (i.e., lasso).

#### 2.8.3 Algorithms with embedded dimensionality reduction

Some regression models reduce the number of features in a dataset by projecting them onto a lower dimensional space. Principal component regression (PCR) involves performing principal components analysis over the feature space and regressing the resulting components on the response variable using ordinary least squares (e.g., Jolliffe et al., 1982). Unseen data is projected into the principal component space of the training data and the regression model fit to the training data is tested on the new samples. Here, we tuned the number of components *c* in PCR to retain in the lower-dimensional space (each *c* value from 1 to 25 was tested). PCR aims to predict the response variable from orthogonal components that maximize explained feature variance. Partial least-squares (PLS) regression is a similar technique that involves performing least squares regression over component scores. PLS is distinguished by the fact that the response variable is used to identify scores that have large covariance between features and the response variable (Jong, 1993). This can permit PLS to explain variability in the response variable using fewer components. We used PLS while tuning the number of components *c* using the same range as PCR. PLS may also be used to perform feature selection using the variable importance in projection (VIP) measure (Chong & Chi-Hyuck, 2005). VIP scores are the weighted sum of squares of PLS weights and reflect the amount of variance explained in the response variable by the feature as projected into shared component space. Consistent with common recommendations, we refit the PLS model to features that scored above 1 on VIP (Chong & Chi-Hyuck, 2005).

#### 2.8.4 External ensemble feature selection algorithms

Stability selection is a framework for ensemble feature selection that wraps around any feature selection method to identify stable features while providing finite sample error control for false discoveries (Meinshausen & Buhlmann, 2010). Core to this framework is the application of a feature selection method to many differently perturbed versions of the same dataset across the entire range of possible hyperparameters. Aggregation of selected features across datasets and hyperparameters shapes the definition of a stable feature set of more consistently selected features that more robustly predict the response variable. The stable set may be formed based on an arbitrary consistency threshold (e.g., a feature must be selected in more than half of perturbed datasets) or informed by a predetermined per-comparison or per-family error rate.

Perturbation of datasets typically follows a subsampling procedure that draws half the samples in the dataset, but a larger proportion of the dataset may be drawn when sample sizes are small (Meinshausen & Buhlmann, 2010). For more details on stability selection, see supplemental section 1.8.

Recent studies have demonstrated stability selection to work well in a variety of contexts (e.g., Ahmed et al., 2011; Gilhodes et al., 2020; Hofner et al., 2015; Hyde et al., 2022; Lu et al., 2017; Ryali et al., 2012; Yin et al., 2022). Here, we apply stability selection within a nested cross-validation scheme to improve predictive models. Although lasso is commonly used in stability selection (e.g., Meinshausen & Buhlmann, 2010; Ryali et al., 2012), we elected to use an elastic net to avoid potential issues caused by collinearity (see section 2.8.2 above). We also initially tested stability selection using a simple correlation filter that always selected the 30 highest Pearson correlation coefficients in a subsample.

We implemented stability selection by developing a comprehensive MATLAB package that supports this framework in the context of 14 different regression-based feature selection methods (https://github.com/alexteghipco/StabilitySelection). In the current study, stability selection was performed using the originally proposed subsampling scheme. Data were resampled 200 times with each subsample randomly retaining 40 participants (93% of the data) to account for the relatively low sample size. Per-family error rate was set to p < 0.05. When stability selection was used with correlation as a filter, *q*Λ (i.e., number of retained features in each subsample) was set to 30 and the appropriate π_thr_ (i.e., threshold used to form the stable set) was automatically determined (see supplemental section 1.8 for full formulation). When using an elastic net, *q*Λ was indirectly determined by Λ (i.e., the set of λ regularization hyperparameters tested). Definition of Λ and *a* was identical to the description in section 2.8.2.

We additionally tested a genetic algorithm for ensemble feature selection. Genetic algorithms (GA) are population-based search algorithms that can be used to select features by treating different models as chromosomes and simulating the process of natural selection. Models that improve prediction reproduce and evolve. Here, we used simple OLS regression as our models and the genetic algorithm selection strategy described by Leardi & Lupianez (1998). Briefly, 30 chromosomes were randomly defined across the data, with each chromosome containing 30 features on average. Root mean squared error was used as the fitness function in conjunction with an 8-fold cross-validation procedure (venetian blinds). Best models replicated over 100 generations to identify relevant variables. The evolutionary process was repeated 200 times. The probability of mutation was always 10% and the probability of cross-over was 50%. The frequency with which each variable was selected in the best model of each repeat of the evolutionary process was computed for feature selection. Features were retained if they were selected more than 8 times. This value was chosen as it produced feature subsets comparable in size to stability selection.

### 2.9 Predictive modeling

The feature selection procedures described above were paired with regression algorithms of varying complexity to make predictions about object naming deficits, including OLS, OLS with the L2-norm penalty (ridge regression), OLS with the L1-norm penalty (LASSO), OLS with a combination of L1 and L2-norm penalties (elastic net), Principal Component Regression (PCR), Partial Least Squares (PLS), Support Vector Regression (SVRs), Random Forests (RFs), and Gaussian Process Regression (GPR) models (see Figure 1 for visual overview). Including methods that further reduced the feature space *after* feature selection permitted multiple feature selection strategies to shape or refine a feature set. While not guaranteed, this has the potential to mitigate the risk of obtaining false-positive features by relying on a secondary feature selection process that uses new data to inform the feature set and remove redundant or noise variables (e.g., Yarkoni & Westfall, 2017).

Lasso, ridge and elastic net regression models were constructed in the same way described in section 2.8.2. PLS and PCR models were constructed as outlined in section 2.8.3. Multiple SVR models were independently tuned using different kernels to evaluate whether model complexity could improve prediction (i.e., SVR with linear kernel, 2^nd^ order polynomial kernel, 3^rd^ order polynomial kernel and gaussian kernel). Linear SVRs share similarity to Lasso and can produce comparable solutions (Jaggi, 2013) but aim to find a hyperplane of best fit to the response variable using a different objective function—the L2-norm of the coefficient vector. To accomplish this, a maximum acceptable error term, D, is tuned for accuracy. As some errors may fall outside D, slack variables are introduced to capture deviations from the margin. An additional hyperparameter, *C*, tunes the tolerance of the model to such deviations. We tuned log scaled D values between 1e-100 and 6. In MATLAB, higher *C* values assign stronger penalties to deviations from the margin, resulting in fewer support vectors at the cost of longer training times. We tested a range of linearly spaced *C* values between 10,000 and 100,000. In SVR, the kernel trick is used to efficiently transform data into a higher dimensional space through which a hyperplane can be more successfully optimized using different kernel functions. Using the gaussian kernel introduces a third hyperparameter, D, for defining the kernel radius. The D parameter was tuned using a different and probabilistic method for efficiency, by measuring deviation across subsampled training datasets on a quality criterion (see Scholkopf & Smola, 2001).

We also developed RF models, which are an ensemble regression method that integrates predictions across many weak decision tree learners. Decision trees are trained on bootstrapped datasets that have as many samples as there are in the dataset (i.e., bagging). To improve the accuracy of bagged trees, every tree in the ensemble randomly selects predictors for each decision split (Breiman et al., 2001). Here, we implemented RF regression while tuning the number of weak learners (i.e., ranging from 10 to 500), as well as their minimum leaf size (i.e., ranging from 1 to 94) and maximum number of splits (i.e., 1 to 188). RFs tend to reduce overfitting as averaging uncorrelated trees (i.e., with random predictors) lowers overall variance in estimation (e.g., Ghojogh & Crowley, 2019).

Finally, GPR is a probabilistic Bayesian method for implementing gaussian processes for regression (e.g., Schulz et al. 2018). In GPR, a gaussian process is used to describe a distribution over the infinite number of functions that can be used to fit the training data (i.e., gaussian process can be equated to an infinite dimensional multivariate Gaussian). At minimum, GPR requires defining a kernel that can be used to describe the shapes of the functions fit to the data, as well as the initial value for the noise standard deviation that describes the probability density, σ^2^. We tuned σ^2^ within the range of 1e-4 and 200 and the optimal kernel function from a total list of 10 (i.e., exponential, squared exponential, matern32, matern52, and rational quadratic kernels with and without separate length scale per predictor). GPR has the benefit of providing uncertainty measures around predictions, which researchers have exploited for normative modeling (e.g., Rutherford et al., 2022).

### 2.10 Evaluating feature importance with Shapley Additive Explanations

As we tested many different machine learning algorithms for predicting object naming deficits, we evaluated feature importance using Shapley Additive Explanations (SHAP), an algorithm agnostic method that would accommodate the whichever model performed best. Shapley values are a game theoretic approach for quantifying the average marginal contribution of a player in a cooperative game (e.g., Roth, 1988). That is, the Shapley value for a feature describes its role in deviating the prediction from the average or baseline prediction with respect to a specific sample in the data. SHAP is an extension of Shapley values that uses the conditional kernel with k nearest neighbors (corresponding to 10% of the samples) for evaluating feature importance (Aas et al., 2021). Critically, this formulation of SHAP does not assume feature independence.

## 3. Results

### 3.1 Predicting naming scores from perfusion in the left hemisphere object naming network

The overarching goal of the present research was to generate a model that could successfully predict object naming impairments in acute stroke from regional perfusion measures collected during routine care. To that end, we first evaluated whether prior knowledge of regions contributing to object naming performance combined with simple ordinary least squares (OLS) multiple linear regression could produce a successful model. At this stage, we focused on left hemisphere regions of interest (ROIs) only (N=22) as OLS tends to fail when dimensionality exceeds sample size (e.g., Wang et al., 2016). Prior to fitting the model, we computed pairwise Pearson correlation coefficients between object naming performance and perfusion in our ROIs to confirm that the regions we selected are reasonably associated with the response variable and that the model may be successful in principle (Figure 1). We found that 68% of our preselected ROIs exhibited a statistically significant relationship after false discovery rate correction (p < 0.05), with additional regions trending towards significance. Individually, the ROIs explained at most 22% of the variance in object naming. Significant relationships were all inverse, showing that higher Tmax, reflecting delayed collateral supply and possible hypoperfusion, was associated with worse object naming performance. Cross-validated predictions made by the model in LOOCV were not significantly correlated with true BNT scores, r(41) = 0.13, p = 0.4. Mean absolute errors were 29% higher compared to a naïve model guessing based on mean training scores (i.e., accuracy percent, or AP; see Figure 1). A follow-up analysis showed that even if the model were fit to the entire dataset without cross-validation, it would not be significant and would not contain any significant terms (p > 0.05), despite generating high correlation between in-sample predictions and true scores, r(41) = 0.73, p < 0.0001.

### 3.2 Improving model prediction with simple feature selection methods

Correlations presented in Figure 2 suggest data-driven feature selection can enhance model performance. Our ROIs showed significant correlations with object naming, but also included plausibly uninformative features not associated with naming scores while also skirting unspecified features that explained a substantial amount of variance. Before attempting to maximize the predictive accuracy of our model using more complex approaches, we tested whether the strategy of simultaneously removing noise features while adding more signal to our set of ROIs by relying on simple data-driven feature selection would show evidence of model improvement. Eleven different methods were applied to the entire feature set before fitting a linear regression model with OLS in order to determine which strategies may be more successful. Model prediction errors within a nested LOOCV scheme were compared across methods, but also to a naïve guessing model (i.e., AP). Three control or baseline models were used for further context on model performance: i) theory-driven ROI model from Figure 2, ii) the same model extended to cover right hemisphere homologues, and iii) an OLS model without feature selection.

**Figure 2.**
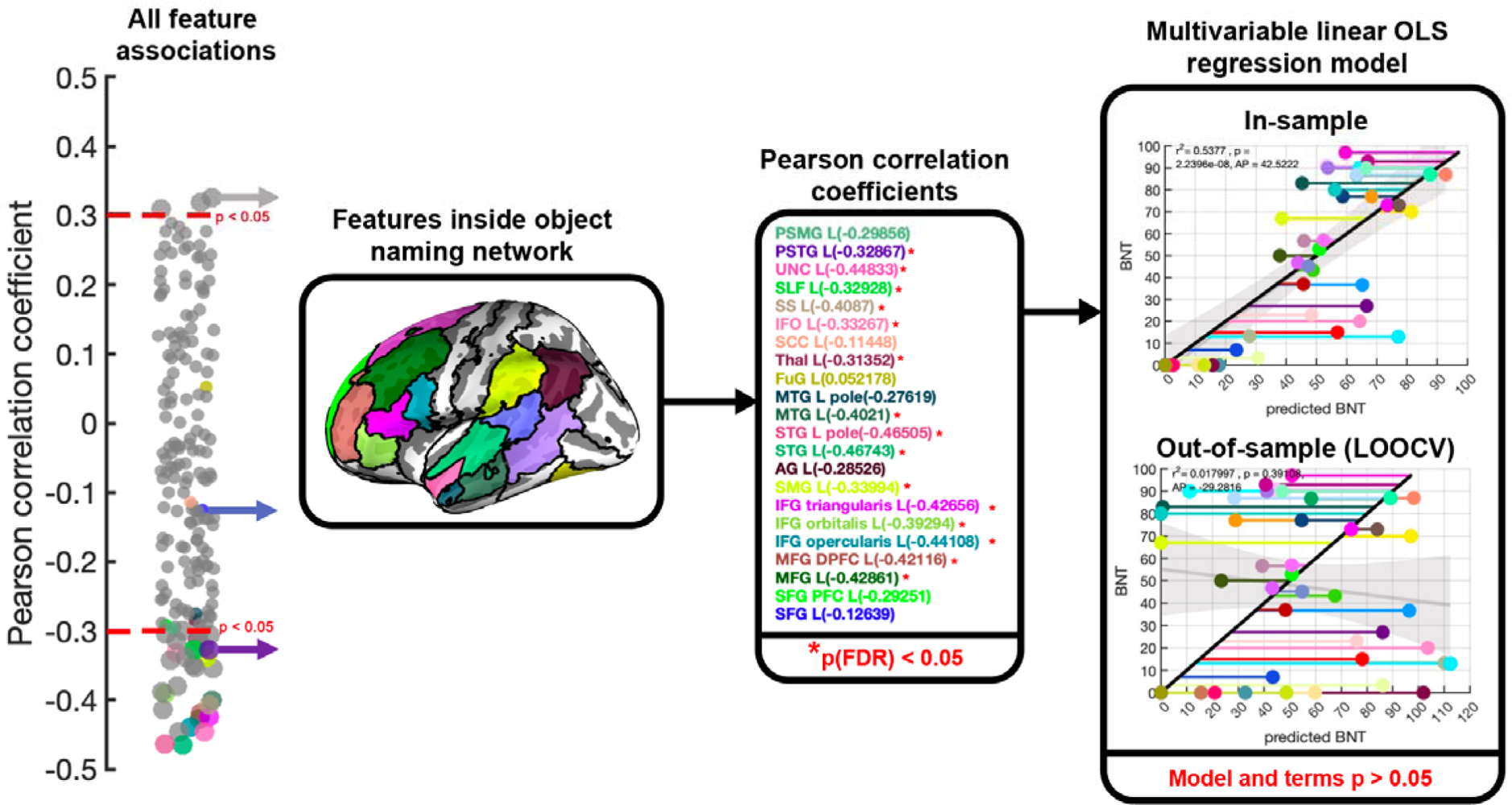
Predicting naming impairment with multivariable linear regression of perfusion in the object network. Pearson correlation coefficients between object naming performance and each feature in the dataset are presented in the swarm chart on the left. Each dot, or feature, corresponds to Tmax within a region of our chosen atlas. Red dotted lines on the swarm chart represent significance thresholds and significant features are also emphasized by increased dot size. Features that did not intersect with our set of theory-driven ROIs were assigned a grey color on the swarm chart. A visualization of the left hemisphere language ROIs is presented in the leftmost black box. The Pearson correlation coefficient for each theory-driven ROI is presented in the middle black box, along with information about which correlations survive multiple comparisons correction. The rightmost black box describes the performance of an OLS multiple linear regression model for predicting object naming scores when fit using all of the theory-driven ROIs. The topmost scatterplot shows in-sample model predictions, and the bottom scatterplot shows model predictions in a leave-one-out cross-validation scheme. Each dot represents a single participant and is assigned a persistent color across figures. The grey line and patch represent the trendline and functional bounds of a linear regression of predictions on true BNT scores. The black diagonal line represents perfect predictions and horizontal lines emphasize model residuals. Superimposed on each scatterplot is an r^2^ term and p-value representing the amount of variance explained in BNT scores by the model predictions and whether this explanation was statistically significant. In addition, accuracy percent (AP) is presented, showing model performance in percent relative to a naïve model that guesses based on the mean of the data.

Initially, we determined that any model improvement over the left hemisphere theory-driven ROI model from Figure 2 would not be due to the omission of right hemisphere regions (black and green colored dots in Figure 3A). A paired two sample t-test showed a significant difference between absolute prediction errors for the left hemisphere ROI model (M=41.2, SD = 30.3) and the bilateral ROI model (M=106.2, SD = 129.1), t(42) = −3.6, p < 0.0001. Using all features (M = 55, SD = 53.3) led to smaller errors than the bilateral ROI model, t(42) = −2.8, p < 0.01 and prediction error was not significantly different than the left hemisphere ROI model, t(42) = −1.5, p = 0.13. Although there was no significant difference between left hemisphere ROI and no feature selection models (p > 0.05), left hemisphere ROIs performed substantially better relative to a naïve model (−30% AP for LH ROI model and −72% for no feature selection model). Consequently, we focus on the left hemisphere ROI model as the primary baseline model. Note, that while we report correlation coefficients for models in Figure 3A, we overall observed a negative association between AP and correlation performance measures across simple feature selection methods, r(12) = −0.65, p = 0.01. We give greater weight to AP for evaluating models as we believe it is a more intuitive measure of the predictive performance of models (see methods for more details). For more comprehensive comparisons between models, see supplemental Figures S2, S3, and Table S1.

**Figure 3.**
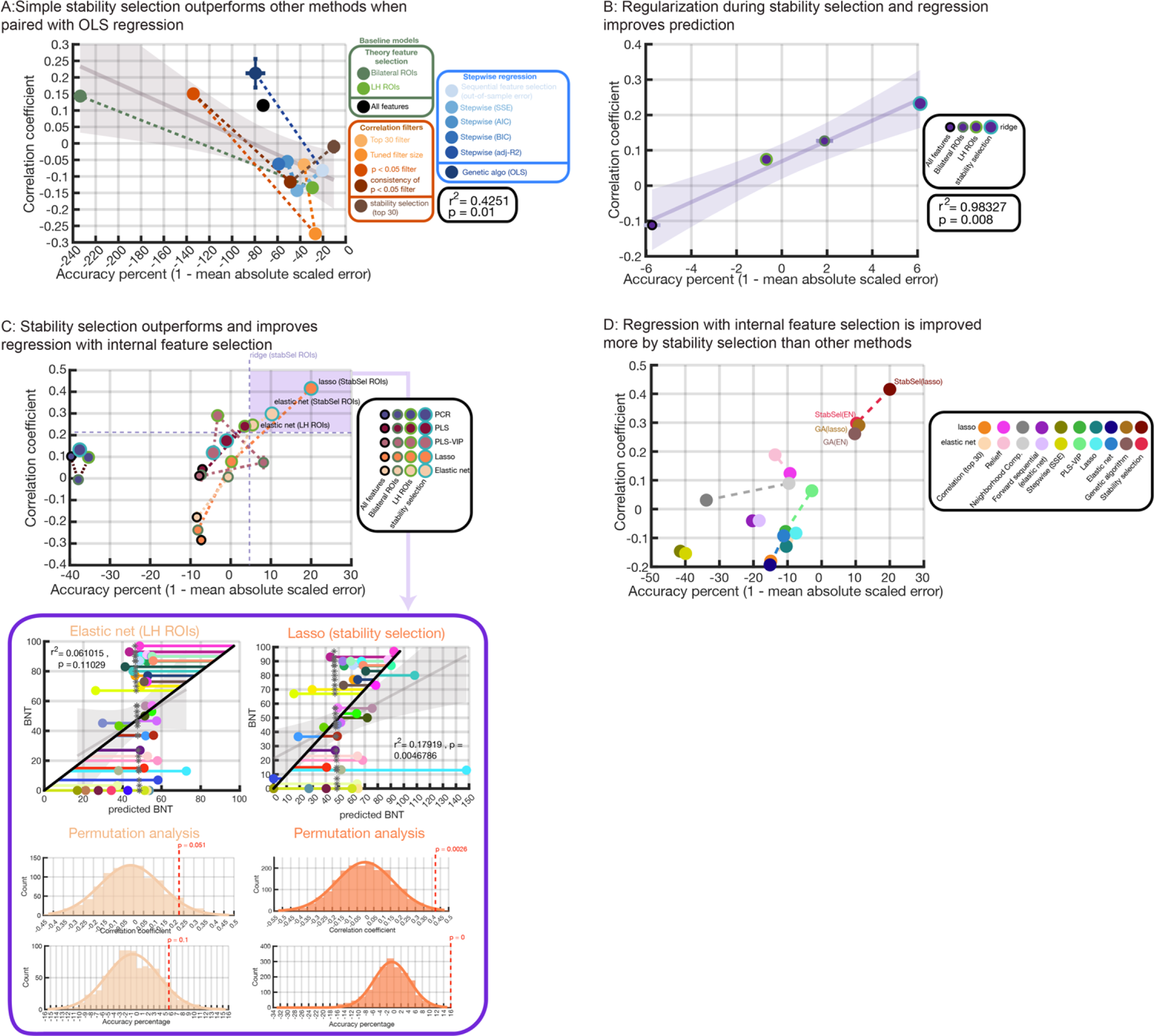
Improving prediction of naming impairment with stability selection. Relatively straightforward feature selection methods are used to improve the simple OLS regression model in **panel A**. Where possible, each method is implemented in a nested-cross-validation procedure and repeated 20 times. Median model performance across repeats is plotted according to correlation between predictions and true values (y-axis) and model error scaled by a naïve model that guesses based on the mean (x-axis). Negative values on accuracy percent represent worse performance than the naïve model. Error bars represent standard error of the median. Baseline models either using no feature selection or theory-driven feature selection are shown in black and green. Feature selection methods that rely on computing the Pearson correlation coefficient between features and object naming scores are shown in orange. Greedy search algorithms are presented in blue. Stability selection is shown in brown. A genetic algorithm is shown in the darkest blue color and grouped with greedy algorithms for similarity. Each category of methods is connected by dashed lines to emphasize differences. The grey line and shaded areas represent the functional bounds of a linear regression between AP and correlation across methods. **Panel B** shows the performance of a model where feature selection is made more complex by applying an elastic net in the stability selection framework and introducing regularization into model estimation for prediction (i.e., using ridge regression). Theory-driven ROIs as well as no feature selection are included as baseline methods (i.e., with regularization applied during model estimation). Dot outlines and sizes correspond to a model’s feature selection method (i.e., smallest dot with black outline corresponds to the no feature selection model, larger dot with dark green outline corresponds to the bilateral theory-driven ROIs model, larger dot with light green outline corresponds to left hemisphere theory-driven ROIs model, and largest light blue dots correspond to stability selection). **Panel C** shows the performance of baseline feature selection methods and stability selection, each paired with regularization methods for estimating the model that embed a feature selection (i.e., lasso and elastic net) or dimensionality reduction (i.e., PLS, PLS-VIP, PCR) step. Colors of dots correspond to the kind of regression model that was estimated for prediction. Dot sizes and outline colors correspond to the feature selection method. Dashed lines isolate models that outperformed stability selection with ridge regression. The top performing theory-driven ROI and stability selection models were submitted for permutation testing, the results of which are shown in the purple box. This box additionally contains scatterplots between the median performing models’ predictions and true object naming values. **Panel D** presents more complex feature selection methods paired with lasso and elastic net for estimating the regression model. Each feature selection method was assigned a unique color. The lighter shade of that color represents pairing that feature selection method with elastic net for model estimation. The darker shade of that color represents a pairing with lasso regression.

We next evaluated forward stepwise regression with different criteria for feature selection (Figure 3A). Stepwise regression with sum of square errors (SSE) performed slightly better (AP= −43%) than other in-sample criteria (BIC: AP = −59%, AIC = −52%, and adjusted R^2^ = −52%) relative to a naïve model. However, there were no significant differences in absolute errors (e.g., SSE vs AIC: SSE M = 45.5, SD = 34.1; AIC M = 50.6, SD = 46.2; t(42) = −1.125, p = 0.27). The SSE model had slightly lower AP than the left hemisphere ROI model, but showed no difference in absolute errors, t(42) = −0.91, p = 0.37. In a final test, features were selected within each training set of an inner cross-validation loop, using mean absolute error on the test sets as the objective function (i.e., forward sequential selection). This approach generally improved the model, increasing AP to −23% when averaging across repeats (i.e., CV was repeated only when it was possible to tune models as LOOCV would otherwise produce identical results on repeat). However, absolute errors for the median performing model across repeats were not lower than stepwise regression with SSE, t(42) = −1.2, p = 0.23, or the left hemisphere ROI model, t(42) = −0.41, p = 0.68.

We also tested correlation filters (Figure 3A). The simplest approach selected the top 30 features in each training dataset based on the absolute magnitude of the Pearson correlation coefficients with object naming scores (see supplemental section 2.2 for justification of 30 features). This approach performed roughly as well as the left hemisphere ROI model based on both AP (−36%) and absolute errors (M =43.5, SD = 48.3), t(42) = −0.3, p = 0.7. Tuning the filter window size within an inner CV loop improved AP (−25%), but did not produce significantly lower errors (M=40.3, SD=24.8), t(42) = −0.432, p = 0.67. Absolute errors were not lower compared to the left hemisphere ROI model, t(42) = −0.17, p = 0.86. Statistical significance of correlation as an adaptive criterion for filter window size was tested as well (i.e., retaining features with correlation of p < 0.05). This approach performed poorly, achieving - 140% AP and showing significantly higher absolute errors (M=74.5, SD=97.5) than the left ROI model, t(43) = 2.38, p < 0.05. In analyses not formally reported here, lowering this threshold and/or applying multiple comparison correction produced comparable results.

To gauge the utility of evaluating feature stability, we tested whether a simple approach for measuring the consistency of significant correlations would reduce prediction error. The proportion of times that a feature survived the p < 0.05 threshold across 8 inner folds repartitioned 30 times (i.e., using the test folds to perform correlation) was recorded. The regression model was fit to features appearing in >50% of folds. The resulting model did not show significantly lower absolute errors (M=47.5, SD=35) compared to the approach that ignored stability, t(43) = −1.8, p = 0.08. However, evaluating stability had a positive impact, substantially increasing model AP (−48%). Thus, we next combined stability analysis with the top 30 correlation filter, which was surprisingly effective relative to other methods while also being one of the fastest to compute. Stability selection streamlined this approach, subsampling the data several hundred times instead of repartitioning an inner CV loop. A stable feature set was formed by selecting a threshold that attempted to ensure 1 or fewer false positives (see methods). The resulting model had the highest AP of all tested models (−10.4%). However, the median performing model across 20 repeats of CV did not produce significantly different absolute errors (M=35.1, SD=28.6) than the left hemisphere ROI model, t(43) = −1.46, p = 0.15, or the tuned filter window size model, t(43) = −1.37, p = 0.18. Finally, a genetic algorithm was tested, which shares the property of being an ensemble approach, and tended to perform relatively poorly according to AP (−80%) and significantly worse (M=57.2, SD=61) than stability selection in terms of absolute errors on the median performing model, t(43) = −2.9, p < 0.0001.

### 3.3 Introducing regularization to feature selection and model fitting

Accuracy percent indicated that stability selection wrapped around a top 30 correlation filter outperformed other simple feature selection methods when paired with OLS linear regression, including the use of theory-driven ROIs as feature selection. Because this strategy still performed worse than a naïve model, we tested whether adding regularization to both feature selection and model fitting would improve performance (Figure 3B). Elastic net was used to select features within the stability selection framework and ridge regression was used to fit the model. As before, baseline models were used to contextualize performance. All compared models required tuning and had stochastic elements. Thus, model comparisons (i.e., paired t-tests) were carried out over performance measures (i.e., AP and correlation) tracked across 20 repeats of CV.

First, we found that stability selection with elastic net and ridge regression produced higher AP (M=5.5,SD=3.4) than the correlation-based stability selection model from Figure 3A (M=-9.35,SD=3.4), t(19) = 12.5, p < 0.000001. Moreover, AP showed that stability selection with elastic net outperformed the left hemisphere ROI (M=-0.9,SD=1.25), t(19) = 7.9, p < 0.0001, bilateral ROI (M=2.04,SD=2.3), t(19) = 3.8, p < 0.01, and no feature selection models (M=-6,SD=2.2), t(19) = 13.3, p < 0.0001 with ridge regression. Comparisons based on prediction correlations to true values showed the same patterns (see supplemental Table S2, Figure S4 and S5). Both stability selection and bilateral ROI models performed better than a naïve model. All models with regularization had higher median AP and correlation performance measures compared to their counterparts from Figure 3A. With the substantial gain in performance afforded by ridge regression, correlation and AP measures were positively associated across models (c.f. Figure 3A), r(2) = 0.99, p < 0.01.

We next tested the possibility that embedded feature selection and dimensionality reduction methods would generate better predictions than stability selection (e.g., using an elastic net for feature selection *and* model estimation, without relying on an external feature selection step; Figure 3C). Virtually all the methods we tested scored between −7 and −8.5% on AP for the median performing model, including PLS, PLS-VIP, elastic net and lasso. All of these models performed significantly worse than stability selection with ridge regression, demonstrating that an external feature selection step can be more effective (highest attained p-value was 0.005). Remarkably, no data-driven method that combined feature selection or dimensionality reduction with model estimation outperformed a simple ridge regression fit to all features in the dataset (p > 0.05). For more detail and pairwise tests see supplemental material (including Figures S6 and S7, and Table S2).

The ridge model fit to stability selection ROIs represents a purely data-driven approach to feature selection. We assessed whether it would be more productive to remove noise features from theory-driven ROIs without considering the addition of other ROIs (i.e., using PLS, PLS-VIP, PCR, lasso, and elastic net). This strategy performed significantly better than stability selection with ridge according to AP only when PLS-VIP was fit to bilateral ROIs (p < 0.0001). The second-best performing approach fit an elastic net to left hemisphere ROIs but did not perform significantly better than stability selection according to AP (p > 0.05). On correlation, only PLS-VIP applied to left hemisphere ROIs outperformed stability selection (p < 0.0001). PLS and elastic net applied to left hemisphere ROIs both performed as well as stability selection according to correlation (p > 0.05). When considering AP and correlation in combination, only elastic net and PLS fit to left hemisphere ROIs approximated the performance of stability selection with ridge regression. Further, elastic net tended to outperform PLS on both measures and performed significantly better on AP (p < 0.00001; see purple box, Figure 3C). For more thorough comparisons see supplemental material (including Figures S6 and S7, and Table S2).

Stability selection with ridge regression may have fared worse because like the preselected ROIs, it may have identified some noisy or redundant features. If this were the case, fitting a model with embedded feature selection or dimensionality reduction to features identified by stability selection should improve performance in the same way. Indeed, this was the case when models embedded feature selection but not dimensionality reduction. For example, stability selection ROIs paired with elastic net for model fitting improved median model AP by 4% compared to ridge regression (i.e., up to ∼10% AP). This model had higher AP than left hemisphere theory-driven ROIs paired with elastic net, but the mean AP difference was not statistically significant (p > 0.05, see Figure S6 and S7). Note, left hemisphere ROIs performed significantly better than bilateral ROIs or no feature selection when the model was fitted with either lasso or elastic net. Critically, stability selection ROIs paired with lasso a achieved the highest AP of any model we tested (20%). This performance was significantly better than all other models (p < 0.0000000001; see Figure S6 and S7). When we performed permutation testing of the best overall models that were built with stability selection and theory-driven ROIs (i.e., left hemisphere ROIs with elastic net and stability selection ROIs with lasso), we found that the stability selection model exhibited both AP and correlation performance measures that were significantly better than chance (p < 0.01), whereas the theory-driven model had AP comparable to chance (p = 0.1), and showed a correlation coefficient which was on the cusp of significance (p = 0.05). We note a pattern whereby stability selection did not perform better than theory-driven ROIs for methods with embedded dimensionality reduction, likely because they benefit from a greater number of features for estimating latent dimensions. PCR performed especially poorly overall. Finally, we report that for most methods, correlation and AP were associated positively and significantly across repeats (Figure S10). This association was either insignificant, or the inverse pattern was observed, but only for methods that tended to perform poorly, suggesting that correlation can be misleadingly high when models make particularly large prediction errors.

### 3.4 Testing stability selection against more complex feature selection approaches

In a final test of its utility, we determined whether stability selection ‘supervised’ by lasso or elastic net could be outperformed by other feature selection methods combined with the same model fitting procedure (Figure 3D). We found that most alternatives performed significantly worse (see Figure S8). The one exception to this was a genetic algorithm paired with elastic net regression, which did not show significantly different performance than stability selection with elastic net according to AP (p > 0.05), but which performed worse according to correlation (p < 0.01). We emphasize, further, that stability selection with lasso outperformed all alternative combinations of feature selection and model fitting (p < 0.0000001). Remarkably, we found that using lasso to estimate the initial set of features, then refining that set and estimating the regression model simultaneously with elastic net performed slightly better than most methods (AP of −12%). Initial feature selection with PLS-VIP followed by elastic net regression performed even better (AP −3%), but not as well as stability selection or the genetic algorithm. A more comprehensive comparison between feature selection methods, including some of the approaches we previously leveraged in Figure 3A (e.g., top 30 correlation filter, stepwise regression with SSE, genetic algorithm), similar approaches amended in an effort to improve predictions (e.g., forward sequential stepwise regression, but using elastic net rather than OLS), and novel approaches (e.g., neighborhood component analysis and RreliefF) can be found in Figure S8 and S9.

We qualitatively demonstrate why stability selection is effective when paired with lasso for model estimation in Figure 4, where we plot lasso coefficients for every training dataset and highlight the features that stability selection consistently identified as stable across all datasets. This figure illustrates that stability selection adapts to the unique training datasets to which it is applied, either removing features identified as stable across most training datasets or adding features deemed unstable across most training datasets. Lasso acts as a second filter, removing features that may have been deemed stable, but which do not facilitate prediction based on new out-of-sample data. For example, Figure 4 shows lasso assigns very high weights to some features that are stable in one or two training datasets, demonstrating that stability selection effectively adapts to the training dataset in a way that facilitates prediction of unseen samples. At the same time, lasso downweighs many features identified to either be stable across many training datasets or unique to some training datasets. We speculate stability selection would identify fewer unpredictive features in larger sample sizes or more homogenous patient groups, where subsampling a smaller proportion of the data would be more successful (i.e., lower subsampling proportions in our small dataset resulted in unproductively small stable sets). Overall, we found no statistically significant relationship between the tendency for a model to have higher prediction errors (i.e., above average versus below average error) and the selection of low consistency features during stability selection (i.e., the presence of features that entered the stable set in fewer than 20% of training datasets), X^2^ (1, N=43) = 0.001, p = 0.97. However, models with below-average prediction error were significantly more likely to have low consistency features *with* higher assigned lasso coefficients (i.e., >50^th^ percentile of absolute weights), X^2^ (1, N=43) = 4.13, p < 0.05. Further, an association between the number of low-consistency features identified by stability selection and model error generally trended towards significance, r(41) = −0.2, p = 0.18.

**Figure 4.**
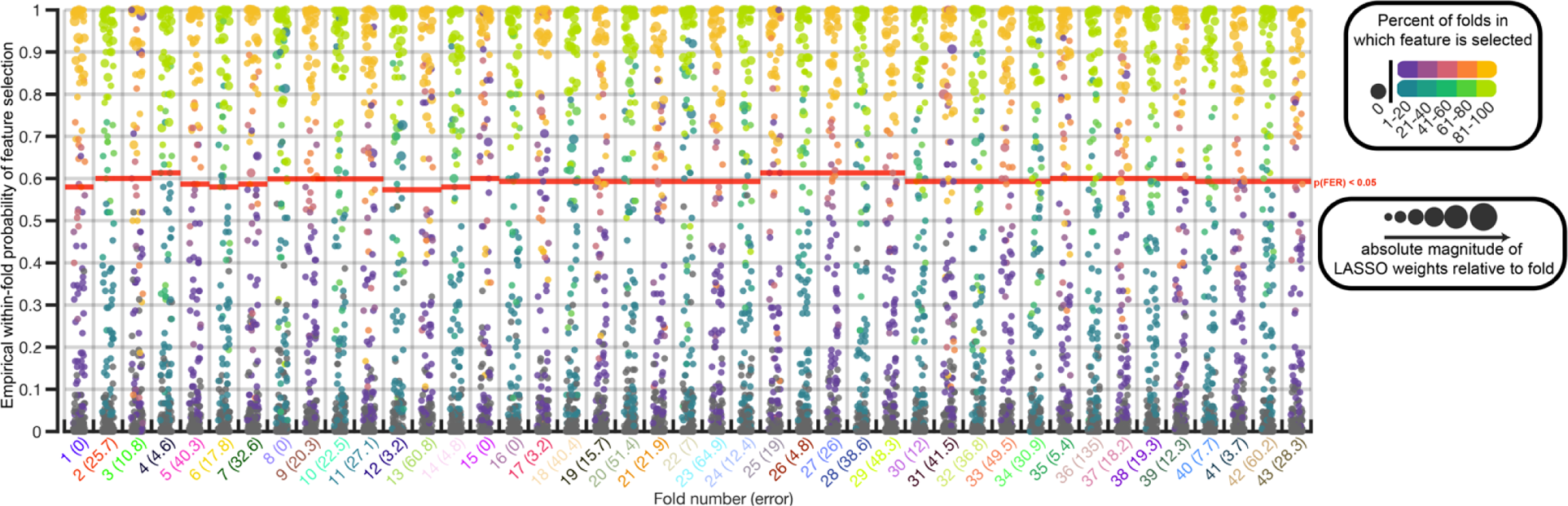
Qualitative visualization of stability selection and lasso for weighting features for prediction. Consistency of feature selection as computed across subsamples (y-axis) is presented separately for each training dataset (x-axis). That is, the proportion of subsamples in which a feature was selected during stability selection with elastic net. Red lines represent the p(FER) < 0.05 cutoff determined by stability selection. Features, or dots, above this line form the stable set for a training dataset. Text identifying each fold reflects persistent subject colors. Parenthetical text shows the prediction error for the model fit to a particular training dataset. Each feature is colored according to the percentage of training datasets (i.e., folds) in which it appears in the stable set. Color schemes alternate for each training dataset to help emphasize patterns. Darker colors represent features that rarely entered the stable set across training datasets. Lighter colors represent features that frequently entered the stable set. The dots that represent each feature are also varied by size to reflect the weight assigned to the feature by lasso regression (i.e., median performing model across repeats). Note, stability selection had high confidence in assigning some features to the stable set that were rarely selected across datasets. Many of these features had a large impact on the lasso regression model (i.e., large dark dots that score well above the red line on the y-axis). On the other hand, some features frequently entered the stable set across training datasets but had a low probability of being in the stable set for particular training datasets (i.e., lighter colored dots scoring low on the y-axis). Many stable features were also assigned low weights by lasso in some datasets (i.e., small dots that score high on the y-axis).

### 3.5 Improving predictions by pairing stability selection with more complex machine learning algorithms for model fitting

In a final attempt to improve model performance, we paired stability selection with more complex machine learning algorithms, including random forest regression (RFR), gaussian process regression (GPR) and support vector regression (SVR) with different kernels. These methods are multivariate, have complementary theoretical foundations that may idiosyncratically boost performance (see methods), and in most cases require tuning more parameters than other methods we have tested, which may result in more robust learning. Remarkably, we found that only GPR outperformed stability selection with lasso regression (Figure 4A). GPR increased AP by roughly 3% (to 23%) and the correlation coefficient by roughly 8% (to 0.51) based on median model performance across 20 repeats. Mean differences in model performance across repeats were statistically significant (p < 0.01; see Figure S11). SVR with a gaussian kernel also performed relatively well, but significantly worse than both GPR and lasso (p < 0.01; see Figure S11). The worst performance was observed for linear SVR and RFR, with median RFR model performance being indistinguishable from naïve guessing. Polynomial and linear SVR performance was not significantly higher or lower than gaussian SVR based on the correlation coefficient (p > 0.05; see Figure S10), but significantly worse on AP (p < 0.05; see Figure S10). Supplemental Figures S10 and S11 show performance distributions for these models and more exhaustive model comparisons. Permutation testing indicated both the GPR and gaussian SVR model predictions were statistically significant (Figure 2B). We speculate that GPR may have been more effective than other methods partly because it required less tuning (i.e., two parameters, in line with linear SVR, except the second parameter for GPR had only 10 possible values). However, we stress that with less complex algorithms tuning more parameters did not necessarily result in worse model performance (see supplemental section 2.5).

To test whether demographics could significantly impact model performance (e.g., interacted with brain features), demographic features (see Table 1) were added to stability selected features and the model tuning procedure was repeated for GPR. The resulting model had slightly lower AP (i.e., based on the median performing model) but showed no statistically significant difference across repeats when compared to the GPR model that excluded demographics (p = 0.285; see Figure S13). However, the GPR model with demographics produced significantly lower correlations with true data (p < 0.01; see Figure S13). As a final test, we confirmed the genetic algorithm paired with GPR did not improve model performance as this was the only other data-driven feature selection method that performed relatively well in prior analyses (p < 0.0001; see Figure S13). As the more complex feature selection methods we use in this analysis may be more robust in higher dimensional datasets, we also confirmed in supplemental analyses (see Figure S14) that none of the models in Figure 4A performed worse than the same models trained on all the data or left hemisphere ROIs (the latter of which tended to outperform bilateral ROIs paired with L1 and/or L2-norm penalty with OLS regression).

**Figure 5.**
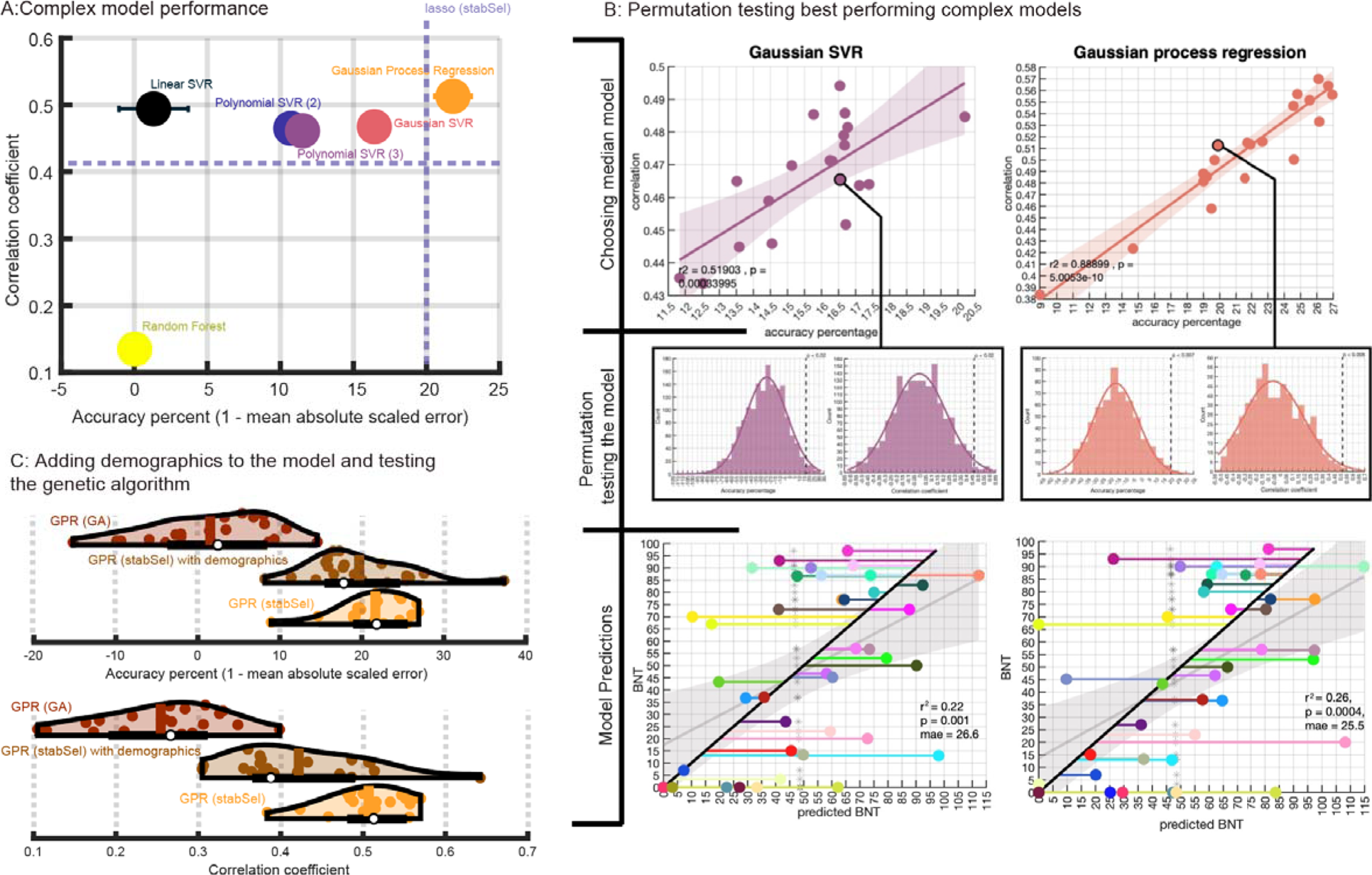
Testing more complex machine learning algorithms with stability selection. More complex machine learning algorithms were paired with stability selection for feature selection. **Panel A** shows median model performance for different machine learning algorithms (assigned random colors) with standard error of the median as measured across 20 repeats of the nested cross-validation procedure for model building. Dotted purple lines represent the performance of stability selection when paired with lasso to emphasize any model improvement. **Panel B** shows a scatterplot of model performance across 20 repeats of the model building procedure for the two best performing regression algorithms (gaussian SVR and GPR). The median performing models in each scatterplot are outlined and connected to distributions of model performance across 1000 permutations of the response data (model were re-tuned in the same way for each permutation). Model predictions are presented in the bottom two scatterplots. **Panel C** shows violin plots of model performance for the best performing regression algorithm (GPR; orange), performance of the same algorithm re-trained with additional demographics data (brown), and the same algorithm re-trained with another top performing data-driven feature selection method (genetic algorithm; red). The topmost plot shows performance as measured by accuracy percent. The bottommost plot shows performance as measured by the correlation between true and predicted naming scores.

The direct impact of stability selection on more complex machine learning models is presented in Figure 6. Notably, fitting RFR models to stability selection ROIs did not produce significantly better performance than fitting RFR models to all features according to AP or correlation (p > 0.05). However, using stability selection ROIs did lead to significantly better performance for GPR (see supplemental Figure S15, S16 and Table S6). These findings confirm that stability selection can be used as a preprocessing step in the model fitting process to generate more successful models, even when modeling involves state-of-the-art machine learning approaches.

**Figure 6.**
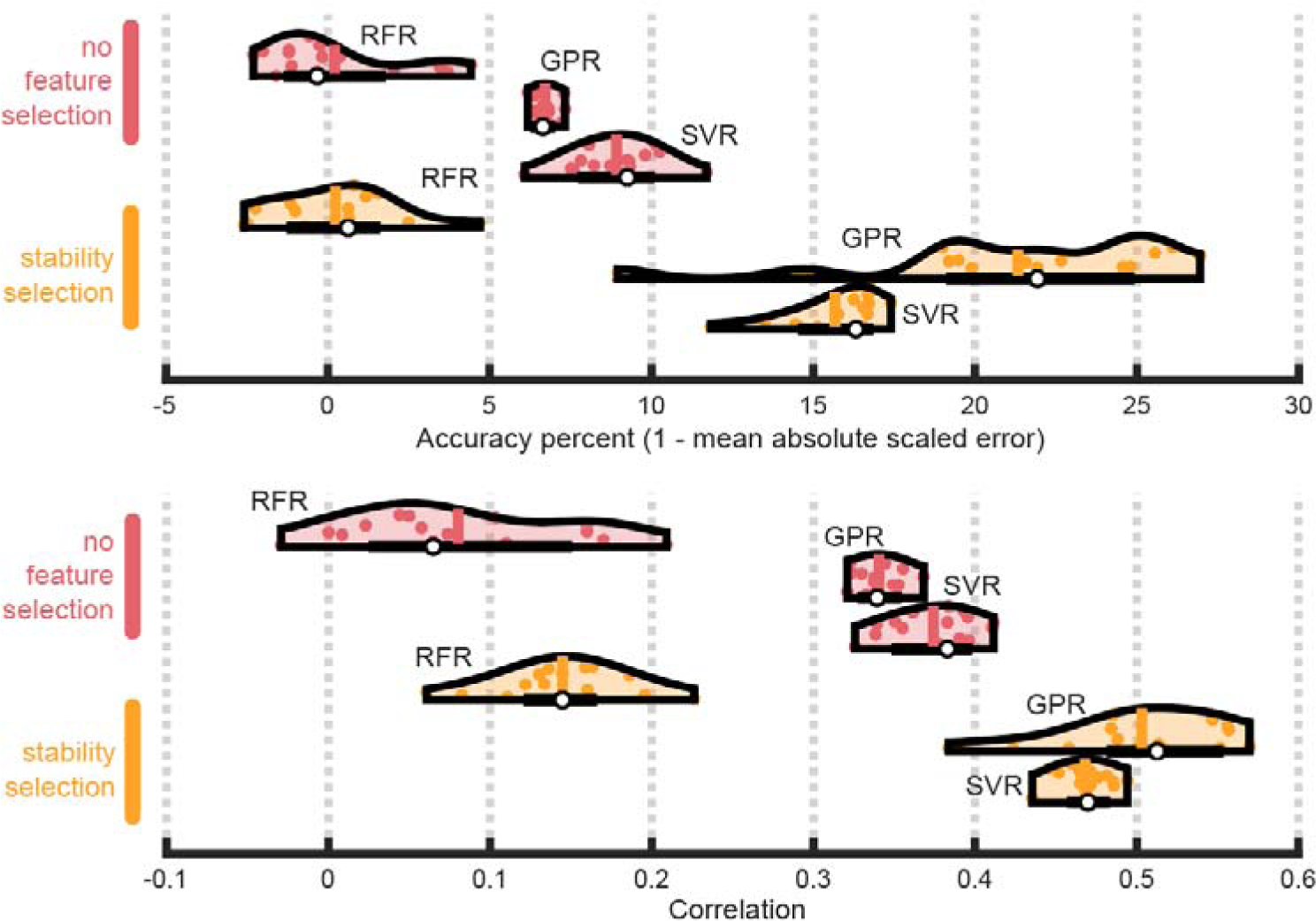
Impact of stability selection on more complex machine learning algorithms. Top panel shows violin plots representing the distribution of model performance as measured by accuracy percent over 15 repeats of cross-validation (not all models were cross-validated 20 times so we restrict analysis to the first 15 repeats). Bottom panel shows violin plots, but performance is measured by correlating true naming scores with predicted scores. In both panels, the white dot represents the median performing model (i.e., out of 15). The vertical line inside each violin shows the mean performing model. Other colored dots inside the violin represent the remaining models’ performance. Black horizontal lines represent a box plot. Violin colors correspond to the ROI set to which different machine learning algorithms were fit: yellow is theory-driven ROIs (i.e., left hemisphere ROIs), salmon is all ROIs (i.e., no feature selection), orange is stability selection ROIs. Within each set of violin plots (i.e., belonging to the same color), the topmost violin shows performance of random forests (RFR), middle violin shows performance for gaussian process regression (GPR), and bottommost violin shows performance for support vector regression (SVR).

### 3.6 Explaining prediction of object naming impairments

The median performing GPR model was interrogated with SHAP. SHAP values were collapsed across both cross-validated models (i.e., in the outer CV loop) and samples to generate global measures of feature importance. We first analyzed absolute SHAP values, which reflect the total influence that a feature exerts on model predictions. Note, whenever SHAP values were manipulated, the operation was performed prior to collapsing them. These values are visualized separately for grey matter (Figure 7A) and white matter (Figure 7B) structures. The most influential features were distinguished by identifying absolute SHAP values above the 90^th^ percentile, which retrieved 29 regions (see Table 2 for full list and SHAP values). Values precipitously dropped after the top 8 features, which was confirmed by programmatically identifying the knee in values sorted by descending order (i.e., finding the largest Euclidean distance between each value and a line that connected the first and last values). Top influential regions in descending order of importance were: the left ansa lenticularis (lAns), left superior temporal gyrus (lSTG), left pars triangularis segment of the inferior frontal gyrus (lTri), left posterior insula (lPIns), left lingual gyrus (lLing), left superior temporal gyrus pole (lSTGP), left occipital lateral ventricle (lOLV), and left insula (lIns).

**Figure 7.**
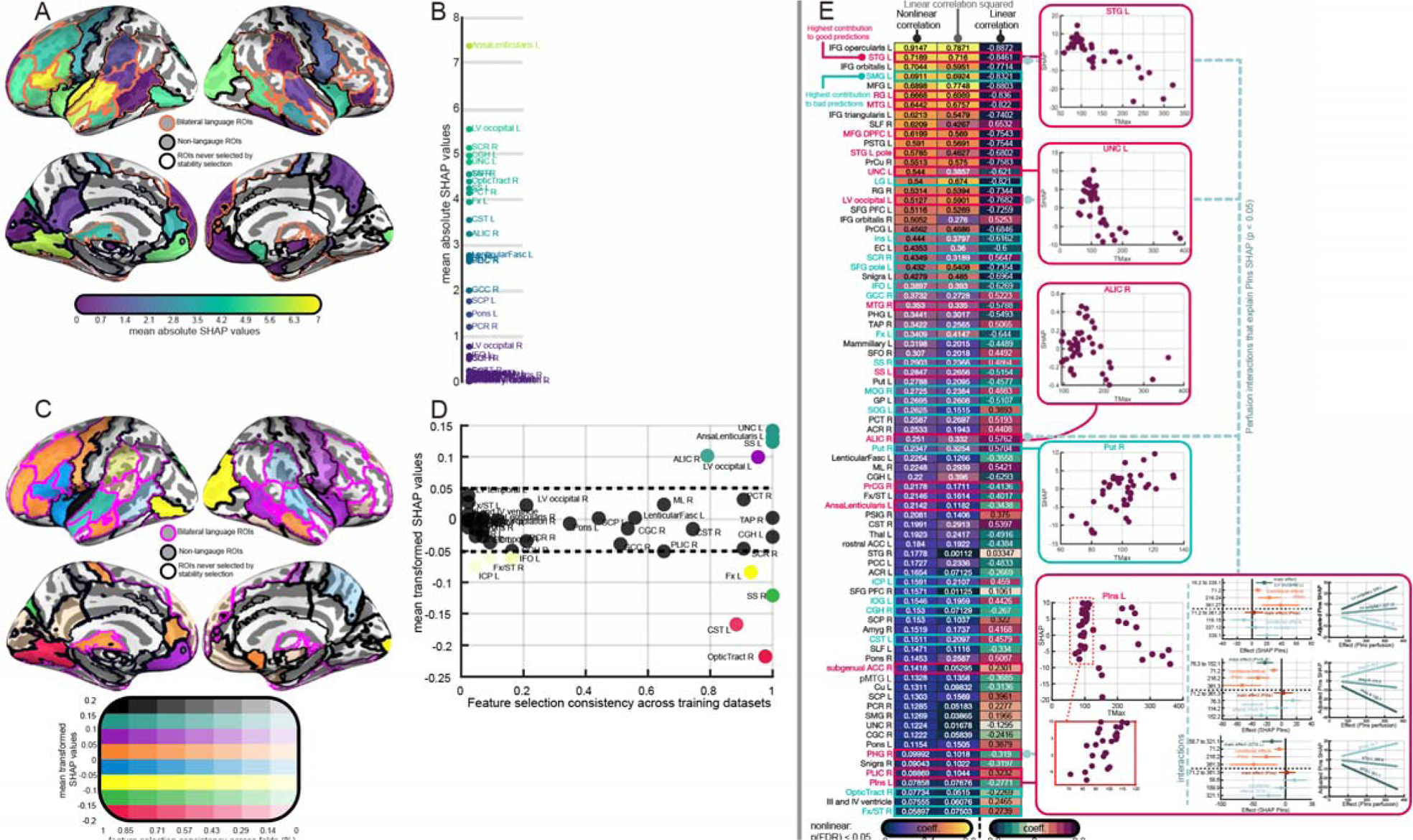
Explaining model predictions. In **panels A and B**, absolute SHAP values are collapsed across models from the outer CV loop, reflecting total impact of the feature on predictions. **Panel A** shows absolute feature contribution in cortical regions. Brighter region colors represent greater influence of the region on predictions. Regions without color were not selected by feature selection for any training dataset. Regions with orange outlines intersect with our set of theory-driven ROIs and regions with black outlines do not. **Panel B** shows absolute feature contribution for white matter regions. Brighter region colors represent greater influence of the region on predictions. In **panels C and D** SHAP values are also collapsed but first transformed so that above-zero values indicate the feature moves predictions from baseline (mean of all predictions) and in the direction of true values, whereas below-zero values indicate movement of prediction in the opposite direction of the true value. **Panel C** shows transformed SHAP values in cortical regions. Region colors represent SHAP bins and color transparency corresponds to the consistency with which a feature was selected across training datasets (i.e., including in the stable set during feature selection). Regions with pink outlines represent our theory-driven ROIs. **Panel D** shows SHAP values and feature selection consistency for white matter regions. Features that had a higher positive influence on model prediction are color coded according to the colormap in panel. **Panel E** shows the marginal dependence between perfusion in a feature and its contribution to predictions of higher impairment (i.e., raw SHAP values) across individuals. Here, positive SHAP values reflect predictions of higher BNT scores and negative values reflect predictions of lower scores. Linear (Pearson) and nonlinear (distance) correlation coefficients are presented as columns in a heatmap, with individual features representing rows. To facilitate comparison between the magnitude of linear and nonlinear association (i.e., the distance correlation is bound between 0 and 1), Pearson correlation coefficients are squared. A third column shows raw Pearson coefficients to demonstrate the direction of the relationship. Cells display coefficients, which also determine cell color. In the first two columns, brighter colors reflect stronger dependence. For the last column, red reflects stronger positive linear dependencies and blue reflects stronger negative linear dependencies. Only features that showed a significant linear or nonlinear relationship at p < 0.05 after FDR correction are shown (i.e., on distance correlation). Features with red labels fall into the top 10% of features ranked by transformed SHAP values, and features with blue labels fall into the bottom 10%. Example scatterplots between perfusion and SHAP are shown in red and blue boxes on the right. The scatterplot for the left posterior insula (lPIns) additionally shows perfusion interaction effects that explain SHAP for lPIns (i.e., the effect of changing one predictor as the other remains constant). The first column of plots shows main effects of predictors in darker colored dots and their conditional effects in lighter colored dots. Horizontal lines indicate 95% confidence intervals. The second column of plots shows interaction model predictions (in-sample) as a function of perfusion in lPIns. Although rALIC showed an interaction effect p < 0.05, it did not appear on interaction plots and was excluded (i.e., confidence intervals overlapped). Remaining effects were significant at p < 0.01.

**Table 2:** Features above 90^th^ percentile based on absolute SHAP values.

| Feature (ROI) | Absolute SHAP |
| --- | --- |
| AnsaLenticularis L | 7.39 |
| STG L | 7.01 |
| IFG triangularis L | 6.57 |
| Plns L | 6.56 |
| LG L | 5.87 |
| STG L pole | 5.73 |
| LV occipital L | 5.61 |
| Ins L | 5.24 |
| SCR R | 5.21 |
| MOG R | 5.18 |
| IFG opercularis L | 5.07 |
| RG L | 5.05 |
| CGH L | 5.03 |
| UNC L | 4.89 |
| PSMG R | 4.76 |
| IFG orbitalis L | 4.7 |
| TAP R | 4.63 |
| SS R | 4.63 |
| IOG L | 4.5 |

To understand which features contributed to more *successful* model predictions, we transformed raw SHAP values such that higher values always represented features moving the model from baseline and in the direction of the true value (see section 2.10 for more information about SHAP). That is, for any sample in the data, the sign of a SHAP value reflects whether the prediction for that sample is pushed higher (positive) or lower (negative) than the baseline prediction (i.e., mean prediction) and the magnitude reflects total feature impact on the model. However, baseline predictions may themselves be lower or higher than true values. Thus, the sign of a SHAP value does not embed information about whether the feature contribution was beneficial to predictions. We inverse transformed SHAP values such that *positive* SHAP always reflected the *positive* influence of a feature on model prediction.

Although transformed SHAP values presented a slightly different pattern of feature importance to absolute SHAP (see Figure 7C and D), many of the same features scored above the 90^th^ percentile on these transformed values (see Table 3), including: lPIns, lAns, lSTG, lSTGP, lOLV. Additional features of overlap between the two sets included the left uncinate fasciculus (lUNC), left rectal gyrus (lRG), and right posterior middle temporal gyrus (rpMTG). Transformed SHAP highlighted unique features as well, though many of them had relatively high absolute SHAP values that were not in the top 10 percent that we’ve focused on. These included: left sagittal striatum (including portions of inferior longitudinal fasciculus and inferior fronto-occipital fasciculus; lSS), right parahippocampal gyrus (rPHG), right anterior limb of the internal capsule (rALIC), right precentral gyrus (rPrCG), right insula (rIns), right middle temporal gyrus (rMTG), left middle frontal gyrus in dorsal prefrontal cortex (lMFG-DPFC), right subgenual anterior cingulate cortex (rsACC), the left lateral occipital ventricle (lLOV), left middle temporal gyrus (lMTG), and right inferior temporal gyrus (rITG). The highest transformed SHAP values—lPIns and lUNC—were notably higher than others, with lUNC marking the knee for sorted values.

**Table 3:**
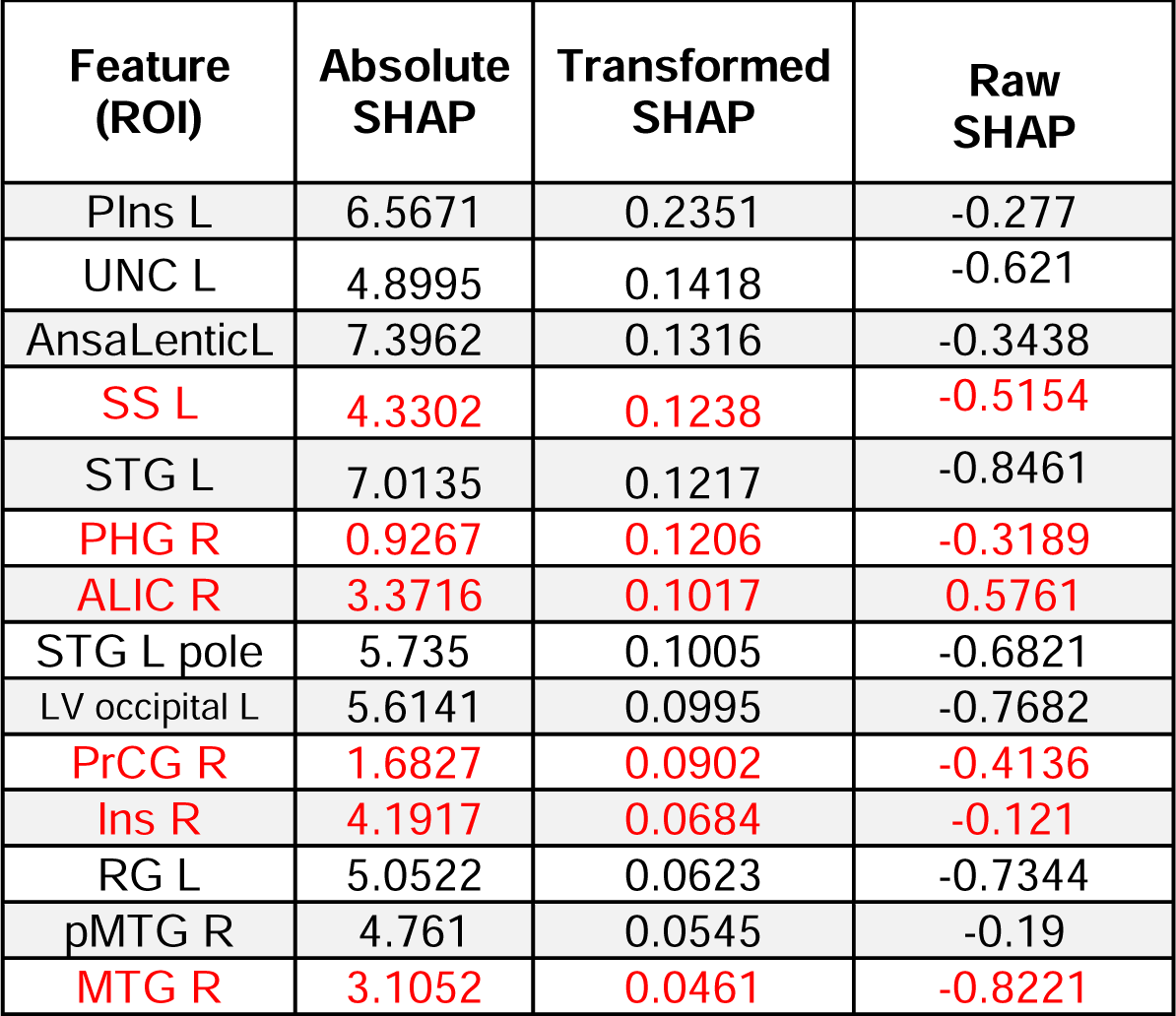
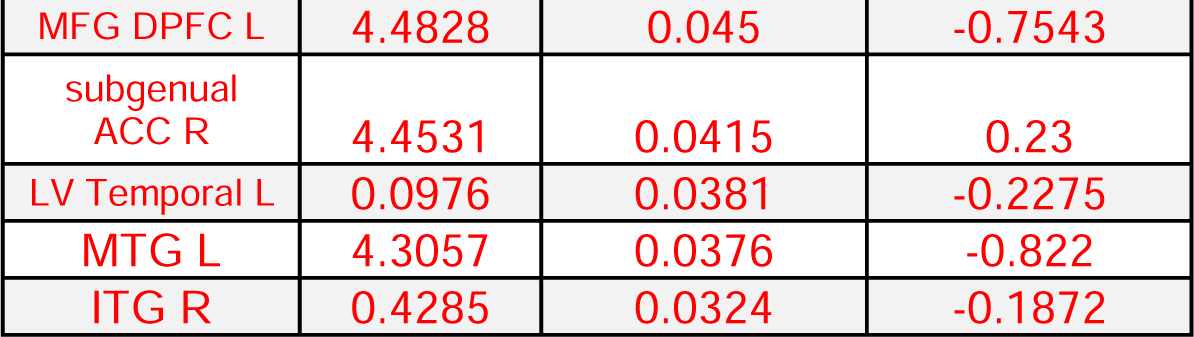
Features above 90^th^ percentile based on transformed SHAP values. Red text highlights features not in Table 2.

| Feature (ROI) | Absolute SHAP | Transformed SHAP | Raw SHAP |
| --- | --- | --- | --- |
| Plns L | 6.5671 | 0.2351 | -0.277 |
| UNC L | 4.8995 | 0.1418 | -0.621 |
| AnsaLenticL | 7.3962 | 0.1316 | -0.3438 |
| SS L | 4.3302 | 0.1238 | -0.5154 |
| STG L | 7.0135 | 0.1217 | -0.8461 |
| PHG R | 0.9267 | 0.1206 | -0.3189 |
| ALIC R | 3.3716 | 0.1017 | 0.5761 |
| STG L pole | 5.735 | 0.1005 | -0.6821 |
| LV occipital L | 5.6141 | 0.0995 | -0.7682 |
| PrCG R | 1.6827 | 0.0902 | -0.4136 |
| Ins R | 4.1917 | 0.0684 | -0.121 |
| RG L | 5.0522 | 0.0623 | -0.7344 |
| pMTG R | 4.761 | 0.0545 | -0.19 |
| MTG R | 3.1052 | 0.0461 | -0.8221 |
| MFG DPFC L | 4.4828 | 0.045 | -0.7543 |
| subgenual<br>ACC R | 4.4531 | 0.0415 | 0.23 |
| LV Temporal L | 0.0976 | 0.0381 | -0.2275 |
| MTG L | 4.3057 | 0.0376 | -0.822 |
| ITG R | 0.4285 | 0.0324 | -0.1872 |

A surprising number of influential features identified by absolute SHAP values drove model performance, on-average, in the *opposite* direction of true object naming scores (i.e., had negative mean transformed SHAP values; see Table 4). These represent difficult to model targets that can ostensibly improve future efforts at prediction. Counterproductive features falling below the 10^th^ percentile included: lLing, right sagittal striatum (rSS), right middle occipital gyrus (rMOG), left inferior occipital gyrus (lIOG), right superior corona radiata (rSCR), and lIns. Other features that were reduced in influence without driving models towards worse predictions included: lTri, left opercularis segment of the inferior frontal gyrus (lOperc), left orbitalis segment of the inferior frontal gyrus (lOrb), left cingulum (hippocampus; lCGH), and right tapetum (rT).

**Table 4:** Features below 10^th^ percentile based on transformed SHAP values. Red text highlights features from Table 2.

| Feature (ROI) | Absolute SHAP | Transformed SHAP | Raw SHAP |
| --- | --- | --- | --- |
| OpticTractR | 4.4818 | -0.217 | -0.222 |
| CST L | 3.6666 | -0.167 | 0.458 |
| LG L | 5.8742 | -0.164 | -0.82 |
| SS R | 4.636 | -0.12 | 0.486 |
| SOG L | 4.4347 | -0.102 | 0.39 |
| MOG R | 5.1803 | -0.084 | 0.488 |
| Fx L | 4.0445 | -0.083 | -0.643 |
| IOG L | 4.5079 | -0.081 | 0.442 |
| SMG L | 1.2154 | -0.076 | -0.832 |
| ICP L | 0.23 | -0.073 | 0.459 |
| SFG pole L | 2.1285 | -0.07 | -0.735 |
| Put R | 4.2242 | -0.067 | -0.458 |
| Fx/ST R | 0.4196 | -0.061 | -0.401 |
| IFO L | 0.7535 | -0.061 | -0.627 |
| PLIC R | 2.7897 | -0.05 | 0.323 |
| CGH R | 0.6797 | -0.049 | -0.629 |
| SCR R | 5.218 | -0.046 | 0.565 |
| Ins L | 5.2484 | -0.04 | -0.616 |
| GCC R | 2.1615 | -0.038 | 0.522 |

Feature selection consistency across training datasets (i.e., the proportion of times a feature was included in the stable set during stability selection) is also presented in Figure 7C and highlights the extent to which a feature was beneficial or detrimental to specific configurations of patients in training datasets. Most of the influential features discussed above were selected in *<u>all</u>* of the training datasets, including the highly influential lPIns and lUNC, but also lAns, lSS, lSTG, lSTGP, lMFG-DPFC, lMTG and rITG. However, other influential features were not as frequently selected, reflecting stability selection adapting to unique training datasets. For example, the following features appeared in less than 80% of models: rMTG (79%), rPrCG (47%), and rPHG (17%). Features that pulled predictions away from true values also differed by consistency, but ultimately, consistency was not associated with transformed SHAP values based on the Pearson correlation coefficient, r(89) = 0.08, p = 0.4 and there was no mean difference in feature consistency between the top 10% (M=0.79, SD=0.34) and the bottom 10% (M=0.7, SD=0.36) of features based on a two sample t-test, t(36) = 0.9, p = 0.36.

We also captured the statistical association between the perfusion of each feature and the models’ tendency to associate that feature with greater language impairment across individuals. Our transformation of SHAP values purposefully supplanted the meaning of the sign such that negative values no longer represented a feature driving model prediction towards lower BNT scores. Consequently, we looked at linear (i.e., Pearson) and nonlinear (i.e., distance correlation; Szekely, Rizzo, Bakirov, 2007) correlations between perfusion and raw SHAP values across individuals. For completeness, we show all features that exhibited a significant linear or nonlinear relationship after FDR correction (p < 0.05; Figure 7E). Features we previously identified to be associated with more successful model predictions had SHAP values among the strongest related to perfusion, including lSTG, lRG, lMTG, lMFG-DPFC, lSTGP, lUNC, and lLOV. Only 3 of the 19 top features contributing to successful model predictions did not exhibit a significant relationship between SHAP and perfusion (i.e., rIns, lLTV, and rITG). However, our SHAP values reflected feature contributions to interaction effects that would be missed by marginal conditional dependencies. Many of the features that contributed to poorer model predictions also showed significant relationships between SHAP and perfusion, although curiously many of them showed a counterintuitive linear trend. For example, 11 of the 19 features contributing to worse predictions showed a positive linear association whereby longer delays as represented by higher Tmax were associated with predictions of higher BNT scores. Although we observed some unidentified positive linear relationships directly between Tmax and BNT scores in Figure 2, the fact that only 3 of 16 features contributing to better model predictions showed the same trend (i.e., rALIC, lAns, and rsACC) suggests that the phenomenon of higher Tmax contributing to better language performance is typically unproductive for predicting BNT scores. Instead, most of these features exhibited an inverse relationship such that higher Tmax was linearly associated with greater language impairment.

Features important for making better model predictions that showed significant, yet low statistical associations between SHAP and perfusion likely derived their importance from interaction effects. As there were not many of these features, we focused on the weakest statistical association, which also happened to be the feature that had the largest positive influence on predictions—the lPIns. Using an interaction-only robust linear regression model with the bisquare function (tuning constant 4.685) to downweigh the role of outliers, we found that 3 of the 9 features with the highest contribution to more successful models had perfusion that significantly interacted with perfusion in the lPIns to explain its association with SHAP (p < 0.05; Figure 7E, bottom right box; see supplemental section 2.9, Table S7, Figure S17 for more details including scatterplots). Interaction plots highlighted that when Tmax in either lSTG or rPHG was low, the model interpreted higher Tmax in lPIns to translate into better naming performance (i.e., higher lPIns SHAP). However, when Tmax in these regions was moderate the effect weakened, and when Tmax was high, it reversed, such that higher Tmax in lPIns predicted lower naming performance. Thus, up to a certain point, lower Tmax in lSTG or rPHG was associated with a beneficial effect of higher Tmax in lPIns on language. A reverse pattern was found for the interaction between lPIns and lLOV, whereby only when Tmax in lLOV was moderate or high was there a positive effect of higher Tmax in lPIns.

### 3.7 StabSel: a toolbox for stability selection

The pipeline we have implemented for stability selection can be leveraged for other studies using the stabSel MATLAB toolbox that we have made publicly available. Although the present work focused on wrapping stability selection around elastic net, our package supports stability selection using many of the external feature selection methods that we have tested, including various correlation-based filters, RreliefF, NCA, lasso, and elastic net. We additionally provide our own implementations of adaptive lasso and elastic net, which have the oracle property (for implementation, see: Zou, 2006; Zou & Zhang, 2009). Further, the package implements PLS-VIP as a feature selection approach, permitting multi-task learning. We provide the option to turn any of the implemented selection techniques into a rudimentary multi-task learning method by concatenating response variables. Our package also permits implementation of out-of-bag error for RF models, automatic relevance determination using GPR, robust linear regression, ridge regression, the F-test, t-test, Wilcoxon test, Kullback-Leibler divergence, and maximum redundance maximum relevance algorithm for ranking features for stability selection. Some of these approaches can be used for classification and regression problems (e.g., RF), while others are only available for regression (e.g., elastic net) or classification (e.g., t-test).

Given the extensive options available for ranking and selecting features, we provide detailed documentation with the package. This includes multiple comprehensive tutorials with interactive ‘live’ code that describe how error control is implemented in stability selection, as well as details about algorithm implementations (Figure 8). Tutorials involve analysis of public ‘toy data’. In one tall dataset, multiple stability selection approaches are implemented to improve simple models fit to audio data in order to predict the year in which a song was produced (Bertin-Mahieux, 2021). In another publicly available dataset, discourse measures are used to predict individuals at-risk for mild cognitive impairment based on another behavioral assessment (Wilson et al., 2023). Both tutorials go beyond stability selection, showcasing how to properly set up cross-validation schemes, demonstrating the incredibly misleading results that can be attained by failing to prevent data leakage, showing how to fit many different models available in MATLAB’s machine learning toolbox (e.g., decision trees, random forests, support vector machines, etc), and providing code for tracking the results of model tuning live so that users can monitor the consistency of solutions (Figure 8). Finally, as we anticipate this toolbox to be helpful for analysis of neuroimaging data, code is also provided for loading preprocessed imaging data in standard formats (i.e., NIFTI).

**Figure 8.**
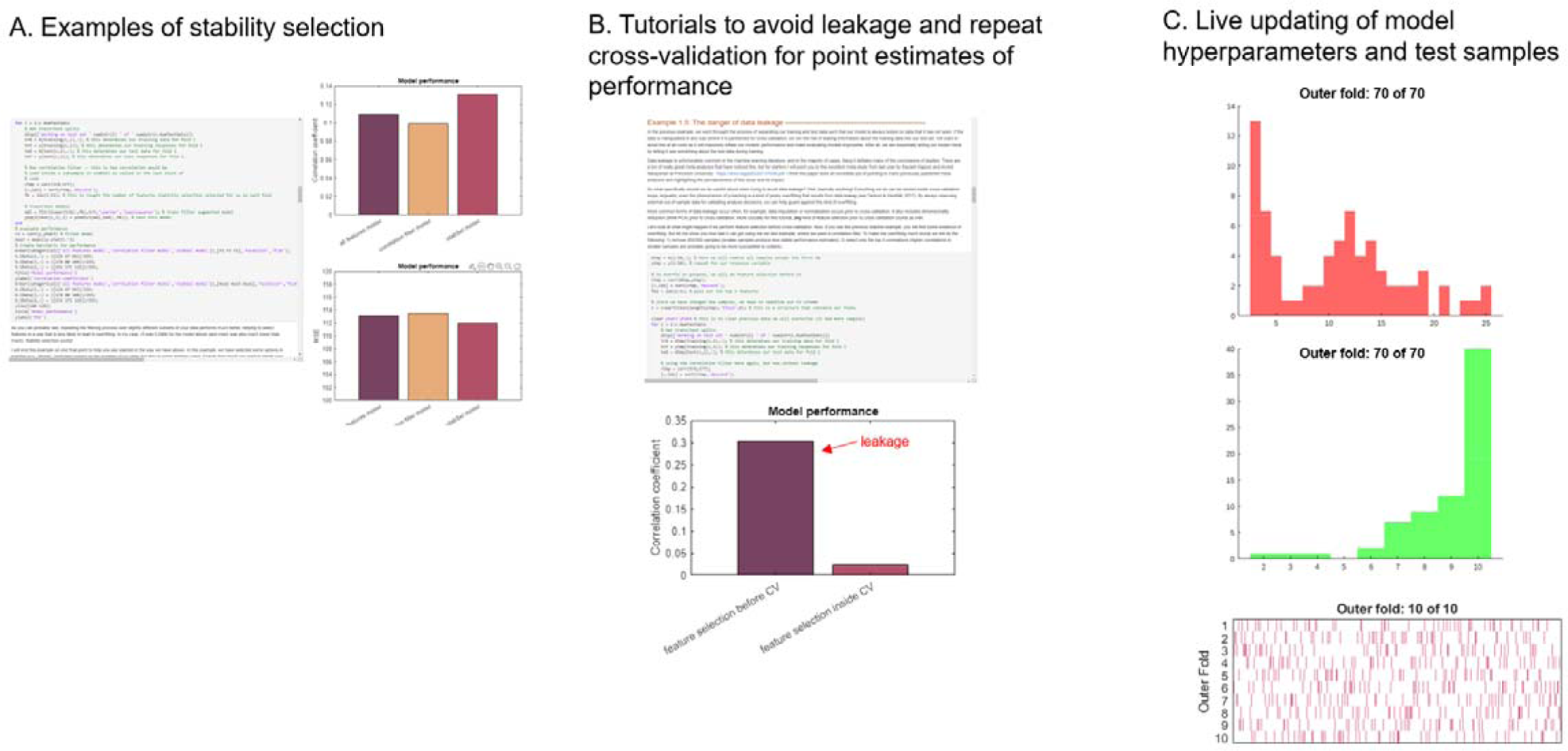
Additional material packaged with the stabSel toolbox for performing stability selection. **Panel A** showcases a small portion of one tutorial that implements one of many examples of different stability selection approaches applied to toy data. The editor displays interactive code that updates bar charts on the right. **Panel B** shows a different portion of the same tutorial demonstrating the impact of data leakage. Code is provided that can be readily adapted to establish proper repeated and/or nested cross-validation schemes. **Panel C** presents an example of code output that updates the results of hyperparameter tuning and data sampling for testing in real time.

## 4. Discussion

### 4.1 Towards larger whole brain perfusion datasets in stroke

The success of machine learning in fields like computer vision has galvanized an explosion of studies applying these techniques to neuroimaging data (e.g., Davatzikos, 2019; Mateos-Perez et al., 2018; Nielsen et al., 2020; Arbabshiriani et al., 2017; Ashley, 2016; Bonkhoff & Grefkes, 2022; Bzdok et al., 2018; 2020; 2021; Bzdok & Ioannidis, 2019; Kosorok & Laber, 2019; Poldrack et al., 2019; Shmueli, 2010; Mwangi et al., 2014). The most impressive machine learning results have emerged from datasets that are both less complex, more homogenous, and orders of magnitude larger than typical clinical datasets (e.g., Hulsen et al., 2019; Varoquaux & Cheplygina, 2022). For example, clinical data are more difficult to organize and share as it contains private health information and tends to require a large amount of preprocessing. Calls that current clinical neuroimaging work with machine learning is “data starved” have been numerous (e.g., Hulsen et al., 2019; Kohli et al., 2017; Poldrack et al., 2019; Shah et al., 2019; Varoquaux & Cheplygina, 2022). Indeed, recent estimates suggest clinical neuroimaging studies utilizing functional scans tend to have a median of just ∼24 participants (Szucs & Ioannidis, 2020).

The smaller clinical datasets that are currently available make modeling more challenging by limiting power and increasing risk of overfitting (e.g., Raudys & Jain, 1991). When a limited amount of data is available for testing and validating models, data partitions might exhibit reduced signal, an increased presence of influential outliers, and represent a less comprehensive sampling of the complete distribution from which the sample was originally drawn. This can translate into suboptimal choices of model complexity during tuning and substantially more variable (typically optimistic) estimates of model performance on test sets, reflecting overfitting to noise (e.g., Flint et al., 2021; Lever et al., 2016; Varoquaux, 2018; Varoquaux & Cheplygina, 2022). While there is no substitute for large high-quality datasets, a promising approach for addressing the current issue plaguing machine learning applications to clinical neuroimaging data is the acceleration of large-scale datasets that capitalize on data already collected during routine care. While this type of data generally lags behind state-of-the-art research acquisition protocols (Chalela et al., 2007), the reduced signal to noise ratio at the level of individual scans may be an acceptable trade-off for better-trained models with more realistic estimates of model performance. For example, recent work has suggested overoptimistic estimates of model performance in studies with smaller sample sizes are primarily caused by small test partitions (Varoquaux & Cheplygina, 2022). Despite the understanding that assessments of model performance during cross-validation can be imprecise and optimistic (Varoquaux et al., 2017), machine learning studies in neuroimaging continue to underutilize practices such as permutation testing to help address these concerns (Cearns, Hahn & Baune, 2019; Ojala & Garriga, 2010; Varoquaux et al., 2017).

In the current study, we have demonstrated the feasibility of amassing large scale functional CT perfusion datasets in acute stroke by describing a straightforward approach for converting proprietary perfusion images output by industry software that is commonly used in the clinic into scalar data. Specifically, we have demonstrated that this data can be preprocessed using pipelines common to research studies, permitting the pooling of whole brain CT perfusion data across groups of patients. Going further, we showed that this preprocessed whole brain perfusion data can be a useful predictor of stroke outcome.

The majority of studies leveraging CT perfusion data collected in clinical settings have focused on gross measures related to the lesion, both for the technical reasons outlined above and because perilesional information has been shown to improve selection of patients for interventions (Albers et al., 2018; 2021; Nougueira et al., 2018; Jovin et al., 2015; Goyal et al., 2015; Martins et al., 2020). Using a relatively limited sample size, we established that whole brain CT perfusion data can predict the severity of naming impairment in acute stroke, significantly surpassing the performance of a naïve guessing model as confirmed by permutation analysis. Strikingly, these predictions were achieved by models that assigned a moderate degree of importance to right hemisphere regions not directly damaged by the stroke, clearly indicating information outside the lesion facilitated prediction.

The emphasis of the model that we have developed on regions outside the direct area of injury contributes to a growing appreciation that extralesional tissue integrity represents a unique source of variance in language outcomes. A wealth of evidence shows that coarse information about lesion location and size explains a comparatively large amount of variance in acute and chronic stroke outcomes, including language (Goldenberg & Spatt, 1994; Johnson et al., 2022). However, stroke induces widespread changes in brain morphology and function (e.g., neuroinflammation, Wallerian degeneration; Hinman et al., 2014; Jayaraj et al., 2019; Shen et al., 2021). Indeed, a number of recent studies have reported functional and structural brain changes distal to the lesion that are related to language outcomes (e.g., Koh et al., 2021; Catani & Mesulam, 2008; Kristinsson et al., 2021; Yourganov et al., 2016). Further, studies have associated stroke outcomes with the accumulation of microvascular injuries (Varkanitsa et al., 2020; Wright et al., 2018) and shown that accelerated age-related patterns of atrophy across the brain are associated with increased risk of mild cognitive impairment post-stroke (Aarmodt et al., 2023), and generally poorer stroke outcomes (Liew et al., 2023).

Our work broadly underscores the utility of granular information from CT perfusion for outcome prediction. This type of information has been overlooked in previous work evaluating whether gross CT perfusion features around the lesion and perilesional area (e.g., number of voxels in the penumbra and core lesion as assigned by difficult to interrogate industry software) adds unique predictive information relative to other imaging modalities. For example, Brugnara and colleagues (2020) trained gradient boosted decision trees on multimodal imaging features to predict modified Rankin Scale scores, finding that adding CT perfusion features to the model did not improve prediction. However, a more recent study using a similar set of imaging data ranked CT perfusion features among the most important for prediction (Jiang et al., 2021). Another recent study reported the same pattern (Jabal et al., 2022; see also Bivard et al., 2017 for earlier work with similar results). The models in these more recent studies demonstrated comparable or better performance and implemented a more efficient algorithm for boosting decision trees (i.e., xgboost), which may have permitted the discovery of a more optimal set of hyperparameters that led to a relatively greater weighting of perfusion measures in the model. More complex deep learning approaches have also indicated that inclusion of CT perfusion data to other clinical features improves prediction of stroke severity (Bacchi et al., 2020) and our work aligns with this more holistic approach for evaluating the contribution of perfusion.

### 4.2 Stable features enhance predictive models

The increasing prevalence of wide and tall datasets is challenging conventional modes of analysis that rely on classical statistical methods, most of which were tailored for situations we would now characterize as having small sample sizes and low dimensionality (Bzdok, Nichols, Smith, 2019). Here, we presented a case where classical statistical modeling using a simple multivariable linear regression of whole brain imaging data failed at both inference and prediction, even when the proportion of samples to features was tamed by collapsing individual voxels into large regions within an atlas and only using regions with published evidence of association to naming impairments to improve model estimation (i.e., theory-driven feature selection). Notably, our sample size was roughly twice as large as the typical clinical neuroimaging study with functional data (Szucs & Ioannidis, 2020). We stress this models’ failure for two reasons. First, this finding underscores that classical inferential analysis, although perhaps more suited to small sample sizes, can be incompatible with a data-driven perspective. Although engaging in pre-modeling feature selection may have improved the model (e.g., by investigating intercorrelated features), this strategy would risk inflating significance and compromise the generalizability of the resulting ‘explanation’. The primary objective of inference is to draw conclusions about the population based on the observed data, and a purely data-driven analysis that incorporates all relevant features ensures a more unbiased exploration of relationships. Our second reason for drawing attention to this models’ failure is to emphasize that simple models can fail to generalize and that increasing model complexity within reason, which we do in subsequent analyses, can enable predictive models in relatively small clinical neuroimaging datasets.

Increasing model complexity in clinical neuroimaging datasets is challenging because wide datasets are the norm, where hundreds of thousands of features or voxels are collected in each individual. Consequently, complex models are more likely to take advantage of noise or data idiosyncrasies that do not generalize. In a demonstration of the concept that models promoting sparsity (i.e., feature selection) trade-off stability (Xu et al., 2011), we found that data-driven feature selection using methods that embedded feature selection during model estimation produced predictions of naming impairments that performed worse than a naïve guessing model. Indeed, we found that a much more successful strategy for building predictive models in limited sample sizes was to apply these same modeling techniques to regions preselected on theoretical grounds. Thus, a very simple manipulation of the model using results from prior studies was effective at constraining the feature space to the point that regularization techniques could be exercised to strike a good balance between model complexity and stability. For example, techniques such as lasso, ridge, and elastic net were all able to estimate feature coefficients that were more effective at predicting language impairments.

However, we discovered that models tended to perform best when stable features were identified and refined in a purely data-driven way through stability selection. In our dataset, this approach resulted in predictions that outperformed a naïve model significantly better than chance, which the alternative theory-driven selection approach did not. Critically, we found that stability selection boosted model performance across a wide range of machine learning and simple regression algorithms. Furthermore, it tended to perform substantially better than other external feature selection methods, demonstrating the advantage of prioritizing feature stability over generalizability. Feature stability is more advantageous for feature selection in small sample high dimensional datasets because typical feature selection approaches amount to building preliminary models that suffer from the same general trade-off between model sparsity and stability. By evaluating the consistency with which an algorithm selected features and ignoring out-of-sample errors, stability selection improved model estimation. Consistent with this interpretation, all feature selection based on out-of-sample error tended to perform poorly in our dataset (including dimensionality reduction) and the only approach that was competitive with stability selection was a genetic algorithm that similarly evaluated feature consistency during the selection process but did not rely on this exclusively. These findings support other results in relatively small-sample imaging datasets that suggest that discovering and incorporating stable features into models acts as a stabilizing factor, bolstering their predictive performance (Rondina et al., 2014; Jollans et al., 2019). Here we have built on this prior work, demonstrating a similar result in a regression problem (c.f., Rondina et al., 2014) with a comparatively smaller sample size, and while formalizing the identification of stable features with stability selection using an elastic net (c.f., Jollans et al., 2019).

One point of difference between our approach and others is that we report some of the lowest model errors when stability selection is ‘supervised’. For example, using LASSO to estimate the model after stability selection resulted in better predictions than using ridge or elastic net. This strongly suggests that stability selection benefitted from a more aggressive secondary filter for features. While theory-driven feature selection benefitted from this type of secondary feature selection step as well, the benefits were greater for stability selection with elastic net. Given these results, we suggest stability selection outperformed theory-driven feature selection by identifying unanticipated features that predict language impairment. However, it was also less selective, identifying stable but not highly predictive features. While the tradeoff between stability and prediction has been pointed out by previous authors (Tian & Zalesky, 2021; Varoquaux et al., 2017), here we found that stability analysis is highly effective for *enabling* predictive modeling in clinical neuroimaging datasets, which tend to have relatively small samples. Future work using stability selection to stabilize models should consider the additional component of refining the stable set using out-of-sample data. We emphasize that while stability selection provides error control, it does not evaluate the generalizability of features directly, and the optimal implementation of error control remains a topic of interest (e.g., Hofner et al., 2015; Ahmed et al., 2011; Dai et al., 2022). For example, while the original authors of stability selection suggest that subsampling a larger proportion of the data or manipulating the threshold for defining the stable set should not have a major impact on feature significance (Meinshausen & Buhlmann, 2010), this is likely not the case (Shah & Samworth, 2013). Moreover, data assumptions in the original stability selection framework impose highly conservative bounds and may be relaxed (Ahmed et al., 2011; Shah & Samworth, 2013). Given the stringency with which the upper bound on false discoveries is established, stability selection may work best in small samples as a preliminary model building step that identifies the most plausibly predictive features and may be more useful as a standalone tool for feature discovery in larger sample sizes.

One of the exciting aspects of the stability selection approach that we have employed in this study is that it is highly flexible. For example, as an ensemble feature selection method it may be used to fuse multiple complementary feature selection approaches to identify more unique subsets of features than we were able to investigate here. While recent packages have been implemented for stability selection in R (Hofner, 2021) and python (https://github.com/scikit-learn-contrib/stability-selection), much of the neuroimaging community relies on MATLAB for preprocessing and analysis (Ashburner, 2012). We have publicly published a MATLAB toolbox for stability selection that can be used to replicate the approach that we have taken here (i.e., elastic net), as well as to implement 14 other classification and regression algorithms that leverage MATLAB’s machine learning toolbox. We additionally see this package as an opportunity to highlight to researchers the dangers of data leakage that have become problematically common in neuroimaging studies (Poulin et al., 2019; Eitel et al., 2021; Kambeitz et al., 2015; Pulini et al., 2019; Whelan & Garavan, 2014; Mateos-Perez et al., 2018; Yagis et al., 2021; Kapoor & Narayanan, 2022; Rosenblatt et al., 2023; 2023; Poldrack et al., 2019), and package our toolbox with a variety of tutorials for implementing appropriate cross-validation with feature selection. We anticipate this package will remain useful even as clinical neuroimaging datasets grow in size as it is well-established that even in datasets with a limited number of features or more equal ratio of features to samples, removing redundant or noisy features can improve model estimation and performance by reducing the amount of noise that is available for the model to overfit (e.g., Bzdok et al., 2018; Hawkins, 2004; Heinze et al., 2018).

### 4.3 Brain regions supporting naming

Language production involves a series of complex psycholinguistic processes that result from the orchestrated interplay of activity within large-scale networks (Hickok & Poeppel, 2007; Indefrey & Levelt, 2004; Price, 2012). Commonly assessed using picture naming, gross language impairment as a result of stroke has been associated with a frontotemporal network (Stark et al., 2019; Tochadse et al., 2018; Dell et al., 2013; Na et al., 2022). Here, we found that perfusion in left inferior frontal gyrus (pars triangularis), left posterior insula, and left superior temporal gyrus had the largest impact on model predictions of any regions in cortex. Upon closer inspection, we found that pars triangularis had a strong but deleterious effect on models, pushing their predictions away from true values. Further, the left posterior insula emerged as having a slightly higher overall positive effect on model predictions than the left superior temporal gyrus. While some studies have found an association between damage in the inferior frontal gyrus and naming impairment, recent work suggests that this effect is not consistent across studies, unlike associations in temporal and inferior parietal cortex (Piai & Eikelboom, 2023). Our results further show that despite exhibiting a significant association with language impairment, perfusion in pars triangularis does not consistently predict impairment.

Naming studies typically involve objects, however, there is evidence that object and action naming dissociate (e.g., Breining et al., 2022; Hillis et al., 2002; Thompson et al., 2012) and several studies have implicated the insula in action naming (e.g., Akinina et al., 2019; Breining et al., 2022; Luzzatti et al., 2006). Posterior insula represented the only feature in our dataset with markedly high model influence that exerted its effect through interaction with other regions. One of the most interesting patterns that emerged from our data is that our predictive model downweighed significant positive relationships between delayed perfusion (i.e., higher Tmax or time-to-maximum) and language impairment. Only a single instance of this counterintuitive relationship aided prediction—perfusion in the left posterior insula showed a beneficial impact of longer Tmax on language performance, but only when Tmax was concurrently low in left superior temporal gyrus or right parahippocampal gyrus. Thus, this beneficial effect of longer delays disappears when there is dysfunction in left superior temporal gyrus or right parahippocampal gyrus, likely due to direct damage (i.e., left hemisphere) or widespread mechanisms such as diaschisis (i.e., right hemisphere). Ultimately, this suggests that the left superior temporal gyrus acts as the strongest catalyst for accurate prediction of naming impairment. Together, these findings are not inconsistent with prior work in acute stroke that has used similar regression techniques (i.e., lasso but without stability selection and not for out-of-sample prediction) to show that damage to the superior temporal gyrus is the strongest predictor of object naming (Breining et al., 2022).

Outside of cortex, we also observed a strong effect of the left uncinate fasciculus on successful model predictions. Damage to this limbic tract has been previously implicated in object naming (Pisoni et al., 2018) and it connects the temporal lobe in the vicinity of anterior parahippocampal gyrus with orbitofrontal cortex (Olson et al., 2015). Other medial regions also showed relatively high influence on model predictions, including left ansa lenticularis and left sagittal stratum. The impact of left sagittal stratum is consistent with prior work showing an association between poorer language recovery post-stroke and damage to this region (Sul et al., 2019). The sagittal stratum is a broad corticosubcortical white matter bundle that connects parietal, occipital, cingulate, and temporal regions to the thalamus (as well as brainstem structures). Recent meta-analysis has indicated that damage to the basal ganglia, which includes the ansa lenticularis, as a result of stroke or other pathologies is associated with naming impairment relative to controls (Camerino et al., 2022).

Notably, our models integrated information from less highly influential regions that spanned the entire brain. Many regions from both hemispheres had a small to moderate impact on the model and were not as closely connected to previous studies on language impairments in stroke. For example, the bilateral precentral gyri but especially right precentral gyrus, contributed to successful prediction. Evidence from neuroimaging, electrocorticography and direct stimulation suggests that multiple regions within the precentral gyrus are critical for speech and embedded in larger networks contributing to speech (Hickok et al., 2023). Perfusion in these regions may be affected by downstream effects of more conventional damage to temporal or parietal targets of the language system. Other regions of moderate influence included the right posterior middle temporal gyrus. The middle temporal gyrus sits at an intersection of many white matter tracts linked to different aspects of language, potentially explaining association between damage to this region and a variety of language deficits (Griffis et al., 2017; Turken & Dronkers, 2011). In many cases, this set of less influential regions highlighted the contribution of intact right hemisphere homologues of regions critical to language to successful prediction of deficits, confirming the role of these right hemisphere regions in supporting language and indicating that our model capitalized on dysfunction within large-scale language. Finally, we point out that the brain structures discussed were selected consistently by stability selection across all of the training datasets in our cross-validation scheme. Consequently, these effects are less likely to be driven by outliers or idiosyncrasies in the data, though we would also point out that we found evidence that models which assigned more importance to less frequently selected features tended to have smaller errors, suggesting that such features were often meaningful as well and that ‘supervised’ stability selection effectively adapted to unique training datasets.

### 4.4 Limitations

A challenge to our investigation into algorithmic differences in model performance is that we may have encountered false positives or overfitting as a result of increasing the effective degrees of freedom (Hosseini et al., 2020). Further, small sample sizes can inflate model performance (Varoquaux et al., 2017). We acknowledge these criticisms but would counter in the following ways. First, we don’t see specific differences between algorithms as paramount to our goals of demonstrating that: i) evaluating feature stability tends to enhance predictive models, and ii) clinical perfusion imaging can be used to predict language impairments successfully. We found that evaluating feature stability improved models consistently, irrespective of the specific algorithm that was tested. This pattern is conceptually supported by the fact that feature selection in machine learning is demonstrably unstable, particularly when model estimation prioritizes sparsity. Moreover, underestimation of imprecision in small sample sizes is particularly problematic for hypothesis testing and one way to mitigate this problem is permutation testing (Varoquaux et al., 2018). In this way, we have shown that stability selection produced models that performed better than chance, whereas more conventional qualitative selection based on prior work (i.e., theory-driven) did not. We don’t contend that other researchers may have had more success with a particular feature selection method, or even with qualitative feature selection. Indeed, our own results show that qualitative feature selection is preferential to purely ‘data-driven’ analysis using all features. Instead, we would argue this approach is difficult to formalize (Yarkoni, 2022) and evaluation of feature stability offers a robust alternative.

## Code availability

Virtually all models were trained, tested and tuned using in-house code relying on MATLAB’s machine learning toolbox functions (The MathWorks Inc, 2021). The one exception to this is the genetic algorithm, which was implemented using the MATLAB regression toolbox (Consonni, 2021). The core code we have used leverages the stability selection toolbox developed for this work and which can be found alongside tutorials for performing the same or similar analyses at: https://github.com/alexteghipco/StabilitySelection. Any other code will be made available upon reasonable request to the corresponding author.

## Data availability

Anonymized data will be made available upon reasonable request to the authors, subject by the Johns Hopkins University School of Medicine Institutional Review Board resulting in a formal data-sharing agreement.

## Supplemental material

### 1. Methods

#### 1.1 Cross-validation

We elected to use leave-one-out cross-validation (LOOCV) to estimate model performance in order to maximize the amount of training data available for the models while also minimizing the variability in estimates of model performance. For instance, in classification tasks, LOOCV has been reported to better estimate prediction error than *k*-fold cross-validation with small *k* and split-sampling when the data is characterized by small sample sizes and many features (Molinaro et al., 2005). The cross-validation scheme was repeated as data partitioning ‘noise’ can have substantial impact on estimates of model performance (e.g., Krstajic et al., 2014).

#### 1.2 Approach to hyperparameter tuning

As described in the main text, we used grid search for tuning most models, including feature selection when it involved hyperparameter tuning. To accommodate the larger set of hyperparameters in more complex machine learning algorithms used for predictive modeling (i.e., support vector regression, random forest, gaussian process regression), a more efficient random search was performed.

#### 1.3 Exceptions to normalization during feature selection

Occasionally, feature selection did not involve normalization, however, model building always did. For example, normalization has no influence on correlation-based feature selection methods. In addition, we elected not to perform normalization while using the RreliefF algorithm and neighborhood components analysis.

#### 1.4 Correlation-based filters for feature selection

In using correlation-based filters, one of our strategies entailed selecting the top 30 features based on their correlation with naming scores. The arbitrary value of 30 was chosen as it roughly approximates the number of theory-driven ROIs we expect to be predictive of object naming (i.e., of the total 44 ROIs, we expect only a small portion of the 22 right hemisphere ROIs to contribute to the model but have no theoretical grounds on which to exclude them). In other analyses, we tuned the top *n* features to select using an inner cross-validation loop. The *n*-sized filter windows that were evaluated were: 5, 10, 15, 20, 25, 30, 35, 40, 50, 60, and 70. As another strategy we retained all features with significant correlations at p < 0.05 but we note that during testing, we also used p(FDR) < 0.05, p < 0.01 and p < 0.001 cutoffs and performance was comparable across all of these thresholds. We present the results for p < 0.05 in the main text because the number of surviving features better matches other feature selection methods that were more successful.

Note, several correlation filters retained the 30 highest correlated features with naming scores. This number of features was arbitrarily chosen as the filter window because: i) it roughly aligns with the number of significant correlations between features and response values that we observed in Figure 1, and ii) it roughly corresponds to the number of regions we might expect to be involved in object naming when extending our preselected left hemisphere ROIs to right hemisphere homologues (i.e., assuming all left ROIs support object naming and roughly half or fewer of the right hemisphere homologues do as well).

#### 1.5 Stepwise and sequential methods for feature selection

Stepwise regression methods were tested using BIC, AIC, adjusted R^2^ and significant difference in sum of squared errors on an *F*-test (i.e., p < 0.05). Both BIC and AIC were included as AIC seeks to identify the best approximating model by minimizing Kullback-Leibler divergence between estimates and the true distribution, while BIC seeks to identify the true model from amongst a set of alternatives by optimizing posterior model probability. For sequential feature selection, we used mean absolute error (MAE) on out-of-sample data as the criterion. MAE was chosen instead of sum of squared errors because it is the basis of the accuracy percent measure we use to evaluate model performance overall.

#### 1.6. L1 and L2-norm penalties for feature selection

In smaller datasets, tuning an additional parameter may encourage overfitting and produce a model inferior at generalizing to unseen data. For elastic net, we used two different sets of a parameters for feature selection and model building. The feature selection range of a values is presented in the main text (section 2.8.2). When elastic net was not used strictly for feature selection, we reduced the a range to 6 linearly spaced values between 0.5 and 1 (i.e., excluding ridge regression entirely).

#### 1.7 Neighborhood component analysis

For NCA, we chose to tune the ε value for the loss function. By using an ε-insensitive loss function, NCA can be made more robust to outliers. NCA shrinks feature weights to zero, but in practice, we found it to sometimes shrink weights to similar near-zero values. As NCA feature weight magnitudes varied across training datasets, we could not apply a single weight threshold to successfully retain features selected by NCA. Instead, we automatically identified selected features using the elbow of sorted feature weights. The elbow was programmatically identified by finding the maximal Euclidean distance between sorted feature weights and a line connecting the largest and smallest feature weights. NCA is closely related to the k-nearest neighbor algorithm. Although we do not present the results of combining NCA-based feature selection with k-nearest neighbor regression in the main text, we note that during testing, we evaluated this combination and found that it did not produce predictions as good as lasso and elastic net.

#### 1.8 Ensemble feature selection with stability selection

Empirical work has demonstrated that feature selection methods *tend* to fail when the number of features vastly exceeds the number of samples, though we note that our dataset does not have the kind of imbalance more typical of the genetic studies that have often attracted this kind of experimental work (e.g., Kuncheva et al., 2020; Yang et al., 2006). We used stability selection in an effort to identify more reliable features for modeling.

The current study implemented stability selection by developing a comprehensive MATLAB package that supports this framework in the context of 14 different feature selection methods (https://github.com/alexteghipco/StabilitySelection). The package supports data resampling using either subsamples or complementary bootstraps and can implement outlier detection within the resampling scheme (e.g., MAD, one-class support vector regression, isolation forests). To the extent of our knowledge no other MATLAB package for performing stability selection is publicly available. In addition, we have built on features available in the R stabs package (Hofner et al., 2017) by extending stability selection to include an outlier filtering step and by providing access to feature selection methods other than Lasso.

Briefly, the procedure for stability selection can be broken up into 4 steps. Given regularization parameters Λ and *N* resamples. For *i* in 1… *N*:

1. Generate resampled dataset of some predetermined size (e.g., *n*/2)
2. Run a feature selection algorithm on the resampled dataset with regularization parameter λ to get selection set 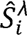
3. Calculate empirical selection probability: 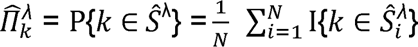
4. Given selection probabilities for each component and for each λ, construct the stable set: 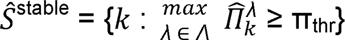

Given a stable set threshold of 0.5 through 1, we can estimate the upper bound on the expected number of falsely selected variables *V* using:

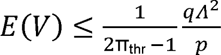

If the average number of features selected, *qΛ*, is known, π_thr_ can be estimated and vice-versa.

##### 2.8.6. Evaluating feature importance with Shapley Additive Explanations

Tuned models were extracted from the repeat that was closest to the median on model performance measures. SHAP values were collapsed across models and samples of the data to generate global measures of feature importance. For any sample in the data, the sign of a SHAP value reflects whether the prediction for that sample is pushed higher (positive) or lower (negative) than the baseline prediction (i.e., mean prediction). However, baseline predictions may themselves be lower or higher than true values. Thus, the sign of a SHAP value does not embed information about whether the feature contribution is overall beneficial to *successful* prediction. As the magnitude of a SHAP value alone reflects the influence of a feature on model output, we analyzed absolute SHAP values. However, we also analyzed raw SHAP values that were inverse transformed so that *positive* SHAP always reflected the *positive* influence of a feature on model prediction for a sample. Few samples required this transform, and we emphasize that transformed values fundamentally reflected patterns that the model learned for making predictions.

### 2. Results

#### 2.1 Relationships between theory-driven ROIs

When used for prediction, OLS regression requires more samples to estimate coefficients precisely and accurately, and relatively large sample sizes have been recommended purely for using OLS as an explanatory tool (e.g., recommendations for the data to contain 10, 30, or more samples for each feature; Algina & Olejnik, 2000; Knofczynski & Mundfrom, 2008; Miller and Kunce, 1973; Pedhazur & Schmelkin, 1991). Thus, one strategy for model improvement is to further reduce the number of features that are fit, which would additionally help address multicollinearities in the data (see Figure S1) that can inflate standard errors of the estimated coefficients. At the same time, successful model prediction may hinge as much on filtering out noise and redundancy as adding more signal to the feature set. That is, adding features outside the putative object naming network as established by prior work, much of which has relied on neuroimaging modalities other than perfusion. Evidence that both of these strategies could improve model performance can be seen in the correlations presented in Figure 2 of the main manuscript, which shows that our preselected ROIs contain features unassociated with naming and miss features with strong associations to naming.

Figure S1 reveals multicollinearities in the set of left hemisphere theory-driven ROIs, consistent with our findings that lasso, ridge, and elastic net all perform substantially better than simple OLS regression when applied to these ROIs (see Figure 3B and C). Figure S1 shows Pearson correlations between perfusion in each pair of left hemisphere theory-driven ROIs that form the putative object naming network (Figure S1). These relationships reveal the presence of a number of potentially ‘redundant’ features that show similar perfusion to most other features in the set, as well as features that are potentially unique. For example, the left middle frontal gyrus exhibits perfusion correlating with almost all features except left fusiform gyrus, left thalamus, and left subgenual cingulate gyrus at p < 0.05. In contrast, the left fusiform gyrus does not correlate significantly with any feature except the left pole of the middle temporal gyrus at p < 0.05. Notably, the left fusiform gyrus also did not correlate significantly with naming, however, the left thalamus overall shows a similar pattern and a significant correlation with naming.

**Figure S1:**
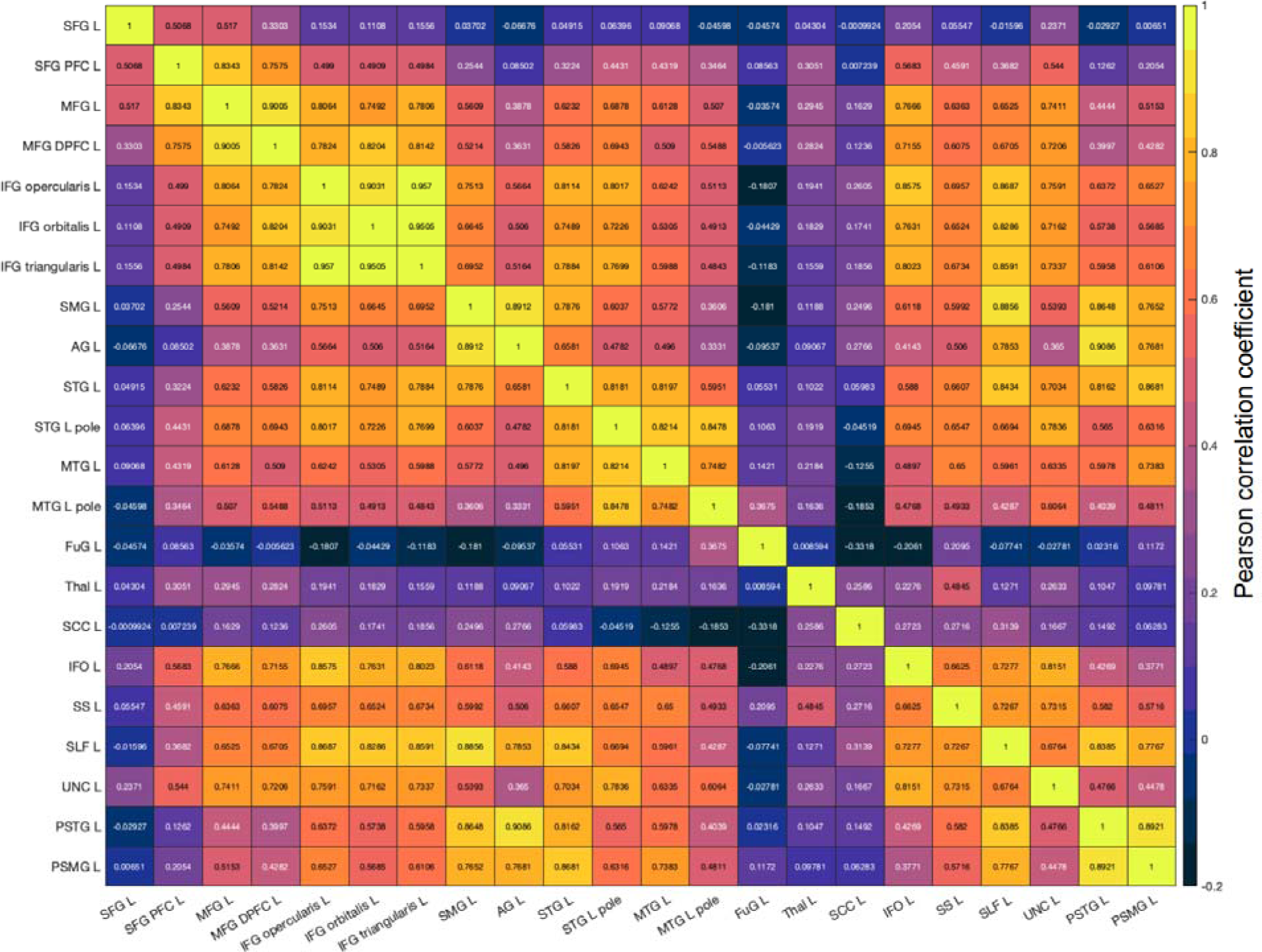
Associations between theory-driven features. Matrix showing the Pearson correlation coefficients between perfusion in each pair of ROIs in the putative object naming network. Brighter colors represent stronger associations. For reference, cutoff for significance at p < 0.05 is r = 0.31 and p < 0.001 is r = 0.49.

#### 2.2 Improving model prediction with simple feature selection methods

We compared models utilizing different feature selection methods in several ways. Where possible, in the main text, we defaulted to performing comparisons between models based on performance measures tracked over repeats of cross-validation (i.e., for correlation and accuracy percent). However, some of the simple feature selection methods paired with OLS regression would produce identical results on every repeat (i.e., because no tuning was required for feature selection or model estimation). Thus, in order to streamline comparisons between simple feature selection methods paired with OLS regression, we extracted the median performing model across repeats (on accuracy percent, which is based on mean absolute error scaled to a naïve model and expressed in percent units). All model comparisons were then performed based on the absolute errors generated by the models. That is, we tested whether mean absolute error was significantly different for one model versus another, taking into account model variability across partitions when model building entailed some tuning that could impact errors (i.e., the outer validation loop used leave-one-out so results would be identical if no inner loop was used for tuning the model).

The results of these paired t-tests are presented in Figure S2A (p-values) and S2D (t-statistics). Significant effects are described in greater detail in the main text, where we noted that of the baseline models, theory-driven bilateral ROIs performed significantly worse than both theory-driven left hemisphere ROIs and no feature selection. Note, although there was no significant difference between the left hemisphere ROI and no feature selection models (p > 0.05), stability selection performed significantly better than the no feature selection model, t(42) = 2.4, p < 0.05 (see Table S1 for mean and standard deviations). Indeed, this was the only feature selection method to perform significantly better than the no feature selection model. Stability selection also significantly outperformed all stepwise models that relied on in-sample criteria (BIC: t(42) = 2.38, p < 0.05; AIC: t(42) = 2, p = 0.05; SSE: t(42) = 2.33, p < 0.05; adjusted R^2^: t(42) = 2.38, p < 0.05). Although sequential feature selection (i.e., stepwise regression with out-of-sample mean absolute error as the criterion) performed relatively well, its model errors were only significantly lower compared to bilateral theory-driven ROIs (t(42) = 3.7, p < 0.0001) and the significance based correlation filter (i.e., retaining all features significantly correlated at p < 0.05; t(42) = 2.4, p < 0.05).

Note, we additionally performed comparisons as initially described, by comparing model AP and correlation across repeats of cross-validation, where this was possible (Figures S2B,C,E,F). In Figure S3 we additionally show violin plots of the distribution of performance over repeats (see also Table S1 for summary measures). The patterns that emerged from these analyses were similar but generally tended to highlight better performance for stability selection compared to other models. One consistent pattern was that correlations between predictions and true values for stability selection were not significantly higher than sequential feature selection (p > 0.05). One difference, however, was that stability selection had higher correlations than other feature selection based on correlation filters (tuned filter size: t(19) = 18.2, p < 0.000001; consistency of p < 0.05 features: t(19) = 12.1, p < 0.000001). The AP measure evaluates model error in relation to a naïve model and showed a slightly different pattern. Here, stability selection performed significantly better than sequential feature selection, t(19) = 2.4, p < 0.00001. In contrast to previous findings for correlation and absolute errors, sequential feature selection did not improve on a naïve model by a significantly higher amount than feature selection using a correlation filter with a tuned window size (p > 0.05).

Violin plots in Figure S3 reinforce the inconsistency between accuracy percent (AP) and correlation as performance measures. For example, they highlight the discrepancy between surprisingly high correlations and low AP for theory-driven bilateral ROIs, which did not show many significant effects in the tests described in the previous section. In other words, AP signaled that some models with high ‘accuracy’ based on correlation performed disturbingly poorly when compared to a naïve model that guesses. Across analyses, we tended to see misleadingly high correlations only when accuracy percent or mean absolute error were particularly low. Violin plots also help highlight significant variability in the performance of some models, particularly forward sequential selection and the genetic algorithm. Variability in model performance was noticeably higher for correlation than accuracy percent, hinting that correlation may be a less stable performance metric in the presence of relatively high model errors.

**Figure S2:**
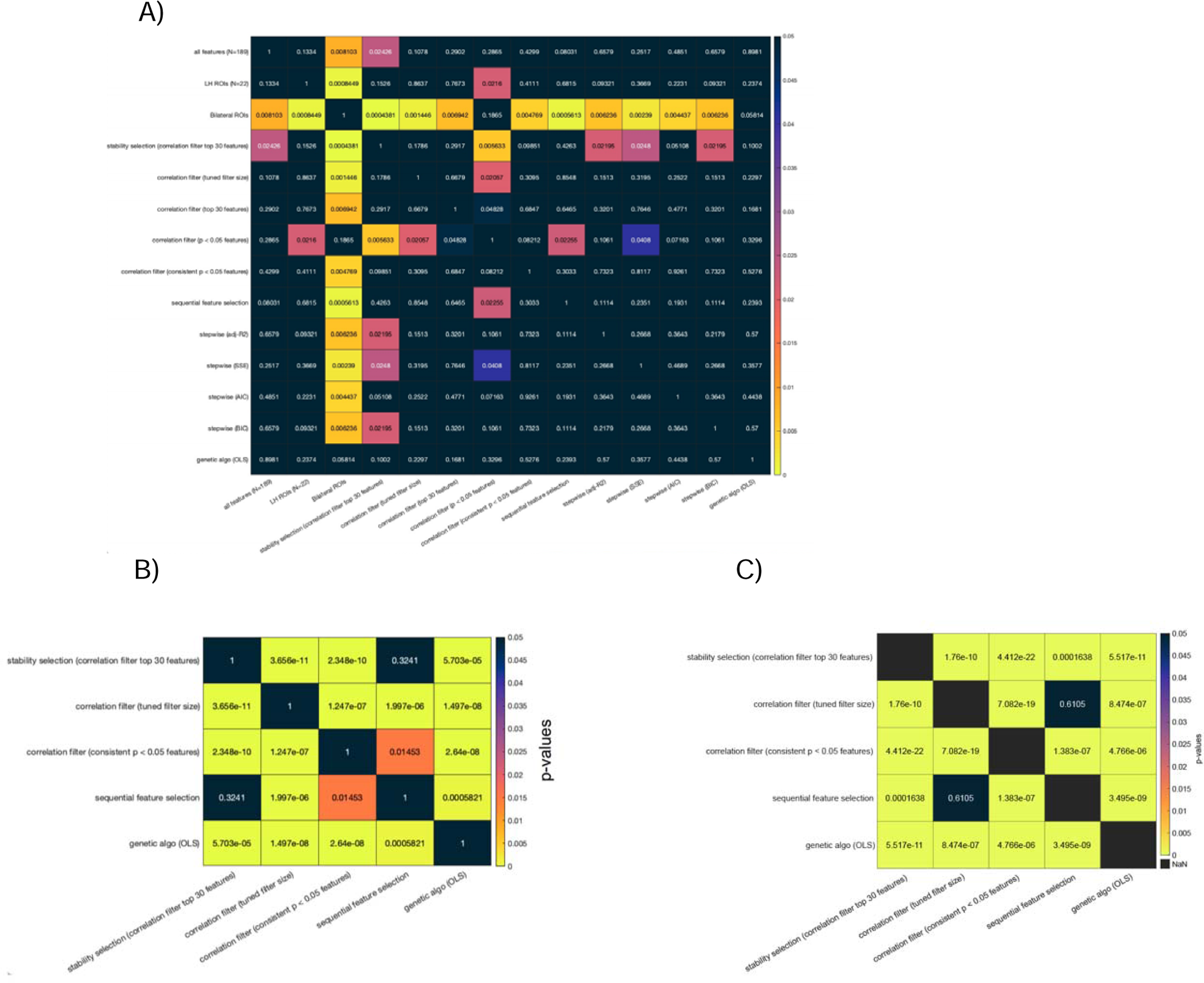
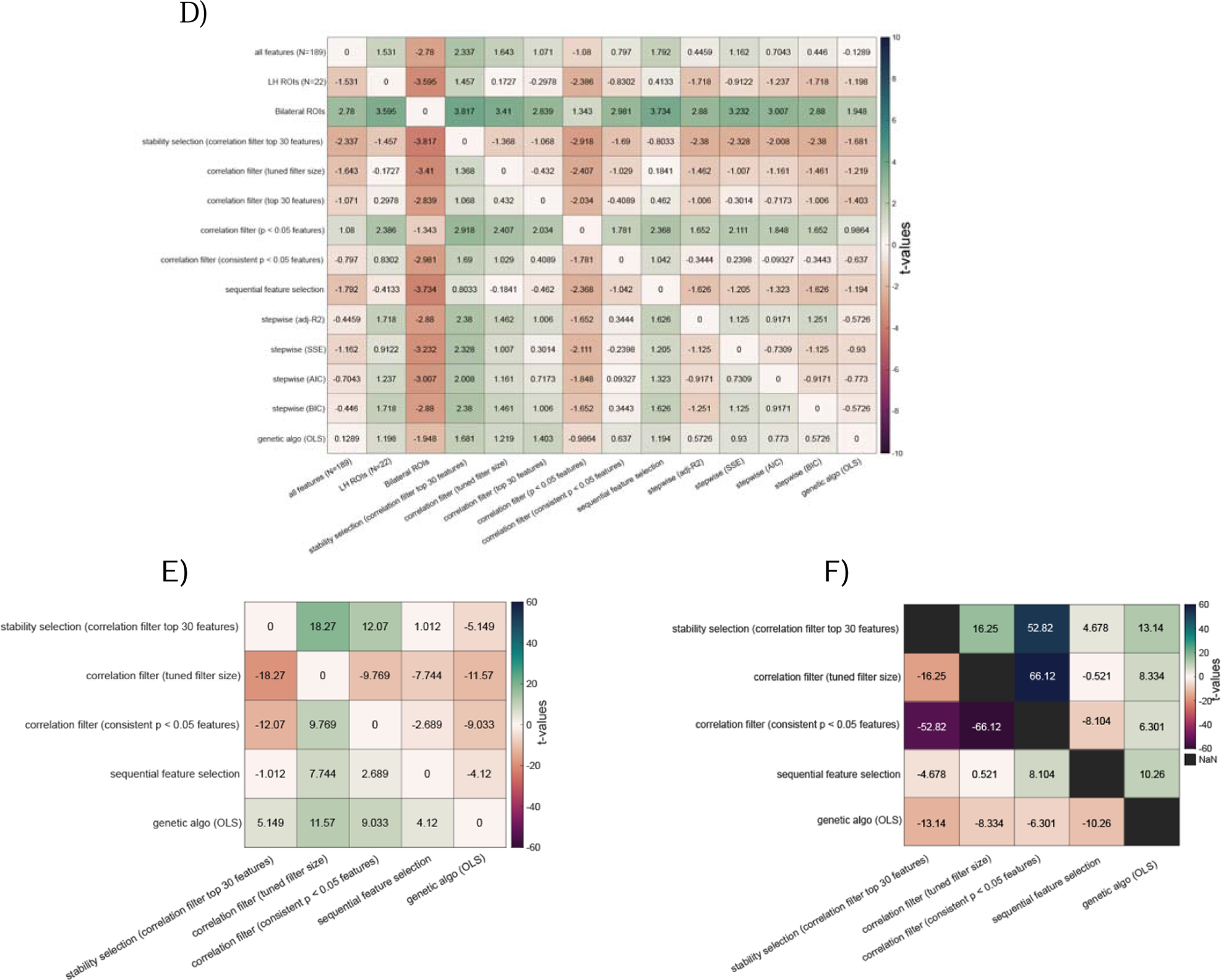
Model performance differences between simple feature selection methods paired with OLS linear regression for model estimation. All panels show information about pairwise comparisons between models based on paired t-tests. Panels A, B and C show the resulting p-values of pairwise tests and panels D, E and F show the corresponding t-statistics. In panels displaying p-values, insignificant models (p > 0.01) are assigned black colors and hotter colors represent lower p-values. In panels displaying t-statistics, more saturated colors represent stronger t-statistics. Higher positive t-statistics (cooler colors) represent a larger mean for the row than the column, and lower negative t-statistics (hotter colors) represent a larger mean for the column than the row. **Panels A and D:** Differences in absolute prediction errors for median performing models as determined over 20 repeats of cross-validation (note, not all models required tuning and therefore produced the same result on each repeat). Panel A shows p-values while panel D shows t-statistics. **Panels B and E:** Differences in correlation coefficient across models as the measure of performance tracked across 20 repeats of cross-validation. Panel B shows p-values and panel E shows t-statistics for each comparison. **Panel C and F:** Differences in accuracy percent between models. Panel C shows p-values and panel F shows t-statistics for comparisons.

**Figure S3:**
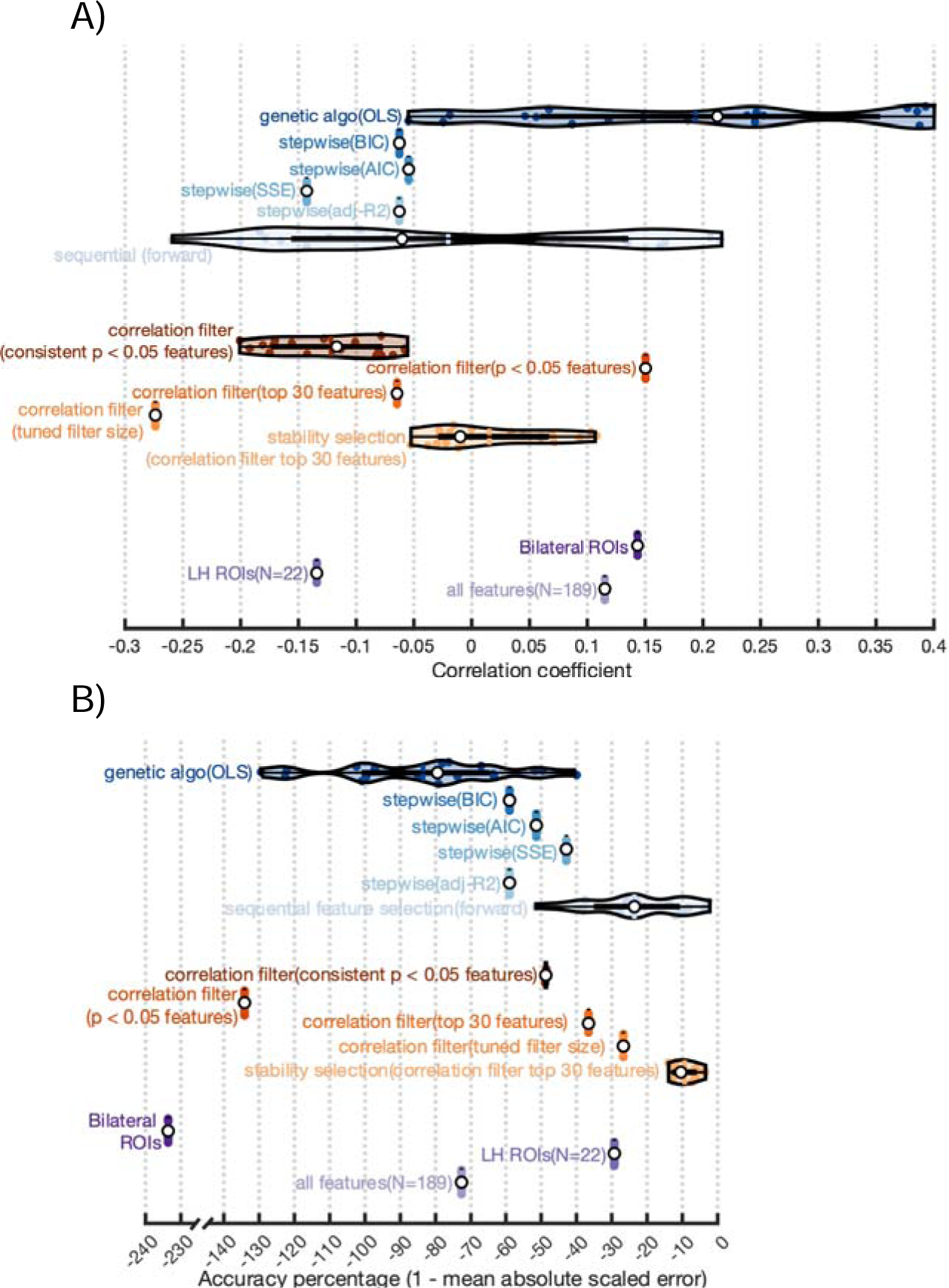
Violin plots showing distribution of model performance for simple feature selection paired with OLS linear regression for model estimation. **Panel A:** Performance is measured by correlations between true naming scores and model predictions. **Panel B:** Performance is measured by percent improvement of predictions over a naïve model that guesses using the mean of the training data (i.e., accuracy percent). In both panels, the white dot represents the median performing model. The vertical line inside each violin shows the mean performing model. Other colored dots inside the violin represent the remaining models’ performance. Black horizontal lines represent a box plot.

**Table S1:** Mean and standard deviation of performance measures for models with simple feature selection and OLS linear regression.

| Model | MEAN<br>(correlation) | SD<br>(correlation) | MEAN<br>(accuracy<br>percent) | SD<br>(accuracy<br>percent) | MEAN<br>(absolute<br>error) | SD<br>(absolute<br>error) |
| --- | --- | --- | --- | --- | --- | --- |
| all features (N=189) | 0.1151 | 0 | -72.64 | 0 | 54.972 | 53.309 |
| LH ROIs (N=22) | -0.134 | 0 | -29.28 | 0 | 41.164 | 30.336 |
| Bilateral ROIs | 0.1434 | 0 | -233.6 | 0 | 106.22 | 129.10 |
| stability selection<br>(correlation filter top 30) | 0.0152 | 0.0521 | -9.346 | 3.3605 | 35.120 | 28.599 |
| correlation filter<br>(tuned filter size) | -0.273 | 0 | -26.71 | 0 | 40.347 | 24.779 |
| correlation filter<br>(top 30 features) | -0.064 | 0 | -36.59 | 0 | 43.494 | 48.305 |
| correlation filter<br>(p < 0.05 features) | 0.1503 | 0 | -134.1 | 0 | 74.551 | 97.497 |
| correlation filter<br>(consistent p < 0.05 features) | -0.121 | 0.0482 | -48.71 | 0.0707 | 47.348 | 34.987 |
| sequential feature selection | -0.020 | 0.1529 | -23.60 | 13.820 | 39.268 | 32.966 |
| stepwise (adj-R2) | -0.062 | 0 | -58.98 | 0 | 50.620 | 46.242 |
| stepwise (SSE) | -0.142 | 0 | -42.88 | 0 | 45.496 | 34.143 |
| stepwise (AIC) | -0.054 | 0 | -51.39 | 0 | 48.206 | 44.346 |
| stepwise (BIC) | -0.062 | 0 | -58.97 | 0 | 50.620 | 46.242 |
| genetic algo (OLS) | 0.1936 | 0.1545 | -83.55 | 24.726 | 57.030 | 89.215 |

#### 2.3 Introducing regularization to feature selection and model fitting

We first introduced regularization into feature selection and model estimation by using stability selection with elastic net and ridge regression for model fitting. Note, we chose ridge regression here as it applies an intuitive penalty term to OLS regression in order to control model complexity without performing any feature selection. Ridge regression was also extended to theory-driven feature selection models for comparison. The results of all pairwise model comparisons on accuracy percent and correlation that are summarized in Figure 3B from the main text are shown below in Figure S4. Here, we focus on effects not reported in the main text: comparisons based on correlation across repeats. First, stability selection with elastic net and ridge regression for model estimation outperformed stability selection with a correlation filter and OLS regression for estimation (elastic net: M=0.23,SD=0.05; correlation filter: M=0.015,SD=0.05), t(19) = 11.5, p < 0.000001. Correlation-based evaluation of baseline models showed the same pattern as AP. Correlations were higher for stability selection than left hemisphere ROI (M=0.07,SD=0.04), t(19) = 11.344, p < 0.0001, bilateral ROI (M=0.13,SD=0.08), t(19) = 4.4, p < 0.001, and no feature selection (M=-0.12,SD=0.04) models, t(19) = 23.51, p < 0.0001. In contrast to our results with OLS regression, here, the bilateral ROI model (AP: M=2,SD=2.3; Correlation: M=0.13,SD=0.08) performed significantly better than the left hemisphere ROI model (AP: M=-0.89,SD=1.25; Correlation: M=0.08,SD=0.04), AP: t(19) = 4.81, p < 0.001; Correlation: t(19) = 2.77, p < 0.05. Overall, no feature selection performed significantly worse than the baseline models. As the previous section, distribution of model performance is presented in violin plots for more detail (Figure S5). Additionally, summary measures for these models are reported in Table S2.

**Figure S4:**
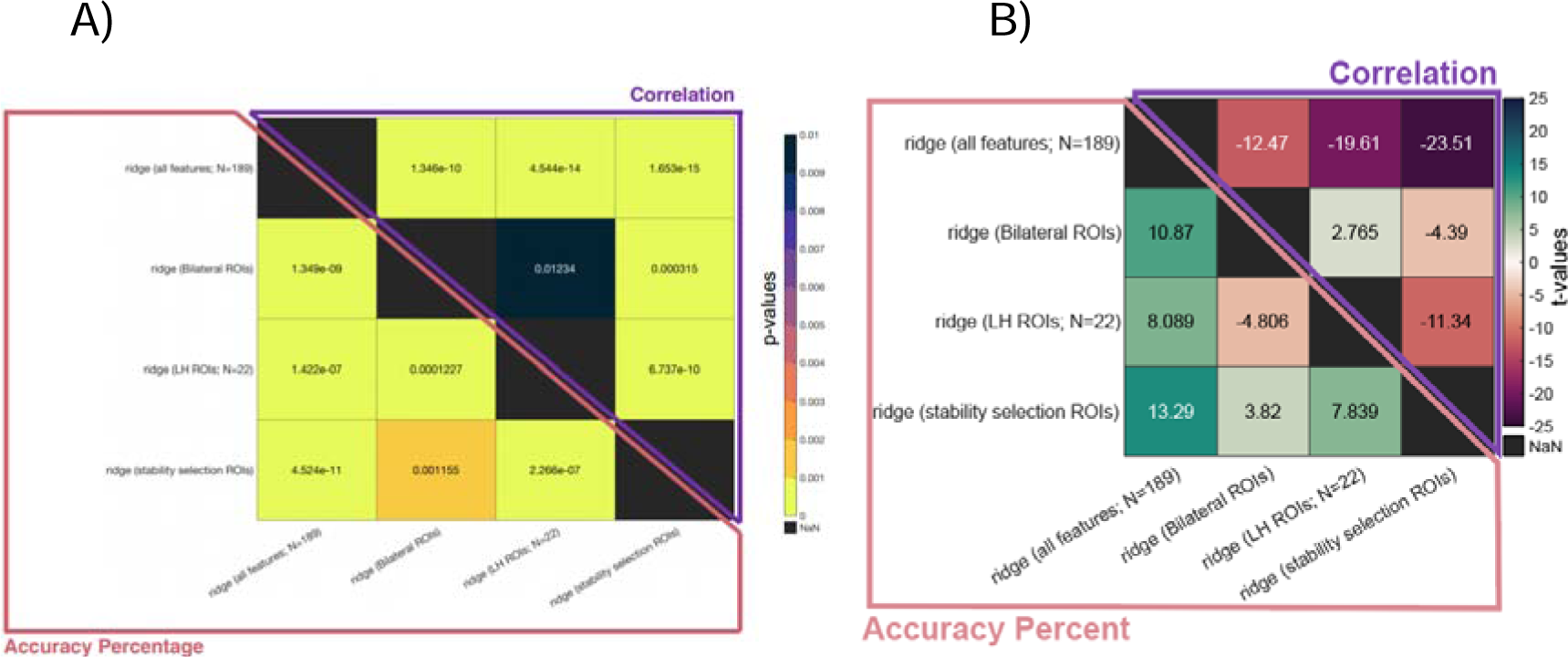
Comparing stability selection with elastic net to theory-driven feature selection when paired with ridge regression for model estimation. See previous Figure S2 for more information on how matrices are constructed. **Panel A:** Each cell shows the p-value for pairwise model comparisons according to a paired t-test. Note, here, the upper triangle represents correlation as the performance measure and the lower triangle represents accuracy percent. **Panel B:** Each cell shows the t-statistic for pairwise model comparisons.

**Figure S5:**
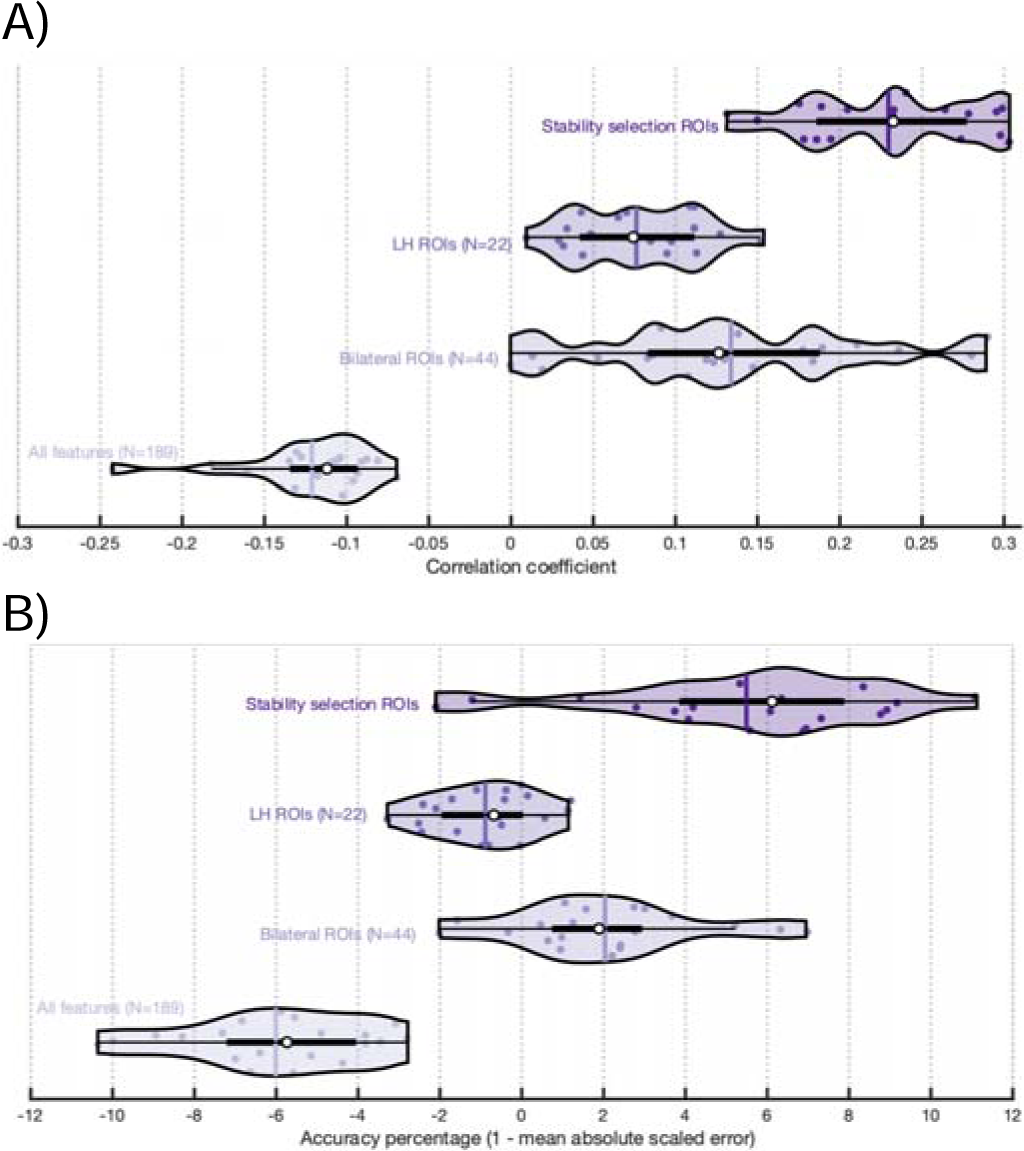
Violin plots showing distribution of model performance over repeats of cross-validation for stability selection and theory-driven feature selection paired with ridge regression for model estimation. **Panel A:** performance is measured as the correlation coefficient between true and predicted naming scores. **Panel B:** performance is measured as improvement in absolute errors scaled to a naïve model that guesses based on the mean training data. See Figure S3 for more information about details of the violin plot.

#### 2.4 Pairing stability selection with regularized models that embed feature selection and dimensionality reduction

We extended our experiments with regularization by implementing a number of different model estimation techniques that embedded feature selection or dimensionality reduction steps. In other words, we repeated our previous analysis, but instead of using ridge regression, we attempted to estimate models using a variety of different methods that embedded dimensionality reduction or feature selection steps.

In the main text, we first focused on comparing the model which combines stability selection and ridge regression to other models which combine theory-driven feature selection with various estimation techniques: partial least squares (PLS), partial least squares with variable importance projection (PLS-VIP), elastic net, and lasso. The results of all model comparisons summarized in Figure 3C in the main text are shown below in Figure S6 and the violin plots of model performance across repeats are presented in Figure S7. Summary measures are presented in Table S2. Given the number of comparisons, we point the reader to the figures for the full set of test results. Here, we will summarize some of the significant model differences that were omitted in the main text, referring to p-values alone for brevity. We focus on AP.

When no feature selection was applied, ridge regression performed as well as lasso, PLS, and PLS-VIP (p > 0.05). In contrast, elastic net underperformed both ridge and lasso (p < 0.01) but performed as well as PLS and PLS-VIP (p > 0.05). We speculate this is due to the additional hyperparameter tuning required for elastic net, which failed in the context of the wider dataset. PCR performed worse than the other models (p < 0.0001). That ridge regression (i.e., no feature selection) performs so well against other methods with internal feature selection or dimensionality reduction suggests that not all data-driven feature selection improves models.

Using bilateral ROIs improved performance of ridge (p < 0.00001), elastic net (p < 0. 00001), PCR (p < 0.00001), and PLS-VIP (p < 0. 00001) relative to no feature selection. However, bilateral ROIs performed equally well to no feature selection for lasso and PLS (p > 0.05). Focusing on the smaller subset of left hemisphere ROIs significantly improved the performance of lasso, elastic net, PCR, and PLS compared to bilateral ROIs. Curiously, bilateral ROIs performed better than left hemisphere ROIs or no feature selection for PLS-VIP and ridge regression (p < 0.01). Thus, theory-driven feature selection boosted model performance in all cases (i.e., versus no feature selection), however, the inclusion of right hemisphere regions was sometimes opportune and other times detrimental to model performance. Correlation as the performance measure showed an overall similar pattern although we are hesitant to interpret differences at small correlation values as our results demonstrate these models tend to perform more inconsistently relative to AP.

Overall, using any method that selects features and estimates the model simultaneously performed worse than stability selection with ridge regression for model estimation. In other words, not all data-driven feature selection performed well, and an external feature selection step appeared to improve models. However, it is worth pointing out that some theory-driven models performed as well or better than this strategy. Specifically, PLS-VIP with bilateral ROIs performed slightly better than ridge regression with stability selection (p < 0.05), and elastic net with left hemisphere ROIs performed as well as ridge with stability selection (p > 0.05). We speculate PLS benefitted more from right hemisphere features for estimating the latent dimensions more precisely, whereas elastic net benefitted from a reduced feature space that enabled more robust hyperparameter tuning.

Notably, using the same estimators with stability selection ROIs tended to result in better models. Stability selection significantly improved ridge regression, lasso, and elastic net (p < 0.05). The best theory-driven models for PLS-VIP (bilateral ROIs, p < 0.0001), PLS (left hemisphere ROIs, p < 0.05) and PCR (left hemisphere ROIs, p < 0.0001) outperformed stability selection. In each of these cases, however, stability selection outperformed the second-best performing theory-driven model. Moreover, and as we emphasize in the main text, stability selection paired with lasso outperformed all other models by a large margin (p < 0.05).

**Figure S6:**
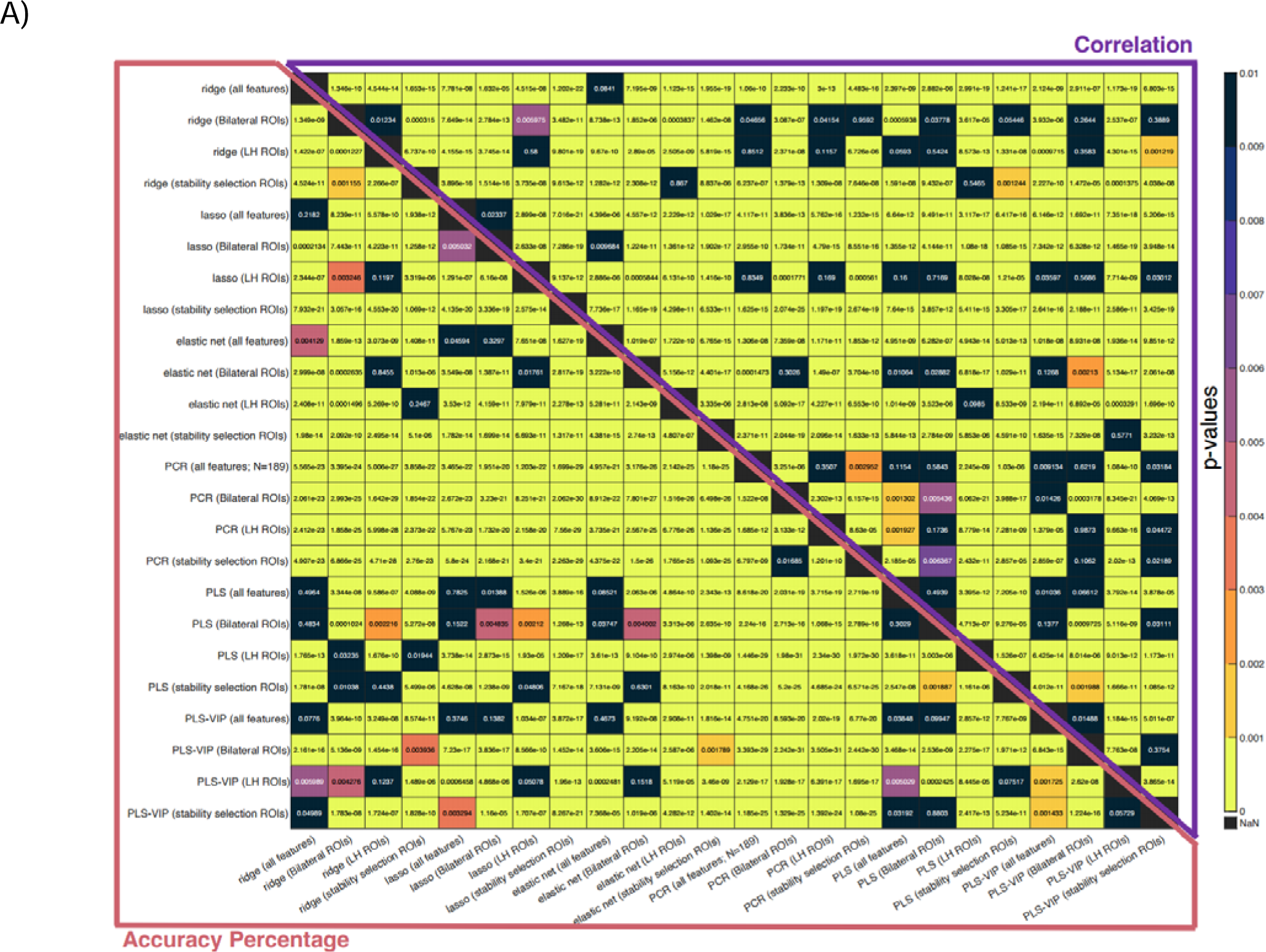
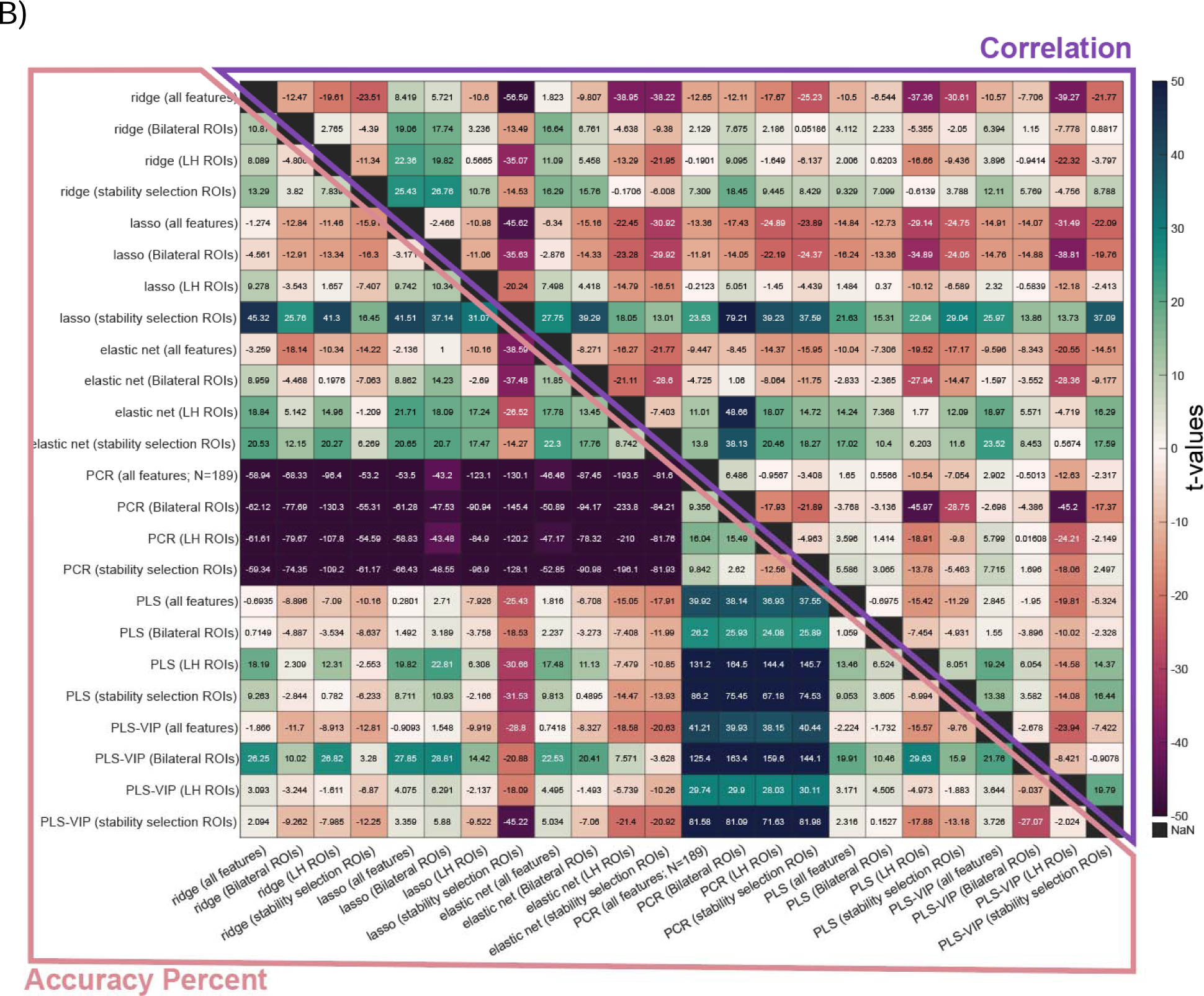
Comparing stability selection with elastic net to theory-driven feature selection when paired with model estimation techniques that embed feature selection or dimensionality reduction. See previous Figure S2 for more information on how matrices are constructed. **Panel A:** Each cell shows the p-value for pairwise model comparisons according to a paired t-test. **Panel B:** Each cell shows the t-statistic for pairwise model comparisons. Note, here, the upper triangle represents correlation as the performance measure and the lower triangle represents accuracy percent.

**Figure S7:**
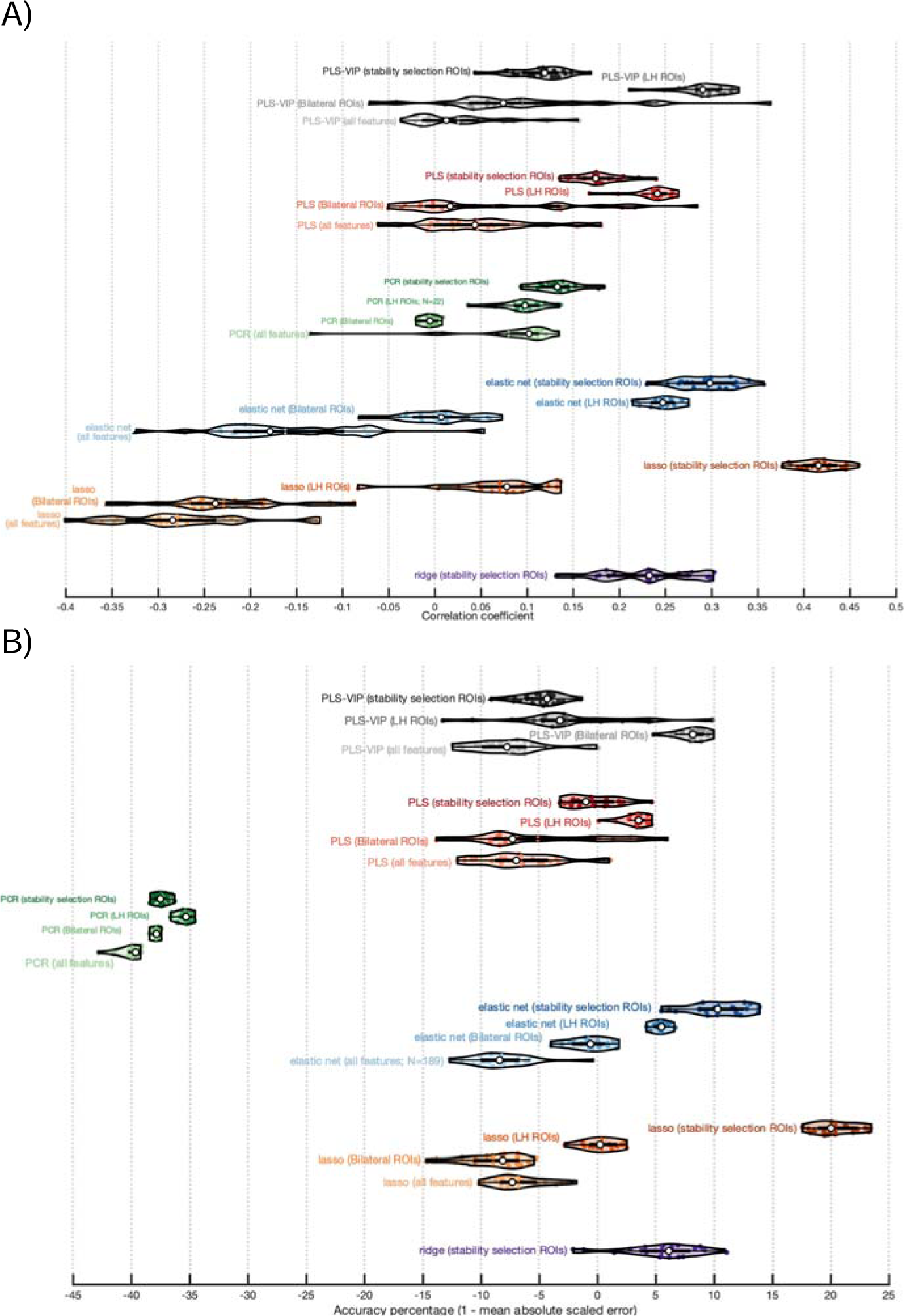
Violin plots showing distribution of model performance over repeats of cross-validation for stability selection and theory-driven feature selection paired with model estimation techniques that embed feature selection or dimensionality reduction. **Panel A:** Performance is measured as correlation coefficient between true and predicted naming scores. **Panel B:** Performance is measured as accuracy percent. See Figure S3 for more information on the composition of violin plots.

**Table S2:** Mean and standard deviation of performance measures for models with stability selection and theory-driven feature selection paired with model estimation techniques that embed feature selection or dimensionality reduction.

| Model | MEAN<br>(correlation) | SD<br>(correlation) | MEAN<br>(accuracy<br>percent) | SD<br>(accuracy<br>percent) |
| --- | --- | --- | --- | --- |
| ridge (all features; N=189) | -0.121 | 0.0394 | -6.010 | 2.2064 |
| ridge (Bilateral ROIs) | 0.1338 | 0.0826 | 2.0355 | 2.3123 |
| ridge (LH ROIs; N=22) | 0.0762 | 0.0381 | -0.889 | 1.2498 |
| ridge (stability selection ROIs) | 0.2295 | 0.0522 | 5.4909 | 3.3830 |
| lasso (all features; N=189) | -0.279 | 0.0718 | -6.819 | 2.2212 |
| lasso (Bilateral ROIs) | -0.229 | 0.0676 | -8.966 | 2.6448 |
| lasso (LH ROIs; N=22) | 0.0700 | 0.0581 | 0.3418 | 1.4973 |
| lasso (stability selection ROIs) | 0.4169 | 0.0230 | 20.410 | 1.8226 |
| elastic net (all features; N=189) | -0.162 | 0.0850 | -8.238 | 2.6449 |
| elastic net (Bilateral ROIs) | 0.0035 | 0.0409 | -0.792 | 1.7329 |
| elastic net (LH ROIs; N=22) | 0.2468 | 0.0188 | 5.4028 | 0.6368 |
| elastic net (stability selection ROIs) | 0.2959 | 0.0320 | 10.376 | 2.4702 |
| PCR (all features; N=189) | 0.0798 | 0.0613 | -39.94 | 0.9508 |
| PCR (Bilateral ROIs) | -0.006 | 0.0091 | -37.90 | 0.2884 |
| PCR (LH ROIs; N=22) | 0.0953 | 0.0244 | -35.40 | 0.5714 |
| PCR (stability selection ROIs) | 0.1328 | 0.0246 | -37.51 | 0.5636 |
| PLS (all features; N=189) | 0.0463 | 0.0614 | -6.566 | 3.5942 |
| PLS (Bilateral ROIs) | 0.0636 | 0.0984 | -5.129 | 5.6240 |
| PLS (LH ROIs; N=22) | 0.2374 | 0.0215 | 3.3817 | 1.0431 |
| PLS (stability selection ROIs) | 0.1769 | 0.0266 | -0.453 | 2.1786 |
| PLS-VIP (all features; N=189) | 0.0253 | 0.0515 | -7.566 | 3.3100 |
| PLS-VIP (Bilateral ROIs) | 0.0949 | 0.1008 | 8.1447 | 1.1980 |
| PLS-VIP (LH ROIs; N=22) | 0.2906 | 0.0271 | -2.634 | 5.2203 |
| PLS-VIP (stability selection ROIs) | 0.1147 | 0.0291 | -4.932 | 1.7485 |

#### 2.5 Testing stability selection against other relatively complex external feature selection approaches

While it’s clear that stability selection outperforms common models that combine feature selection and estimation, it’s not entirely clear from the results we have presented thus far that other external feature selection approaches would not produce equally good results when paired with lasso. To test this, we reintroduced some of the feature selection methods that we initially experimented with in the context of OLS regression, this time fitting models with lasso as well as elastic net. We include elastic net to ensure that none of these methods show a strong preference for less aggressive ‘supervision’ (i.e., feature selection). Dimensionality reduction approaches were not tested as they tended to perform slightly worse. In this analysis, we attempted to improve on the reintroduced feature selection methods and also introduced more complex feature selection techniques (see methods). For more information on the described effects, refer to Figures S8, S9 and Table S3.

One reintroduced method was stepwise regression with SSE. This method performed worse than any other that we tested when combined with lasso or elastic net. We additionally tested the top 30 correlation filter from Figure 3A, which performed relatively well in that analysis without the additional training time required to tune the filter window. While lasso and elastic net improved model performance with this filter compared to OLS linear regression, the difference was relatively small (c.f., AP of −16% versus −36%). When paired with OLS linear regression, sequential feature selection using out-of-sample mean absolute error on OLS linear regression was one of the best performing feature selection methods. Given the success of elastic net in other analyses, we used this method in lieu of OLS linear regression for forward sequential feature selection. However, this approach performed roughly as well as the simpler method, with models achieving AP of roughly −20%. Two new feature selection methods (i.e., NCA and RreliefF) were introduced and performed as well when paired with lasso (AP of −10%). When these methods were paired with elastic net, they performed significantly worse (p < 0.05).

Overall, methods that involved more hyperparameters tended to show higher variance in performance, reflecting tuning difficulties in the exploding hyperparameter space (e.g., NCA, EN, RReliefF, and sequential forward selection with EN). This does not explain all of the variance, however. Evidence that some models may simply be unstable in the context of our dataset comes from lasso and elastic net as external feature selection steps. In both cases, estimating the model either using lasso or elastic net produced roughly the same standard deviations in performance, despite the fact that tuning an elastic net introduced an additional hyperparameter. Thus, different datasets may be more amendable to different feature selection approaches.

The most striking pattern across external feature selection methods was that most tended to perform worse than a naïve model. The only exceptions were stability selection and the genetic algorithm. Further, stepwise regression with in-sample SSE performed significantly worse than all other methods except for NCA fit with lasso. This pattern was present irrespective of the model used for fitting stepwise regression (p < 0.05). The method of model fitting had no impact on the genetic algorithm, but stability selection performed much better when the model was fit with lasso. One explanation for this observation is that stability selection both identified more signal features and more noise features compared to the genetic algorithm. The additional noisy features would explain why stability selection benefitted from more aggressive ‘supervision’ by lasso. At the same time, stability selection identified a better ROI set as it produced a significantly better model when fit with lasso and at the same time produced a model with comparable performance to the genetic algorithm when it was fit with elastic net (i.e., p > 0.05 for comparison between stability selection fit with elastic net and genetic algorithm fit with lasso or elastic net).

**Figure S8:**
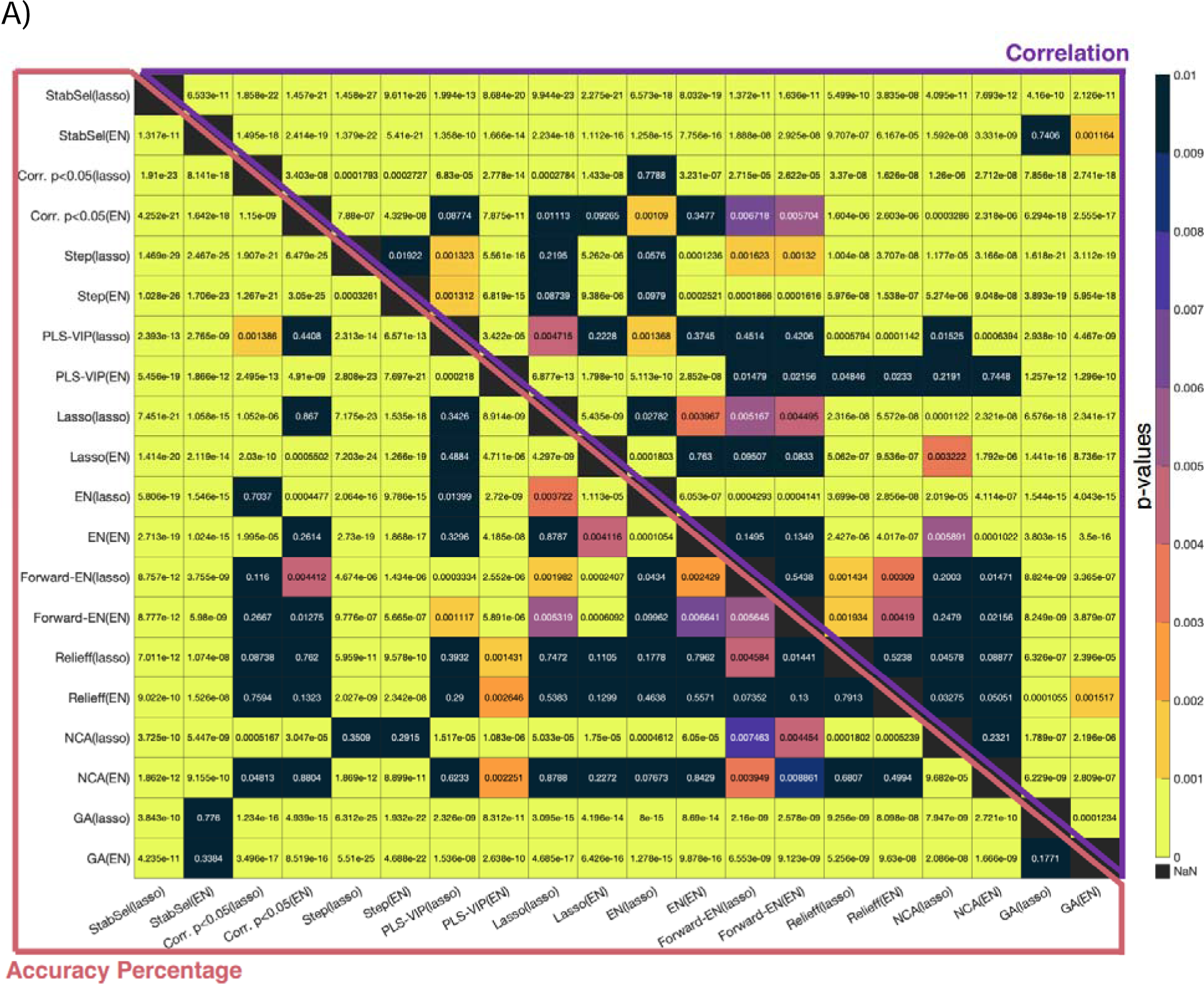
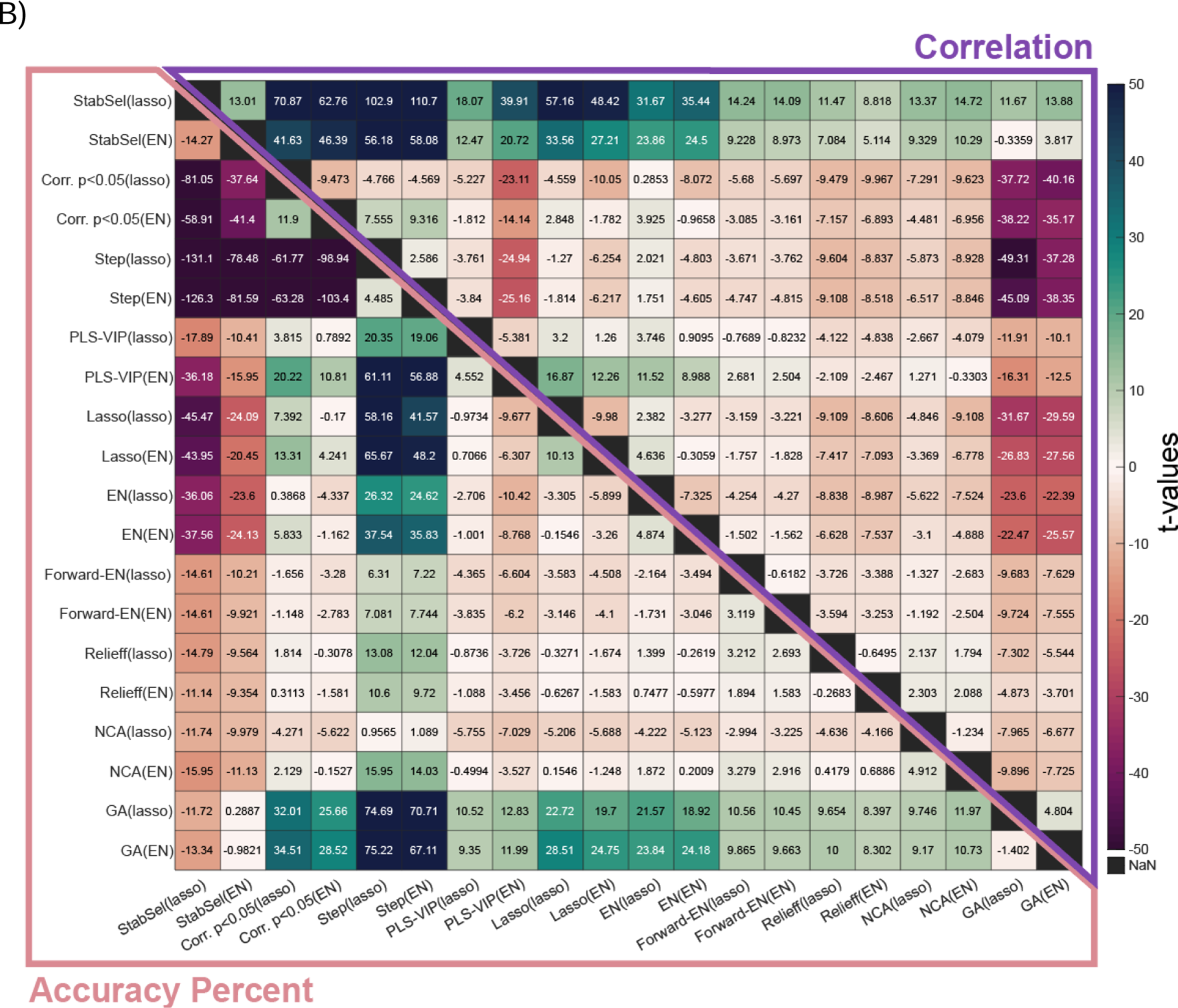
Comparing stability selection to alternative external feature selection methods. See previous Figure S2 for more information on how matrices are constructed. Each label describes the feature selection method, with the method used to fit models described in parentheticals. **Panel A:** Each cell shows the p-value for pairwise model comparisons according to a paired t-test. **Panel B:** Each cell shows the t-statistic for pairwise model comparisons. Note, the upper triangle in both panels shows comparisons based on correlation and the bottom triangle shows comparisons based on accuracy percent.

**Figure S9:**
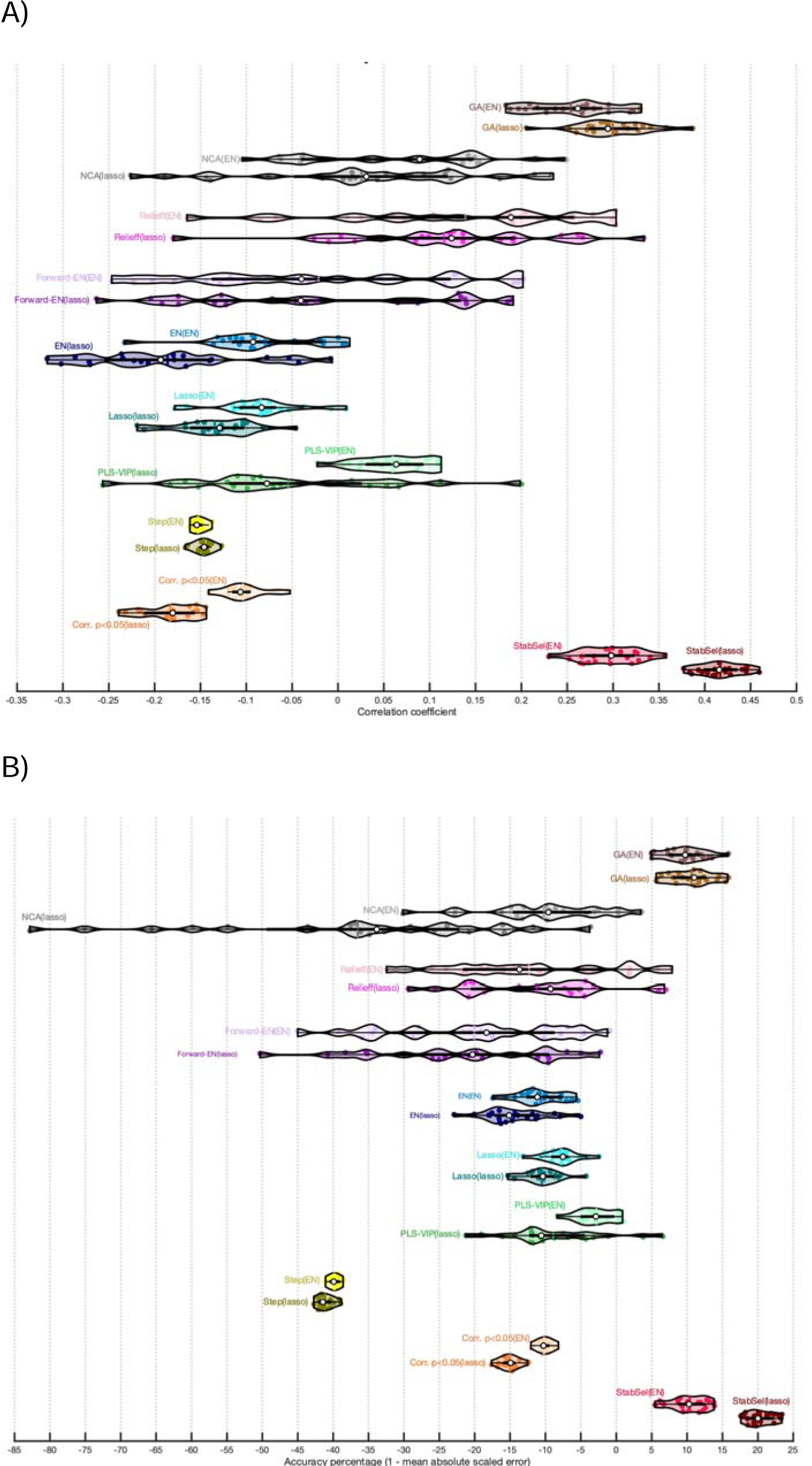
Violin plots showing distribution of model performance over repeats of cross-validation for stability selection and alternative external feature selection methods. **Panel A:** Performance is measured as correlation coefficient between true and predicted naming scores. **Panel B:** Performance is measured as accuracy percent.

**Table S3:**
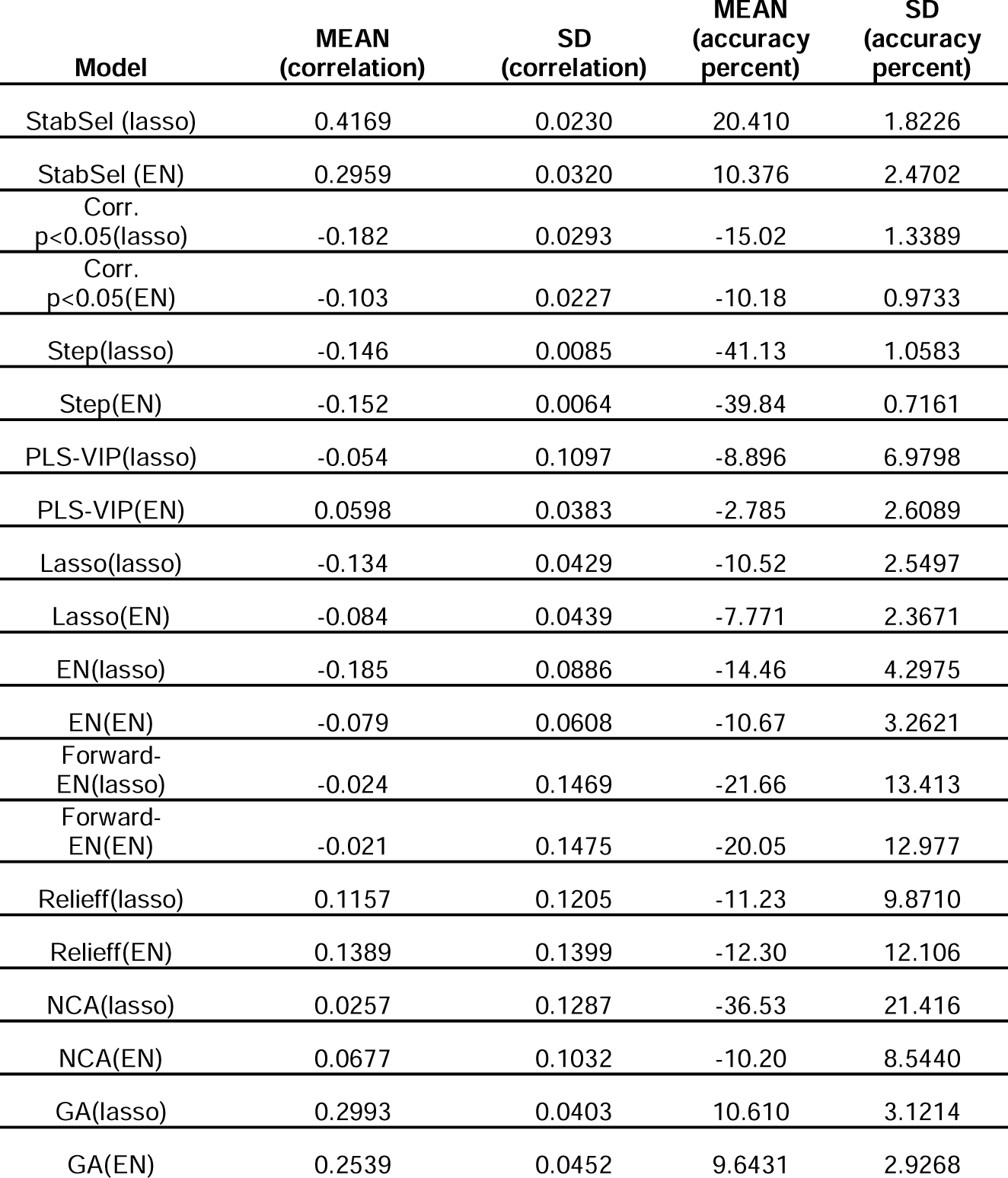
Mean and standard deviation of performance measures for models utilizing alternative external feature selection techniques to stability selection paired with model estimation techniques that embed feature selection or dimensionality reduction.

| Model | MEAN<br>(correlation) | SD<br>(correlation) | MEAN<br>(accuracy<br>percent) | SD<br>(accuracy<br>percent) |
| --- | --- | --- | --- | --- |
| StabSel (lasso) | 0.4169 | 0.0230 | 20.410 | 1.8226 |
| StabSel (EN) | 0.2959 | 0.0320 | 10.376 | 2.4702 |
| Corr.<br>p<0.05(lasso) | -0.182 | 0.0293 | -15.02 | 1.3389 |
| Corr.<br>p<0.05(EN) | -0.103 | 0.0227 | -10.18 | 0.9733 |
| Step(lasso) | -0.146 | 0.0085 | -41.13 | 1.0583 |
| Step(EN) | -0.152 | 0.0064 | -39.84 | 0.7161 |
| PLS-VIP(lasso) | -0.054 | 0.1097 | -8.896 | 6.9798 |
| PLS-VIP(EN) | 0.0598 | 0.0383 | -2.785 | 2.6089 |
| Lasso(lasso) | -0.134 | 0.0429 | -10.52 | 2.5497 |
| Lasso(EN) | -0.084 | 0.0439 | -7.771 | 2.3671 |
| EN(lasso) | -0.185 | 0.0886 | -14.46 | 4.2975 |
| EN(EN) | -0.079 | 0.0608 | -10.67 | 3.2621 |
| Forward-EN(lasso) | -0.024 | 0.1469 | -21.66 | 13.413 |
| Forward-EN(EN) | -0.021 | 0.1475 | -20.05 | 12.977 |
| Relieff(lasso) | 0.1157 | 0.1205 | -11.23 | 9.8710 |
| Relieff(EN) | 0.1389 | 0.1399 | -12.30 | 12.106 |
| NCA(lasso) | 0.0257 | 0.1287 | -36.53 | 21.416 |
| NCA(EN) | 0.0677 | 0.1032 | -10.20 | 8.5440 |
| GA(lasso) | 0.2993 | 0.0403 | 10.610 | 3.1214 |
| GA(EN) | 0.2539 | 0.0452 | 9.6431 | 2.9268 |

#### 2.6 Relationship between accuracy percent and correlation performance measures

When comparing the relationship between accuracy percent and correlation, we have focused on the association across different modeling methods for their respective median performing models as determined over 20 repeats of cross-validation. In general, we have observed the tendency for correlation to be misleadingly high when models perform poorly overall. For example, when we tested simple feature selection approaches with OLS regression (i.e., Figure 3A), we found that all models tended to perform quite poorly according to both correlation and AP measures (i.e., highest median correlation coefficient was < 0.25 and highest AP was < 0). In this context, models with relatively lower AP were associated with higher correlations (see main text for effect). However, when exploring other feature selection methods (i.e., Figure 3B, C, and D), we found a generally positive association across methods. Below we show the same relationship between correlation and AP but evaluated across repeats of CV for each individual method. Most methods showed a strong positive association, however, some methods did not, including PLS-VIP fit to theory-driven ROIs or ridge fit to all features. Indeed, the ridge model fit to all features showed an inverse relationship on the cusp of significance (see Figure S10). Given that all of these models tended to perform relatively well compared to others that showed a highly robust positive association between AP and correlation measures (e.g., >0.9), it cannot be concluded that higher correlations between true values and model predictions reflect lower model errors. Note, again, AP is the mean absolute error (MAE) of a model’s predictions scaled to the MAE of a naïve model. As the figures from the main text illustrate, individual naïve model predictions are highly consistent because the mean across maximally overlapping training datasets (i.e., LOOCV) is remarkably similar. Further, MAE of the naïve model does not differ across repeats of CV (i.e., LOOCV cannot be repartitioned). Thus, variance in AP over repeats of CV principally represents MAE.

**Figure S10:**
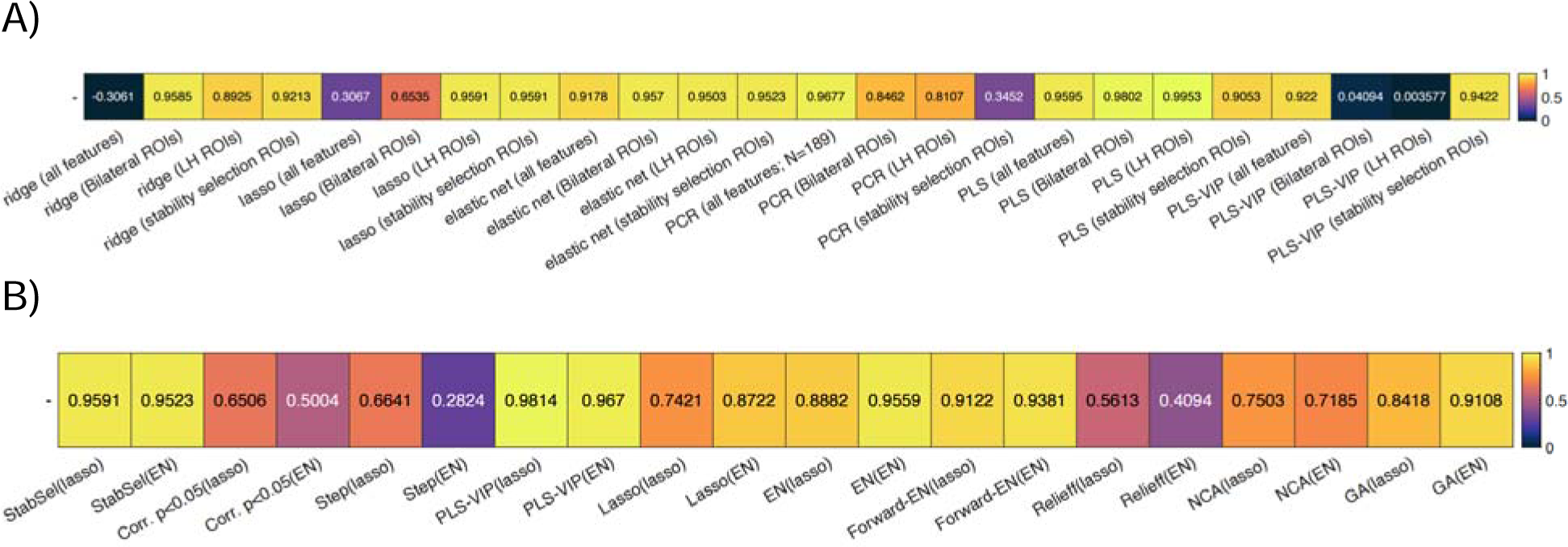
Association between correlation and accuracy percent across repeats. For both panels, each cell describes the correlation coefficient between accuracy percent and correlation performance measures as tracked over 20 repeats of cross validation for a particular combination of feature selection and model fitting procedure. **Panel A:** associations are shown between performance measures for algorithms that have internal feature selection or dimensionality reduction. **Panel B:** associations are shown between performance measures for external feature selection algorithms that represent alternatives to stability selection. Values above 0.31 or below −0.31 were statistically significant.

#### 2.7 More complex methods of model estimation paired with stability selection

Stability selection was paired with more complex algorithms for model fitting in an effort to improve predictions. For SVR models, we found that nonlinear kernels outperformed others (p < 0.01). No differences were observed when using 2^nd^ order and 3^rd^ order polynomial kernels with SVR (p > 0.05). SVR with polynomial kernel outperformed SVR with linear kernel, but gaussian SVR performed even better. Notably, correlation showed much smaller differences between SVR models, and many of these model differences were just under the cusp of significance (p < 0.05). Counterintuitively, correlation was higher for linear SVR than nonlinear SVR (p < 0.05). Although again we saw no difference between 2^nd^ and 3^rd^ order polynomial kernels when inspecting correlation, gaussian SVR did not perform better than polynomial SVR (p > 0.05), and linear SVR outperformed all of those kernels (p < 0.05). Moreover, linear SVR performed as well as GPR based on correlation (p > 0.05). GPR and all SVR models outperformed lasso (p < 0.05). Finally, patterns with respect to RFR mirrored what we observed with AP. For the full set of model comparisons, see Figure S11, S12 and Table S4.

**Figure S11:**
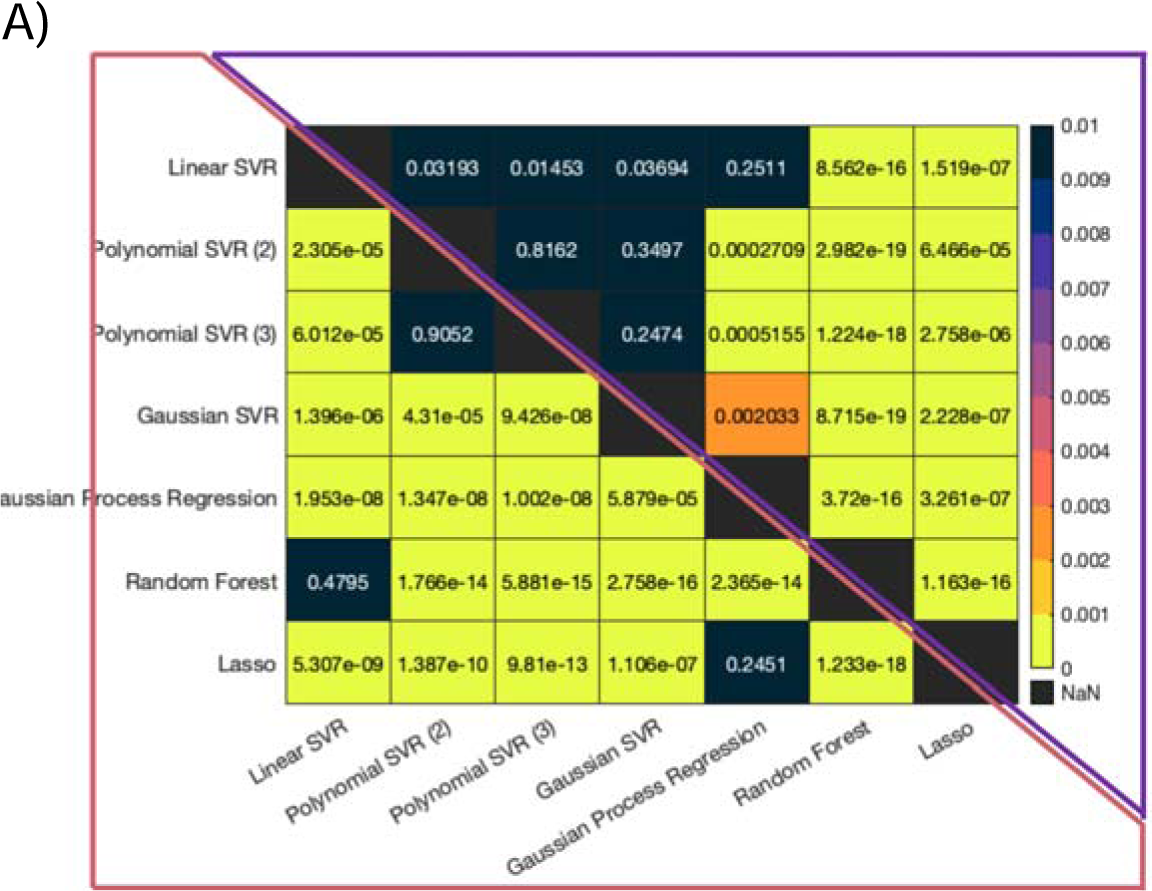
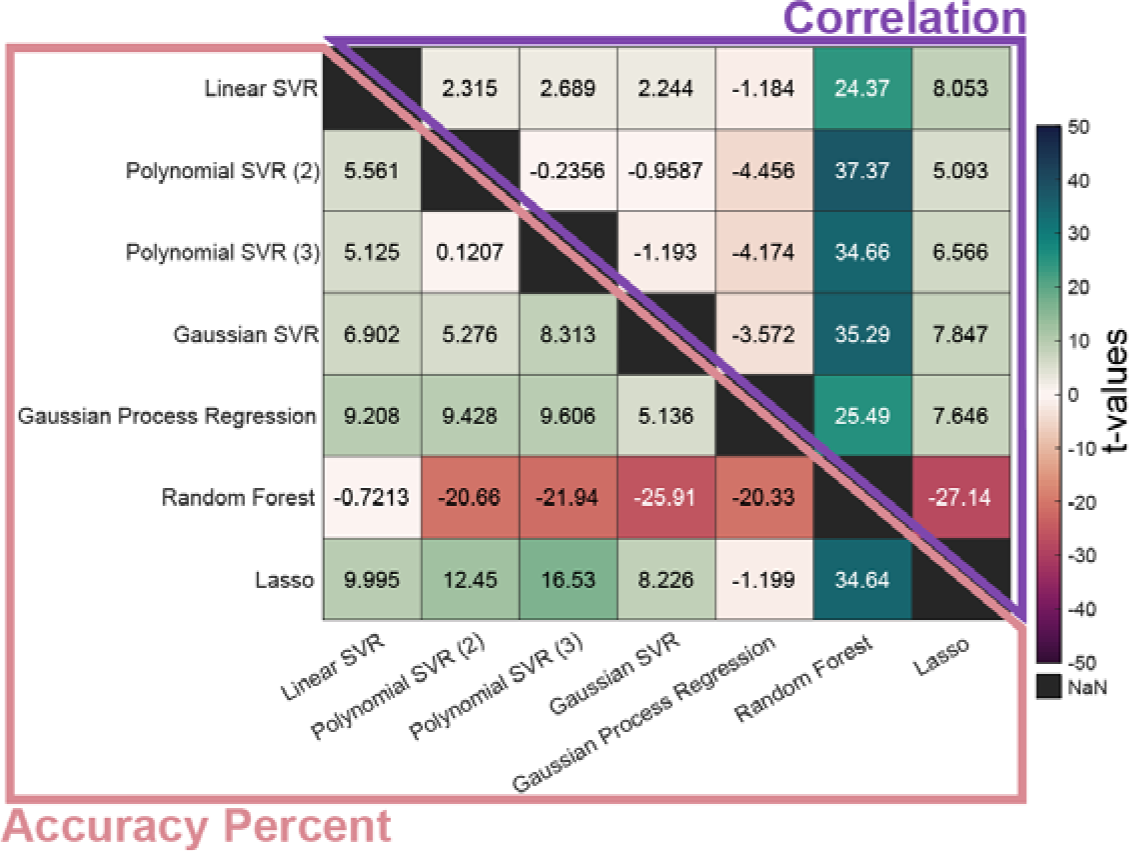
Comparing model building approaches that pair stability selection with more complex machine learning algorithms for model fitting. See previous Figure S2 for more information on how matrices are constructed. **Panel A:** Each cell shows the p-value for pairwise model comparisons according to a paired t-test. **Panel B:** Each cell shows the t-statistic for pairwise model comparisons. Note, the upper triangle in both panels shows comparisons based on correlation and the bottom triangle shows comparisons based on accuracy percent.

**Figure S12:**
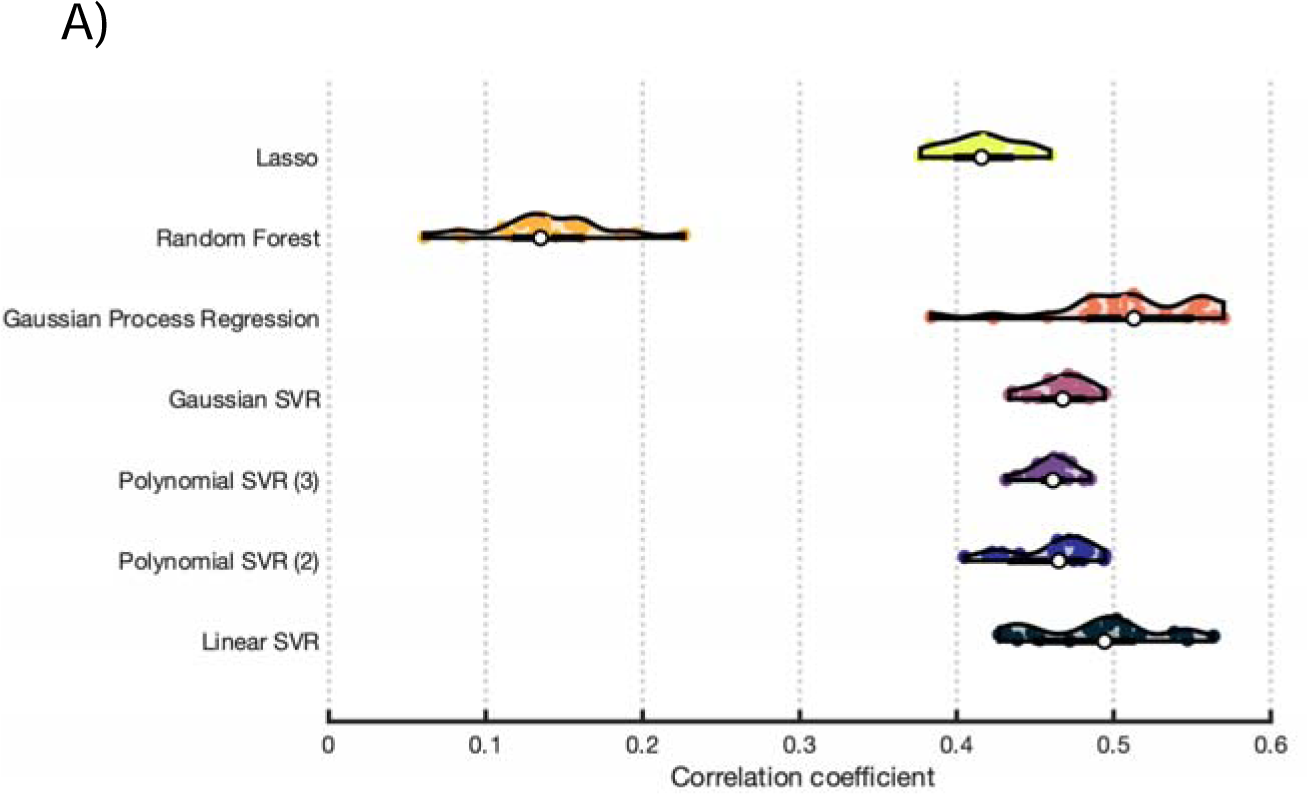
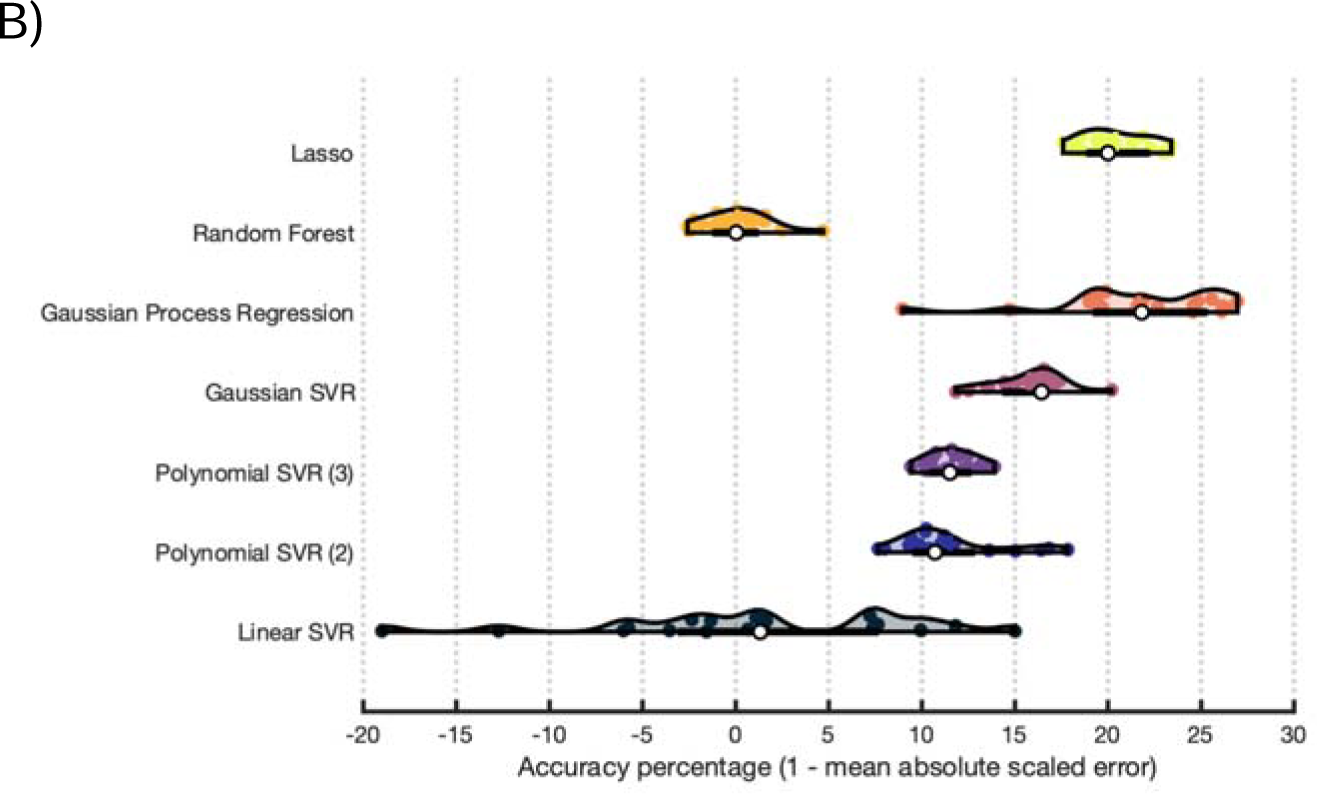
Violin plots showing distribution of model performance over repeats of cross-validation for stability selection paired with more complex machine learning algorithms for model estimation. **Panel A:** Performance is measured as correlation coefficient between true and predicted naming scores. **Panel B:** Performance is measured as accuracy percent. See Figure S3 for more information on the composition of violin plots.

**Table S4:**
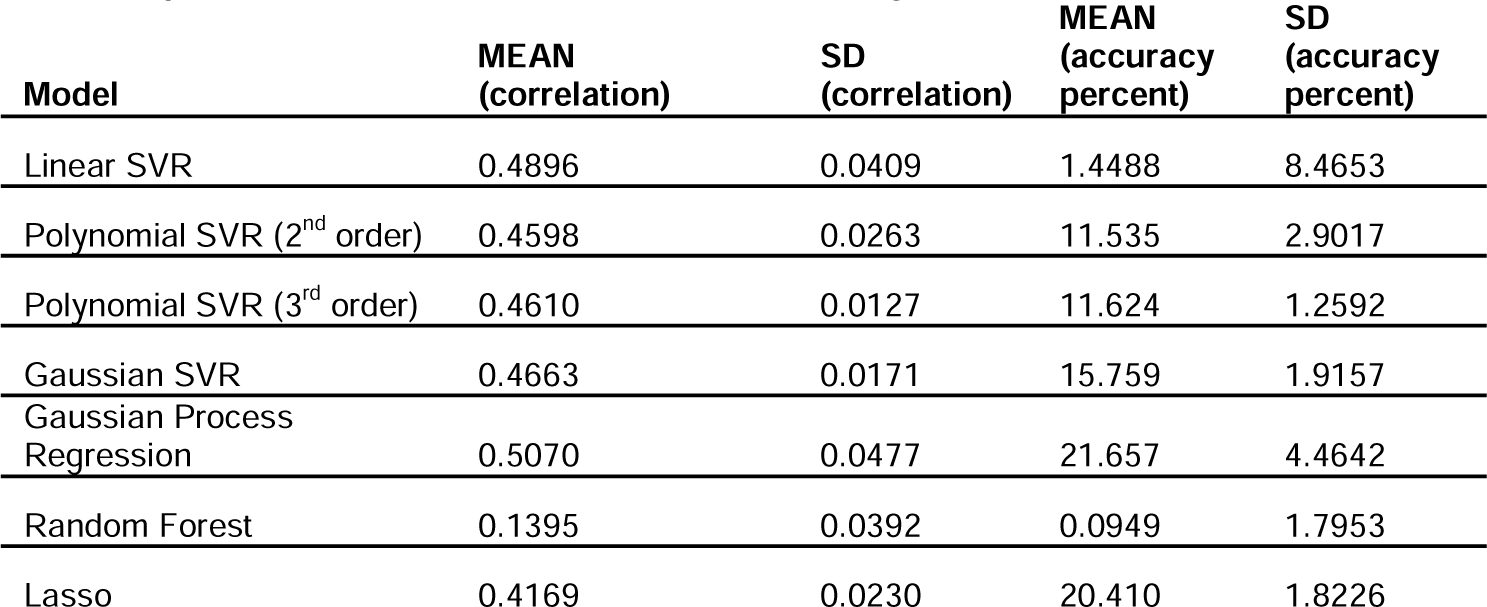
Mean and standard deviation of performance measures for models pairing stability selection with more complex algorithms for model estimation.

| Model | MEAN<br>(correlation) | SD<br>(correlation) | MEAN<br>(accuracy<br>percent) | SD<br>(accuracy<br>percent) |
| --- | --- | --- | --- | --- |
| Linear SVR | 0.4896 | 0.0409 | 1.4488 | 8.4653 |
| Polynomial SVR (2<sup>nd</sup> order) | 0.4598 | 0.0263 | 11.535 | 2.9017 |
| Polynomial SVR (3<sup>rd</sup> order) | 0.4610 | 0.0127 | 11.624 | 1.2592 |
| Gaussian SVR | 0.4663 | 0.0171 | 15.759 | 1.9157 |
| Gaussian Process<br>Regression | 0.5070 | 0.0477 | 21.657 | 4.4642 |
| Random Forest | 0.1395 | 0.0392 | 0.0949 | 1.7953 |
| Lasso | 0.4169 | 0.0230 | 20.410 | 1.8226 |

**Figure S13:**
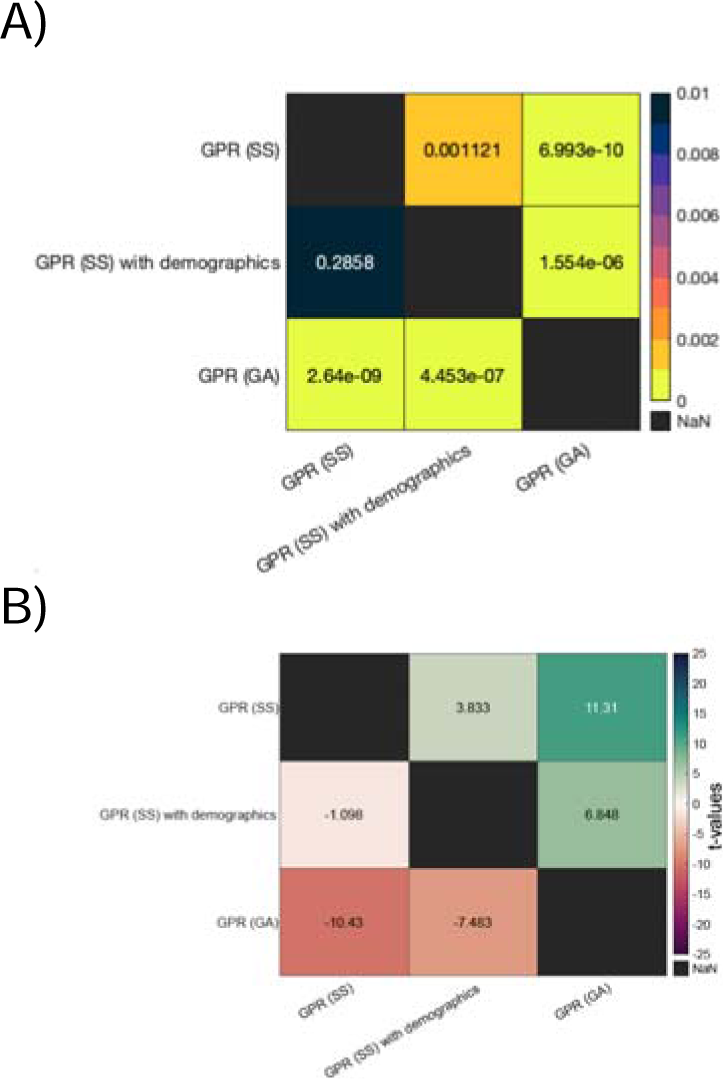
Comparing gaussian process regression models that include demographics and other successful feature selection approaches. In the panels, SS abbreviates stability selection and GA abbreviates genetic algorithm. See previous Figure S2 for more information on how matrices are constructed. **Panel A:** Each cell shows the p-value for pairwise model comparisons according to a paired t-test. **Panel B:** Each cell shows the t-statistic for pairwise model comparisons. Note, the upper triangle in both panels shows comparisons based on correlation and the bottom triangle shows comparisons based on accuracy percent.

**Table S5:** Mean and standard deviation of performance measures for gaussian process regression models that include demographics and other successful feature selection approaches.

| Model | MEAN<br>(correlation) | SD<br>(correlation) | MEAN<br>(accuracy<br>percent) | SD<br>(accuracy<br>percent) |
| --- | --- | --- | --- | --- |
| GPR (SS) | 0.5070 | 0.0477 | 21.657 | 4.4642 |
| GPR (SS) with<br>demographics | 0.4219 | 0.0833 | 19.674 | 6.5183 |
| GPR (GA) | 0.2544 | 0.0776 | 1.5063 | 7.8213 |

#### 2.8 Comparing stability selection to baseline models when paired with more complex algorithms for model fitting

In the main text, we compare GPR, SVR, and RFR with stability selection to the same algorithms applied to the entire feature set. We also compared the performance of the same algorithms when applied to theory-driven feature selection. As these algorithms took substantially longer to tune, only left hemisphere ROIs were used. This smaller set of ROIs tended to produce better performing models than bilateral ROIs in our previous analyses, especially in the context of algorithms that did not involve dimensionality reduction. See Figure S15, S16 and Table S6 for results. Using left hemisphere ROIs generally performed as well as using all features when fitting models with RF or GPR (p > 0.05). However, performance was slightly worse when fitting SVR to left hemisphere ROIs (p < 0.05). When left hemisphere ROIs were used, GPR performed as well as SVR (p > 0.05). The overall pattern which emerged from these model comparisons is that stability selection made a large impact on models whereas theory-driven feature selection had little to no impact. It’s possible that bilateral ROIs would have performed better, possibly exploiting interaction effects between hemispheres. However, we presented a comprehensive comparison between theory-driven and data-driven feature selection methods in Figure 3 of the main text to motivate stability selection in a more intuitive context. Our results do not suggest that theory-driven feature selection will always result in poorer models, but that data-driven feature selection can be highly successful. Ultimately, the purpose of the analysis presented in Figures S15 and S16 is to confirm that addition of stability selection can improve models fit using more complex algorithms (i.e., no feature selection).

**Figure S15:**
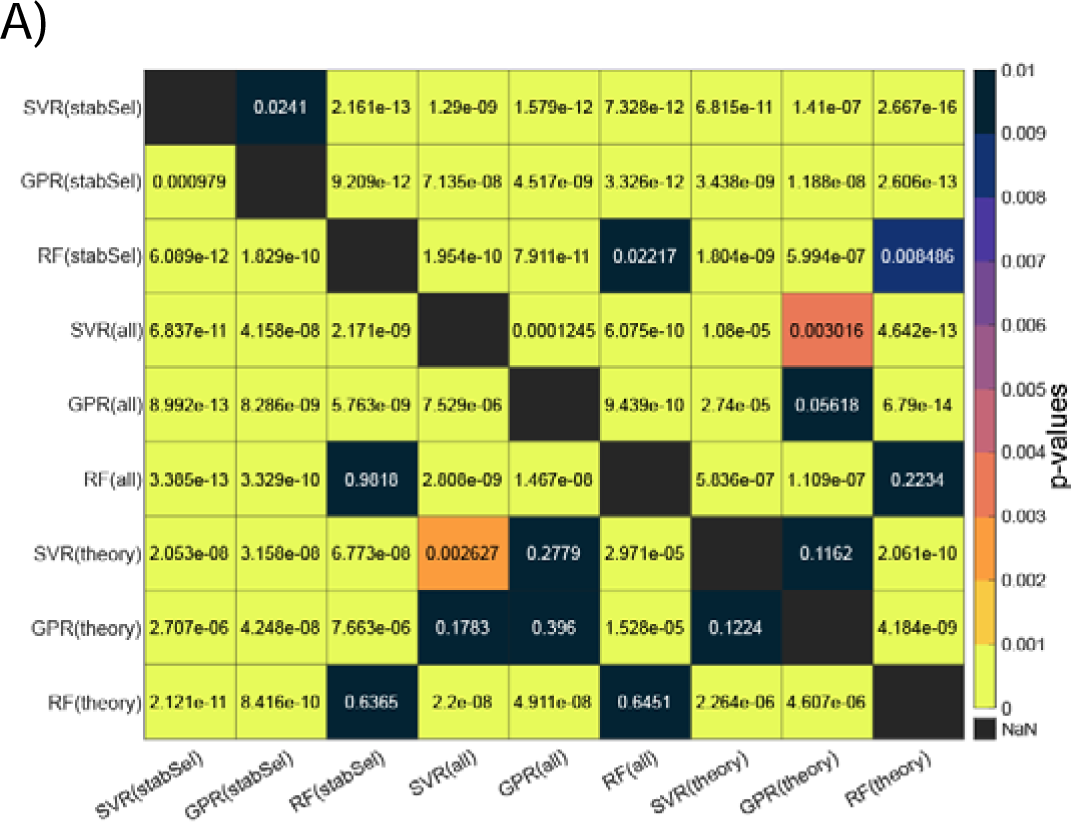
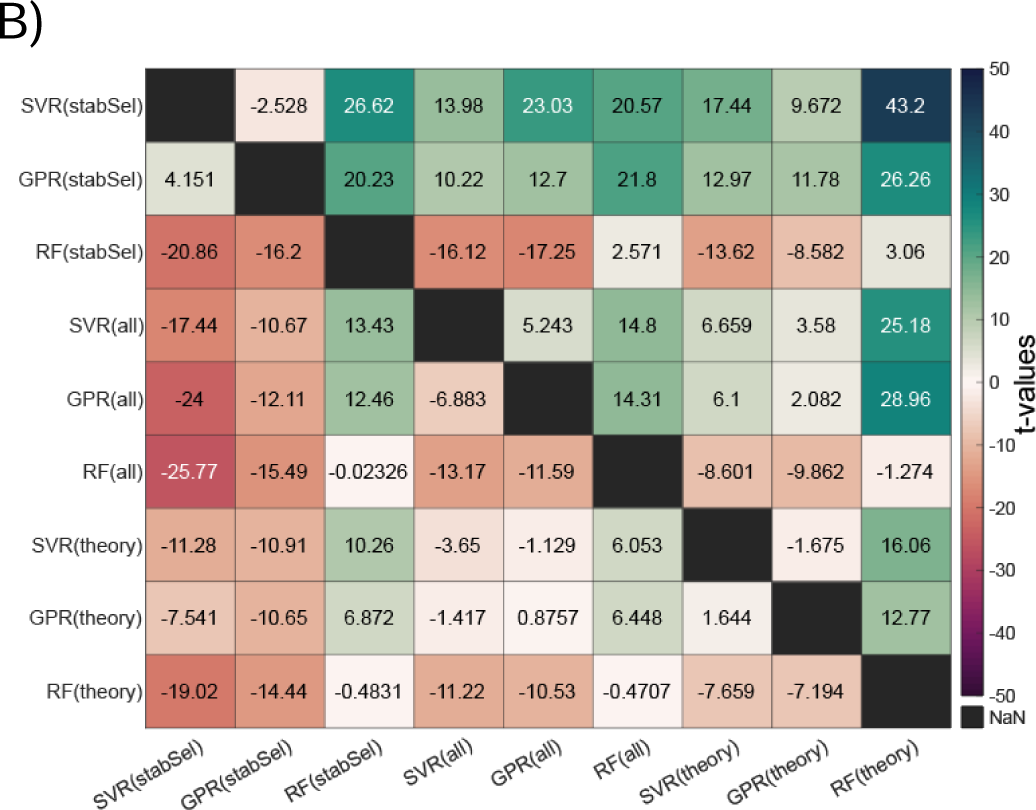
Comparisons between stability selection and baseline models when more complex machine learning algorithms are used for model fitting. In all panels, the type of feature selection for a given model fitting algorithm is labeled in parentheses. The ‘all’ models reflect models fit to all features (i.e., no feature selection). The ‘theory’ models reflect models fit to language regions of interest. See previous Figure S2 for more information on how matrices are constructed. Each label describes the feature selection method, with the method used to fit models described in parentheticals. **Panel A:** Each cell shows the p-value for pairwise model comparisons according to a paired t-test. **Panel B:** Each cell shows the t-statistic for pairwise model comparisons. Note, the upper triangle in both panels shows comparisons based on correlation and the bottom triangle shows comparisons based on accuracy percent.

**Figure S16:**
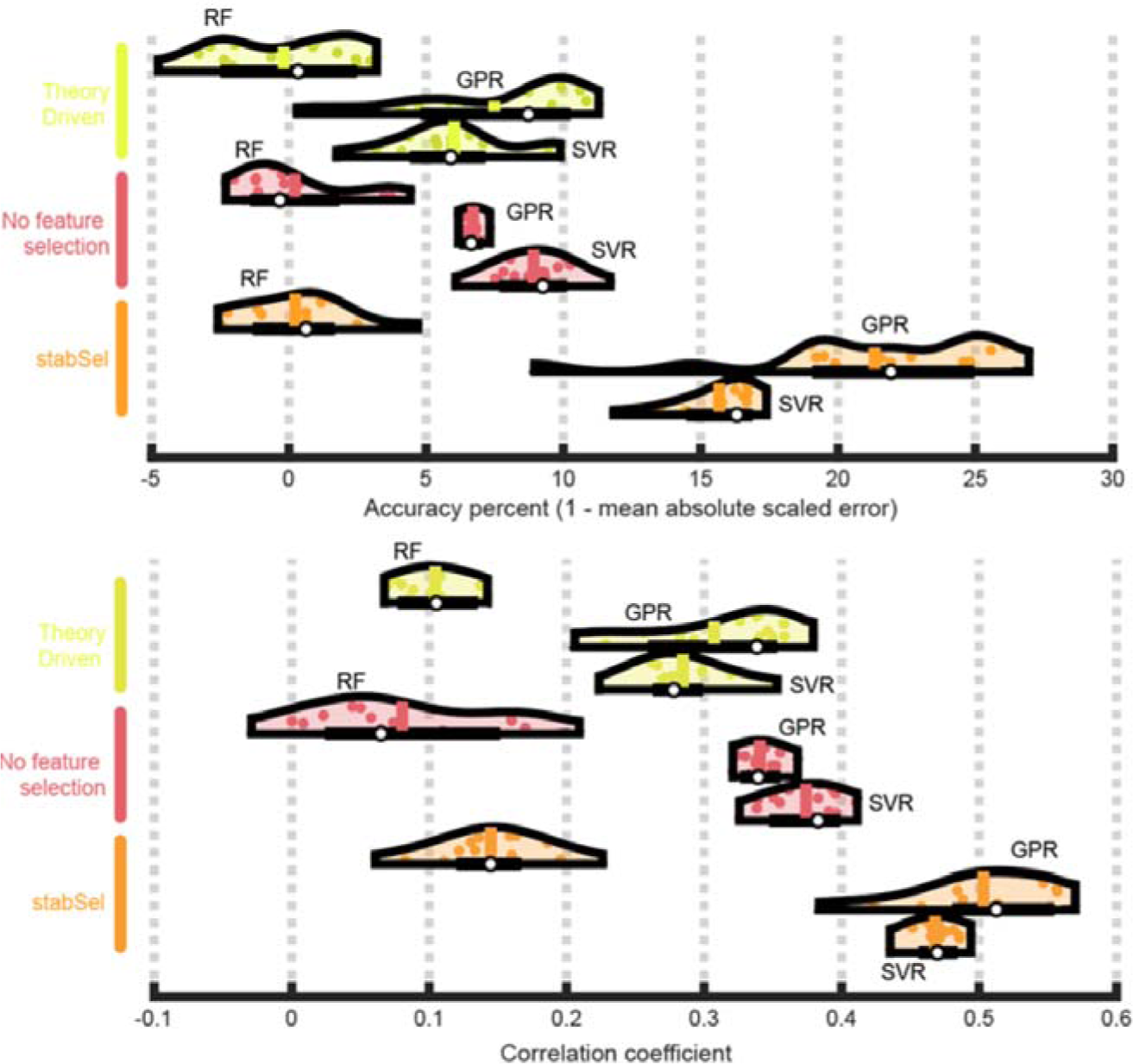
Violin plots showing distribution of model performance over repeats of cross-validation for stability selection paired with more complex machine learning algorithms for model estimation. Yellow violins show performance of models paired with theory-driven feature selection. Salmon violins show performance of models paired with no feature selection. Orange violins show performance of models paired with stability selection. Within each set of violins belonging to the same color, the topmost violin shows model estimation with a random forest (RF), the middle violin shows model estimation with gaussian process regression (GPR), and the bottom violin shows model estimation with support vector regression (SVR). **Top chart:** Performance is measured as accuracy percent. **Bottom chart:** Performance is measured as correlation between true and predicted naming scores. See Figure S3 for more information on the composition of violin plots.

**Table S6:**
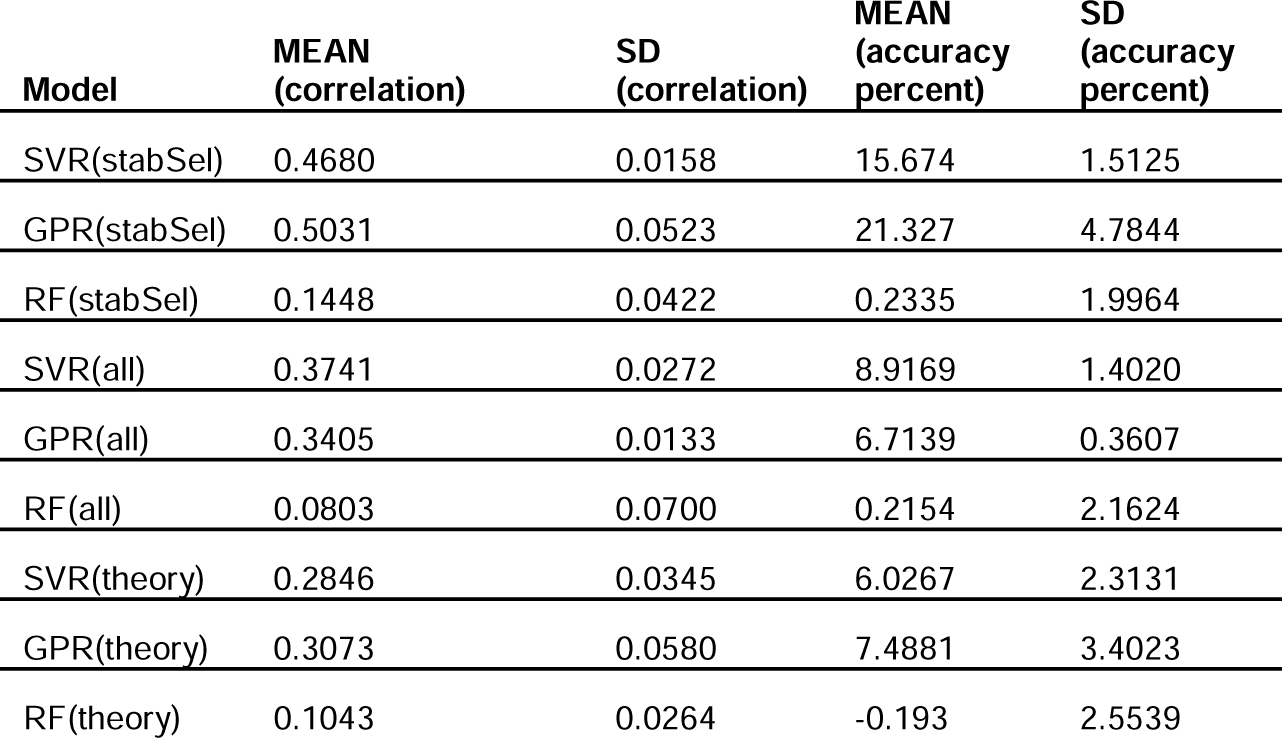
Mean and standard deviation of performance measures for baseline feature selection methods paired with more complex machine learning algorithms for model fitting.

| **Model** | **MEAN**<br>**(correlation)** | **SD**<br>**(correlation)** | **MEAN**<br>**(accuracy**<br>**percent)** | **SD**<br>**(accuracy**<br>**percent)** |
| --- | --- | --- | --- | --- |
| SVR(stabSel) | 0.4680 | 0.0158 | 15.674 | 1.5125 |
| GPR(stabSel) | 0.5031 | 0.0523 | 21.327 | 4.7844 |
| RF(stabSel) | 0.1448 | 0.0422 | 0.2335 | 1.9964 |
| SVR(all) | 0.3741 | 0.0272 | 8.9169 | 1.4020 |
| GPR(all) | 0.3405 | 0.0133 | 6.7139 | 0.3607 |
| RF(all) | 0.0803 | 0.0700 | 0.2154 | 2.1624 |
| SVR(theory) | 0.2846 | 0.0345 | 6.0267 | 2.3131 |
| GPR(theory) | 0.3073 | 0.0580 | 7.4881 | 3.4023 |
| RF(theory) | 0.1043 | 0.0264 | -0.193 | 2.5539 |

